# Low-dose computed tomography and rapid molecular sputum testing in adults with suspected non-ventilator-associated hospital-acquired pneumonia: the HAP-FAST pilot sequential multiple assignment randomised trial with embedded qualitative and costing studies

**DOI:** 10.64898/2026.11.10.26367346

**Authors:** DG Wootton, FC Sherratt, I Powell, ST Abrams, E Adegbenro, A Alvarez Nishio, SJ Aston, R Bazaz, H Clay, AM Collins, K Cook, A Cross, K Edwards, P Evans, F Fyles, N Goddard, AN Guldemond, J Hare, A Howard, J Howells, J Hucker-Shaw, K Irwin, T Jenks, K Kalapurakal, F Khalid, E Lam, D Mclaughlan, C McLeavy, B Pearce, L Pilling, I Poku, N Shafiqa, H Singh, A Tao, E Thwaite, S Trewick, L Turtle, S Willshaw, J Wood, B Young, A Jones

## Abstract

**Background:** Non-ventilator-associated hospital-acquired pneumonia (nv-HAP) diagnosis is imprecise; chest x-rays (CXR) lack detail and sputum culture delays targeted prescribing. However, the evidence base to support diagnostic improvement is sparse. We assessed the feasibility of a trial of low-dose thoracic CT and rapid molecular testing of sputum.

**Methods:** HAP-FAST was a UK NIHR funded, open-label pilot sequential multiple assignment randomised trial with qualitative and costing substudies in four UK hospitals (ClinicalTrials.gov:NCT05483309). Using research without prior consent (RWPC) and site-stratified web-based randomisation, clinicians allocated adults with suspected nv-HAP (excluding pregnancy and palliation), before their second antibiotic dose, 1:1 to CT or CXR; participants subsequently treated for nv-HAP with sputum available underwent another 1:1 randomisation to BIOFIRE FILMARRAY Pneumonia plus Panel (FAPP) with standard microbiology or standard microbiology alone. Feasibility outcomes included recruitment, consent, intervention delivery and data completeness through 90 days. Adverse events were not systematically collected. Analyses were intention-to-treat and descriptive.

**Results:** Between June 2023 and March 2024, 220 adults were randomised at stage-1 (109 CT; 111 CXR), reaching target three months early. Of 154/220 (70.0%) subsequently managed as nv-HAP, 92/154 (59.7%) could not produce sputum. 62/154 (40.3%; 28.2% of stage-1) were randomised to stage-2: 31 FAPP and 31 no-FAPP. Allocated imaging was recorded for 75/109 (68.8%) CT and 99/111 (89.2%) CXR participants; 6/31(19.4%) FAPP tests were not completed. Consent for follow-up was obtained for 142/220 (64.5%). Protocol-defined clinical cure was assessable in 1/220 participants. The day-90 resource-use questionnaire was returned by 55/110 participants (50.0%). Qualitative findings supported RWPC but identified needs for staged communication, lower-burden follow-up and improved diagnostic pathways. Excess bed days and readmissions drove costs but data capture requires refinement.

**Conclusions:** A definitive trial is plausible only with enhanced RWPC, diagnostic delivery and follow-up pathways, improved sputum procedures, stewardship-aligned outcomes and routine economic data capture.

**Key messages:** *What is already known on this topic:* Evidence to guide diagnostic strategies for suspected nv-HAP is limited, and the feasibility of evaluating sequential imaging and rapid sputum testing is uncertain.

*What this study adds:* HAP-FAST showed that recruitment to a sequential diagnostic trial was achievable, but identified important limitations in intervention delivery, follow-up and outcome assessment. Integrated qualitative and costing findings explained priorities for redesign.

*How this study might affect research, practice or policy:* A definitive trial should incorporate staged consent, reliable diagnostic pathways, lower-burden follow-up, feasible clinical and stewardship outcomes, and improved resource-use measurement.

## Introduction

Non-ventilator-associated hospital-acquired pneumonia (nv-HAP) is the most common healthcare-associated infection and causes substantial mortality, prolongs admission and increases costs.^1, 2^ Guidelines recommend broad-spectrum empirical antibiotics.^3^ Stewardship depends on correctly identifying nv-HAP as the cause of the patient’s deterioration and identifying pathogens early enough to guide treatment. However, chest x-rays (CXR) are imprecise, while sputum culture is slow and insensitive.^4, 5^

The 2025 National Institute for Health and Care Excellence (NICE) pneumonia guideline identified few microbial diagnostic studies and no eligible radiological diagnostic-strategy studies in nv-HAP; a systematic review found no randomised diagnostic trials.^3, 6, 7^ Low-dose computed tomography (CT) could reduce overdiagnosis and identify alternative causes of deterioration, while rapid molecular sputum testing could support earlier targeted anti-microbial prescribing.^7, 8^ Their evaluation requires a trial pathway compatible with urgent imaging and antibiotics. Evidence supporting these technologies comes largely from community-acquired pneumonia or critical-care populations, whose diagnostic pathways and case mix differ from ward-based nv-HAP. Establishing whether clinicians can recruit, deliver the tests and collect interpretable outcomes is therefore necessary before evaluating effectiveness.

Suspected nv-HAP arises across wards, often out of hours, in patients who may be older, frail or cognitively impaired.^1, 5, 9–11^ These circumstances constrain recruitment and intervention delivery. Research without prior consent (RWPC) offers a potential solution, but its implementation and acceptability in ward-based nv-HAP require evaluation.^12^ Outcome selection is also difficult: mortality, length of stay, antibiotic exposure, symptoms and quality of life are relevant, but attribution to nv-HAP is complicated by comorbidity.^13–15^

HAP-FAST assessed the feasibility of a definitive diagnostic trial.^16^ Embedded qualitative and costing substudies examined acceptability, barriers to trial delivery, diagnostic decision-making and economic data capture, to explain quantitative feasibility findings and inform future design.^17, 18^

## Methods

### Patient and public involvement

Patient and public representatives reviewed drafts of the original grant proposal and influenced the final study design. They contributed during protocol development by reviewing the protocol and study documents and suggesting changes. They served on trial oversight committees and contributed to interpretation of findings and manuscript review. A public contributor reviewed the qualitative topic guides before data collection. In addition, patients and their relatives and friends who had direct lived experience of participation in the trial participated in qualitative interviews. Patients and public representatives reviewed and commented upon this manuscript and are named authors.

### Study design and setting

HAP-FAST was an open-label pilot sequential multiple assignment randomised trial (SMART), coordinated by Liverpool Clinical Trials Centre (LCTC), in four acute hospitals in north-west England. The protocol was published previously.^16^ The target of 220 participants over 12 months was based on prospective audits at two centres, assuming 30% eligibility and 40% recruitment.^16^ The planned recruitment period allowed assessment across seasons and different hospital contexts.

Two sequential decisions were evaluated: stage-1 imaging (CXR versus low-dose thoracic CT) and stage-2 microbiological investigation with or without the BIOFIRE FILMARRAY Pneumonia plus Panel (FAPP). These defined four dynamic treatment regimens: CXR/no-FAPP, CXR/FAPP, CT/no-FAPP and CT/FAPP. Participants not entering stage-2 remained in follow-up and stage-1 analyses. Stage-2 randomisation followed the clinician’s decision to treat HAP, preserving the sequence of diagnostic decisions rather than assigning all participants to both interventions irrespective of clinical need.

### Participants and consent

Adults with suspected nv-HAP were eligible if initial trial procedures could be completed before their second antibiotic dose. In keeping with the definition of nv-HAP used by the UK NICE guidelines, the study included pneumonia developing at least 48 hours after admission or presentation after discharge from an overnight hospital stay within the preceding 10 days. Exclusions were prior CXR for suspected HAP, ventilator-associated pneumonia, inability to undergo CT, pregnancy, no intention to deliver antibiotics, or previous participation. Stage-2 eligibility required an intention to treat HAP and sputum availability before the second antibiotic dose. Baseline sex was self-reported (male/female).

RWPC permitted randomisation and interventions without delaying treatment.^12^ Written informed consent for continued participation was sought as soon as practicable; personal or professional consultees were involved when participants lacked capacity.

### Randomisation and interventions

An independent statistician generated site-stratified allocation schedules with variable block sizes, held centrally by LCTC and concealed until web-based allocation. Clinicians could confirm eligibility and randomise at any time. Stage-1 allocation was 1:1 to CXR or CT; stage-2 allocation was 1:1 to FAPP plus standard microbiology or standard microbiology alone. Allocations were immediately revealed and notified to clinicians because results informed care. After stage-1 imaging, managing clinicians interpreted CXRs and radiologists reported CTs urgently. For stage-2, instruments were placed in clinical areas and operated by clinicians; three sites used the FILMARRAY TORCH and one site used the FILMARRAY 2.0. FAPP results were interpreted using an antibiotic guideline approved by all sites. Participants in the no-FAPP arm received empirical antibiotics according to local guidelines.

After consent, CAP-Sym symptom and EQ-5D-5L quality-of-life questionnaires were collected where feasible.^19, 20^ Baseline clinical data included medical history and medication use. Daily reviews assessed clinical cure, with subsequent follow-up capturing symptoms, quality of life and further resource use. Participants were reviewed daily until day 10 or discharge, at day 28 in person and day 90 by telephone. Mechanistic research sampling is reported separately.

### Qualitative substudy

We purposively sampled patients, personal consultees and practitioners across sites, roles and participant characteristics. Patients and consultees were contacted at least two weeks after discharge for semi-structured interviews at home or by telephone. Practitioners were recruited through site investigators for face-to-face or online focus groups, supplemented by interviews to include under-represented roles or sites. A non-clinical qualitative researcher conducted all interviews and focus groups after separate informed consent. A public contributor reviewed topic guides before data collection; guides were iteratively refined during analysis.

Verbatim transcripts were checked against recordings and pseudonymised. Collaborative reflexive thematic analysis proceeded alongside data collection.^21^ The qualitative researcher led coding in NVivo whilst a clinician-trialist separately reviewed all transcripts. Patient, consultee and practitioner datasets were initially analysed separately, then compared across groups. Analytic discussions challenged and refined interpretations rather than establishing coding agreement. Findings are presented alongside quantitative process measures to explain feasibility constraints and identify design implications.

### Outcomes and analysis

The primary objective was feasibility, including recruitment, consent, representativeness, retention to day 90, intervention delivery, outcome completeness, acceptability, decision-making and economic data capture. Several potential outcome measures for a future fully powered trial were considered (hereafter referred to as ‘candidate outcomes’): clinical cure, antibiotic defined daily doses (DDD), EQ-5D-5L, length of stay and mortality at days 14, 28 and 90. The prespecified clinical cure outcome, developed from Weiss et al., required at least 35% improvement in symptoms measured by CAP-Sym, clinical stability using modified Halm’s criteria, and radiological stability or improvement at days 7-10.^14, 20, 22^ Additional imaging findings, FAPP pathogens and UK-AWaRe antibiotic categories were described.^23^ Feasibility assessment also considered descriptive estimates and dispersion of candidate outcomes, clinician willingness to randomise, human factors affecting test delivery and factors influencing follow-up. Formal progression thresholds were not prespecified; given the novelty and complexity of HAP-FAST, feasibility was assessed holistically rather than by applying retrospective numerical thresholds. Adverse events were not systematically collected, as prespecified in the protocol.

Analyses followed the prespecified statistical analysis plan (supplementary file) after 90-day follow-up, without interim analyses. Stage-specific comparisons were by intention-to-treat regardless of deviations and were descriptive, without hypothesis testing. Missingness was reported. Real-time eligibility confirmation replaced conventional screening logs, so population denominators for eligibility, approach and recruitment were unavailable. Representativeness was assessed against site-coded nv-HAP episodes in the routinely collected NHS Secondary Uses Service (SUS) database and the clinically validated Advancing Quality (AQ) sample, recognising possible ICD-10 miscoding.^24–26^ Analyses used SAS 9.4 and R 4.4.1.

### Costing analysis

This examined whether inpatient and post-discharge resource use could be captured sufficiently for a future cost-effectiveness evaluation. Costing adopted an NHS and personal social services perspective.^27^ Available inpatient data from two sites were costed using Healthcare Resource Group tariffs and excess bed-day prices based on the NHS Schedule of costs and local hospital finance data. A day-90 questionnaire captured primary care, readmissions, outpatient attendance, investigations, community and domiciliary care, and new medications. Costs covered observed follow-up, truncated at death and capped at 90 days; a 90-day window was assumed where dates were unavailable. Time-dependent items were scaled to observed duration. Readmissions used excess bed-day proxies; other contacts and medications used standard national unit costs with reference to the British National Formulary and NHS drug prices with ranges retained for sensitivity analyses. Return and item completeness were summarised, with available-case post-discharge totals.^18^

### Ethics, oversight and funding

NHS Research Ethics and Health Research Authority approval was obtained (22/WA/0315), following review by a panel configured for research involving adults lacking capacity. The University of Liverpool sponsored the trial, with independent steering committee oversight. Registration was ClinicalTrials.gov:NCT05483309. The National Institute for Health and Care Research funded the study (NIHR300669); bioMérieux loaned instruments and supplied panels. Neither organisation influenced design, conduct, analysis or preparation.

## Results

### Recruitment and consent

Between 13 June 2023 and 12 March 2024, 220 adults were randomised at stage-1 (109 CT; 111 CXR), reaching target three months early. Of 154/220 (70.0%) subsequently managed as nv-HAP, 92/154 (59.7%) could not produce sputum. Thus, 62/154 (40.3%; 28.2% of stage-1) entered stage-2: 31 FAPP and 31 no-FAPP (figure 1). This limits microbiological comparisons to sputum-producing patients managed as nv-HAP and has important implications for sample-size planning. The four sputum-producing pathways comprised CXR/FAPP (n=15), CXR/no-FAPP (n=16), CT/FAPP (n=16) and CT/no-FAPP (n=15); 50 CXR and 42 CT participants treated as nv-HAP followed no-sputum branches.

**Figure 1.**
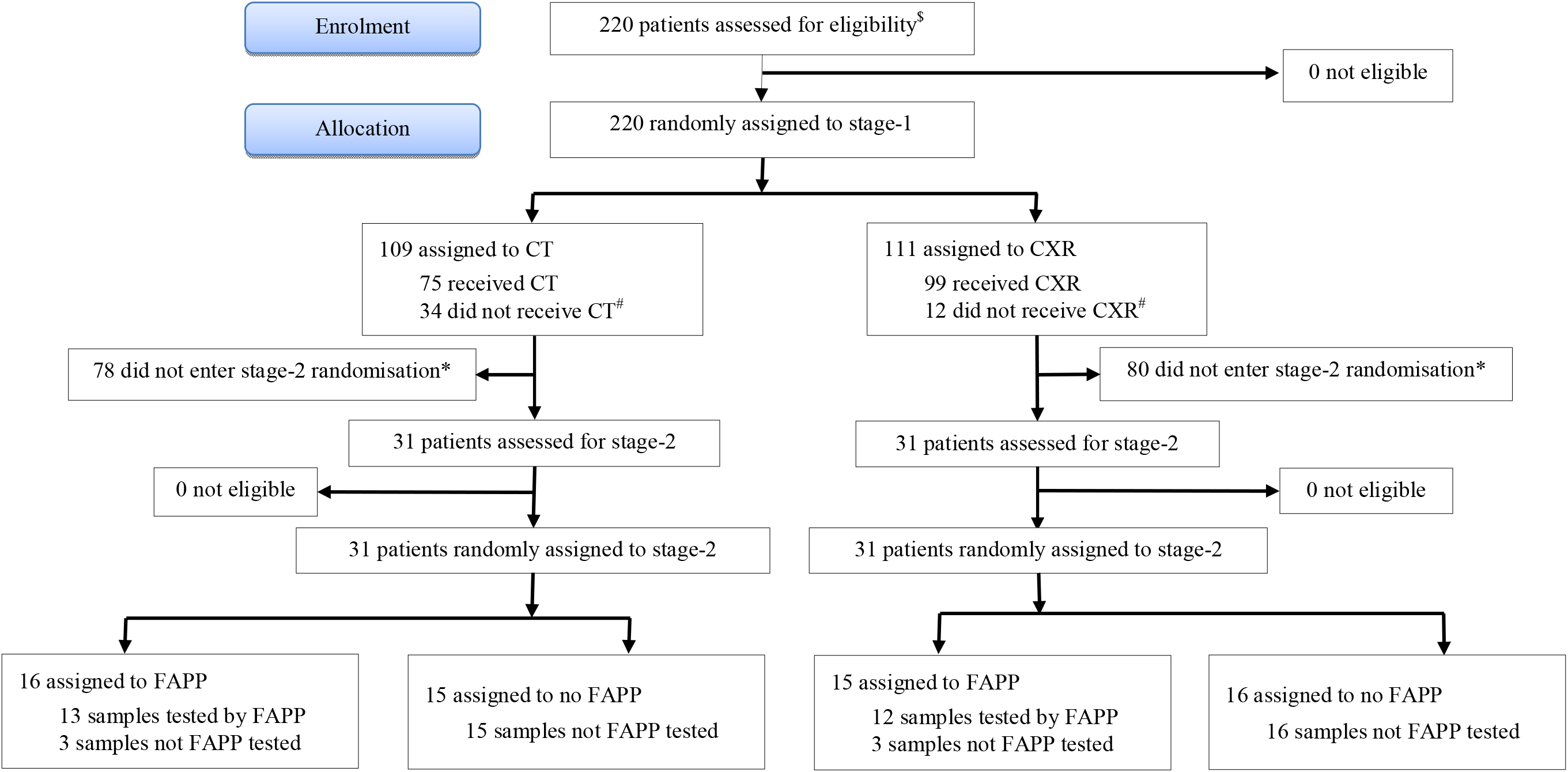
CONSORT flow diagram of participant pathways through the trial. See also Statistical Analysis Report (SAR) provided as supplement. CT = low dose, computed tomography of the thorax. CXR = chest x-ray. FAPP = BIOFIRE® FILMARRAY® Pneumonia Plus Panel. ^$^ Population denominators for all patients with suspected nv-HAP at the four sites were not available. Eligibility assessment here relates to pre-randomisation eligibility checks performed when a clinician interacted with the online randomisation system. Some patients will have been considered by clinicians but will have been clearly ineligible meaning an attempt to randomise on the system will not have occurred-these will not have been captured by the process. ^#^If allocated imaging section of CRF was incomplete this resulted in imaging recorded as “not received”. *Stage-2 randomisation required a decision to treat as HAP and for sputum to be obtained before the second antibiotic dose

Stage-1 characteristics were broadly balanced (table 1). Age and sex resembled comparator cohorts. Stage-2 participants were younger than the overall cohort (median 77 years, IQR 69-84): medians were 74 years (64-80) in the FAPP arm and 73 years (65.5-81.5) in the no-FAPP arm.

**Table 1:** Baseline characteristics, clinical status and admission details for HAP-FAST participants and comparator cohorts.

| Characteristic | Category/statistic | CT (n=109) | CXR (n=111) | FAPP (n=31) | No-FAPP (n=31) | AQ (n=670) | SUS (n=3015) |
|---|---|---|---|---|---|---|---|
| **Demographics** |  |  |  |  |  |  |  |
| Age (years) | Median (IQR) | 77.0 (70.0, 83.0) | 77.0 (67.0, 85.0) | 74.0 (64.0, 80.0) | 73.0 (65.5, 81.5) | 77.0 (68.0, 85.0) | 76.0 (66.0, 85.0) |
|  | Missing n (%) | 17 (15.6) | 10 (9.0) | 0 (0) | 3 (9.7) | 0 (0) | 0 (0) |
| Sex: n (%) | Male | 53 (48.6) | 54 (48.6) | 13 (41.9) | 16 (51.6) | 352 (52.5) | 1592 (52.8) |
|  | Female | 56 (51.4) | 57 (51.4) | 18 (58.1) | 15 (48.4) | 318 (47.5) | 1423 (47.2) |
|  | Missing | 0 (0) | 0 (0) | 0 (0) | 0 (0) | 0 (0) | 0 (0) |
| **Comorbidities and smoking status** |  |  |  |  |  |  |  |
| Comorbidity: n (%) | None of below | 26 (23.9) | 30 (27.0) | 16 (51.6) | 8 (25.8) | 149 (22.2) | 594 (19.7) |
|  | Congestive cardiac failure | 12 (11.0) | 11 (9.9) | 1 (3.2) | 3 (9.7) | 9 (1.3) | 60 (2.0) |
|  | Dementia | 6 (5.5) | 7 (6.3) | 1 (3.2) | 0 (0.0) | 53 (7.9) | 257 (8.5) |
|  | Other form of cognitive impairment | 9 (8.3) | 4 (3.6) | 0 (0.0) | 2 (6.5) | 0 (0.0) | 0 (0.0) |
|  | Myocardial infarction | 6 (5.5) | 4 (3.6) | 0 (0.0) | 3 (9.7) | 61 (9.1) | 334 (11.1) |
|  | Chronic pulmonary disease | 30 (27.5) | 36 (32.4) | 8 (25.8) | 12 (38.7) | 260 (38.8) | 1191 (39.5) |
|  | Rheumatic disease | 1 (0.9) | 3 (2.7) | 0 (0.0) | 1 (3.2) | 20 (3.0) | 110 (3.6) |
|  | Advanced HIV | 0 (0.0) | 0 (0.0) | 0 (0.0) | 0 (0.0) | 0 (0.0) | 0 (0.0) |
|  | Hemiplegia or paraplegia | 0 (0.0) | 1 (0.9) | 0 (0.0) | 1 (3.2) | 18 (2.7) | 71 (2.4) |
|  | Renal disease | 13 (11.9) | 21 (18.9) | 2 (6.5) | 1 (3.2) | 154 (23.0) | 717 (23.8) |
|  | Liver disease | 1 (0.9) | 1 (0.9) | 0 (0.0) | 0 (0.0) | 34 (5.1) | 160 (5.3) |
|  | Diabetes | 23 (21.1) | 10 (9.0) | 5 (16.1) | 4 (12.9) | 110 (16.4) | 539 (17.9) |
|  | Any malignancy | 9 (8.3) | 17 (15.3) | 1 (3.2) | 1 (3.2) | 107 (16.0) | 556 (18.4) |
| Smoking: n (%) | Current smoker | 9 (8.3) | 19 (17.1) | 4 (12.9) | 5 (16.1) | 101 (15.1) | NA |
|  | Ex-smoker | 36 (33) | 39 (35.1) | 14 (45.2) | 11 (35.5) | NA | NA |
|  | Never smoked | 27 (24.8) | 27 (24.3) | 4 (12.9) | 7 (22.6) | NA | NA |
|  | Unknown | 20 (18.3) | 15 (13.5) | 9 (29.0) | 5 (16.1) | NA | NA |
|  | Missing | 37 (33.9) | 26 (23.4) | 9 (29.0) | 8 (25.8) | NA | NA |
| **Medical status prior to HAP suspicion and NEWS2 score at randomisation** |  |  |  |  |  |  |  |
| NEWS2 score | Median (IQR) | 2.0 (1.0-5.0) | 3.0 (1.0-5.0) | 3.5 (2.0, 7.0) | 3.0 (1.5, 6.0) | 3.0 (1.0,4.0) | NA |
|  | **Total missing n (%)** | 19 (17.4) | 13 (11.7) | 1 (3.2) | 3 (9.7) | 6 (0.9) | NA |
|  | Of which: vital signs not collected n (%) | 2 (1.8) | 0 | 1 (3.2) | 0 | 0 | NA |
|  | Of which: NEWS2 not known n (%) | 0 | 2 (1.8) | 0 | 0 | 0 | NA |
| Medically fit: n (%) | No | 62 (56.9) | 56 (50.5) | 18 (58.1) | 16 (51.6) | 623 (93.0) | NA |
|  | Yes | 6 (5.5) | 13 (11.7) | 1 (3.2) | 0 (0.0) | 47 (7.0) | NA |
|  | Not known | 24 (22.0) | 31 (27.9) | 12 (38.7) | 12 (38.7) | 0 (0.0) | NA |
|  | Missing | 41 (37.6) | 42 (37.8) | 12 (38.7) | 15 (48.4) | 0 (0.0) | NA |
| **Details of original admission to hospital** |  |  |  |  |  |  |  |
| Ward: n (%) | Medical | 48 (44.0) | 48 (43.2) | 6 (19.4) | 11 (35.5) | 170 (25.4) | 1072 (35.6) |
|  | Surgical | 13 (11.9) | 12 (10.8) | 7 (22.6) | 3 (9.7) | 84 (12.5) | 465 (15.4) |
|  | A&E | 28 (25.7) | 34 (30.6) | 18 (58.1) | 9 (29.0) | 2 (0.3) | 2 (0.07) |
|  | Acute medical unit | 2 (1.8) | 4 (3.6) | 0 (0.0) | 3 (9.7) | 71 (10.6) | 379 (12.6) |
|  | Other | 1 (0.9) | 3 (2.7) | 0 (0.0) | 2 (6.5) | 1 (0.1) | 19 (0.6) |
|  | Not known | 0 (0.0) | 0 (0.0) | 0 (0.0) | 0 (0.0) | 81 (12.1) | 350 (11.6) |
|  | Missing | 17 (15.6) | 10 (9.0) | 0 (0.0) | 3 (9.7) | 261 (39.0) | 728 (24.2) |
| Admission reason: n (%) | Medical | 80 (73.4) | 89 (80.2) | 26 (83.9) | 24 (77.4) | 495 (73.9) | 2241 (74.3) |
|  | Surgical | 12 (11.0) | 11 (9.9) | 5 (16.1) | 4 (12.9) | 159 (23.7) | 702 (23.3) |
|  | Missing | 17 (15.6) | 11 (9.9) | 0 (0.0) | 3 (9.7) | 16 (2.4) | 72 (2.4) |
| Surgery subtype: n (% surgery) | Elective | 5 (41.7) | 5 (45.5) | 2 (40.0) | 1 (25.0) | 56 (35.2) | 301 (42.9) |
|  | Emergency | 6 (50.0) | 5 (45.5) | 3 (60.0) | 3 (75.0) | 97 (61.0) | 379 (54.0) |
|  | Missing | 1 (8.3) | 1 (9.1) | 0 (0.0) | 0 (0.0) | 6 (3.8) | 22 (3.1) |
| Surgery site: n (% surgery) | Trauma | 1 (8.3) | 2 (18.2) | 1 (20.0) | 0 (0.0) | 60 (37.7) | 236 (33.6) |
|  | Head and neck | 0 (0.0) | 0 (0.0) | 0 (0.0) | 0 (0.0) | 16 (10.1) | 55 (7.8) |
|  | Eye | 0 (0.0) | 0 (0.0) | 0 (0.0) | 0 (0.0) | 0 (0.0) | 0 (0.0) |
|  | Chest | 0 (0.0) | 0 (0.0) | 0 (0.0) | 0 (0.0) | 2 (1.3) | 106 (15.1) |
|  | Abdomen | 4 (33.3) | 3 (27.3) | 2 (40.0) | 0 (0.0) | 39 (24.5) | 168 (23.9) |
|  | Pelvis/urology | 2 (16.7) | 0 (0.0) | 0 (0.0) | 1 (25.0) | 4 (2.5) | 15 (2.1) |
|  | Upper limb | 0 (0.0) | 0 (0.0) | 0 (0.0) | 0 (0.0) | 0 (0.0) | 1 (0.1) |
|  | Lower limb | 1 (8.3) | 2 (18.2) | 2 (40.0) | 1 (25.0) | 4 (2.5) | 18 (2.6) |
|  | Other | 4 (33.3) | 4 (36.4) | 0 (0.0) | 2 (50.0) | 32 (20.1) | 103 (14.7) |
|  | Missing | 0 (0.0) | 0 (0.0) | 0 (0.0) | 0 (0.0) | 0 (0.0) | 0 (0.0) |

CT = low-dose non-contrast enhanced thoracic computed tomography scan. CXR = chest x-ray. FAPP = BIOFIRE FILMARRAY Pneumonia plus Panel. SUS = UK NHS Secondary Uses Service database. AQ = Advancing Quality HAP database. NA = data fields marked NA indicate this data is not available from that source

Consent or consultee agreement for follow-up was obtained for 142/220 (64.5%); 77 did not provide consent for follow-up and consent could not be obtained for one. Recruitment averaged 24.4 participants/month, with site rates of 3.8-13.0. Detailed processes are reported in the Statistical Analysis Report (SAR, supplementary).

Qualitative data comprised 23 patient-linked interviews with 24 participants (20 patients and four personal consultees), including one joint patient–consultee interview. Twenty practitioners participated in four site-based focus groups and four individual interviews. Four themes contextualised trial delivery; illustrative quotations are in table 2. The first theme was conditional acceptance of recruitment. Patients and consultees supported participation perceived as low-burden, clinically sensible and optional, but decisions during acute illness were often trust-based, altruistic or instinctive rather than demonstrating detailed understanding. The second concerned RWPC: this was broadly considered necessary, but acceptability depended on staged communication, later recap, clearer consultee support and separation of research from routine care. These findings contextualise consent rates without equating initial recruitment with informed willingness to continue. Practitioner accounts distinguished objections to later study demands from objections to RWPC itself. Patients and relatives described the need to revisit information because explanations given during acute illness might not be retained. Thus, a successful first approach did not remove the need for subsequent support and clarification.

**Table 2:** Illustrative quotations supporting the integrated feasibility findings.

| Theme | Quote |
|---|---|
| **Recruitment was acceptable but conditional** | **P1**<br>“a lot of it is based on trust so I read that and I go yes that sounds ok to me, I trust you to deal with my information.” |
| **Research without prior consent required staged communication** | **HP14**<br>“I was very surprised…at how acceptable it has been from our participants though. I haven’t had any pushback…. even the patients who have withdrawn from the study the matter of it being deferred consent hasn’t been an issue at all when speaking to them about it, they’re withdrawing for other reasons.”<br>**P18+consultee**<br>[Mother]: … the next time you see the people then they should reiterate everything they said on the first meeting because everything doesn't sink in on the first meeting, does it? |
| **Retention barriers were cumulative** | **HP7**<br>“….patients said to me oh its too much for the day. My priority is to recover you have to slow down, its too much and two of them, because of that particular reason they came off from the study”<br>**P14**<br>“It’s just unfortunate that the parking at [Site 03] is diabolical.” |
| **Intervention delivery depended on pathway fit** | **HP18**<br>“it's the transfer. It's getting them down. On the face of it, like I say, the scan is really quick, but if they have to come on a bed from one of the wards, which is far away, it takes two porters”<br>**P14**<br>I would have felt more discomfort lying down flat on a CT, in a scanner, rather than just sitting on the chair having an x□ray of my chest.<br>**P1**<br>I did save some phlegm in a little bowl but I think that ended up in the bin and nobody collected that<br>**HP19**<br>“….obtain the sample, inform the clinical team of what's going on, go to the machine, which was in another ward at the other side of the hospital, analyse it… do all this process. And then the result …..it is stored in the machine and then you have to go with a USB and just transfer it to the USB and then feedback the result, back to the clinical team.” |

P denotes a participant identifier; HP denotes a health-practitioner identifier. Family and focus-group labels are reproduced as supplied. Repeated speaker identifiers refer to separate excerpts, not additional participants. Wording and existing ellipses are unchanged from the source transcripts.

### Retention and measurement burden

Among consented participants, 28/142 (19.7%) completed the protocol pathway and 25/142 (17.6%) withdrew from intervention or follow-up. Questionnaire completion declined over time in part due to withdrawals. Among expected assessments, EQ-5D-5L return was 104/142 (73.2%) at baseline and 37-45% subsequently. CAP-Sym completion was 66/74 (89.2%) at baseline, 72/132 (54.5%) at day 10 or discharge, and 42-45% at days 28 and 90. The day-90 cost questionnaire was returned by 55/110 (50.0%); item completion varied (SAR tables 12-20 to 12-22).

The third qualitative theme explained cumulative retention barriers. Participants and practitioners described repeated questionnaires and blood tests, travel, parking and uncertainty about appointment purpose as disincentives and reasons for withdrawal. Accounts included participants prioritising unencumbered recovery over multiple assessments, and practical difficulties attending hospital after discharge. The qualitative findings therefore suggest both the content of follow-up and delivery arrangements require refinement; merely reminding participants to attend would not address all the reported barriers.

### Intervention delivery

Allocated imaging was recorded as received in 75/109 (68.8%) participants allocated to CT and 99/111 (89.2%) allocated to CXR. Incomplete imaging forms were classified as non-receipt in the SAR (17 CT; 11 CXR); other reasons included operational constraints, intolerance and withdrawal. Timing analyses included any post-randomisation chest imaging within each arm, with data for 89/109 CT and 98/111 CXR participants. Median time to imaging was 2 hours CT (IQR 1-4) versus 1 hour CXR (1-2); time to formal reporting was 4 hours (3-13) versus 50 hours (18-135). Initial radiology reports were available for 188/220 participants. Based on modality received, common additional findings recorded in CT reports were emphysema and pulmonary nodules (20% each), bronchiectasis (17%) and atelectasis (14%); those in CXR reports were emphysema (9%), pleural effusion (6%), atelectasis (5%) and heart failure (4%). During 90-day follow-up, 27/188 participants (14.4%) underwent additional imaging: 19/112 (17.0%) after initial CXR and 8/76 (10.5%) after initial CT.

Six of 31 allocated FAPP tests were not completed: three misplaced samples, two invalid results and one unavailable sample. The most common FAPP detections were *Staphylococcus aureus*, *Haemophilus influenzae*, rhinovirus/enterovirus, *Pseudomonas aeruginosa* and *Klebsiella pneumoniae group* (Supplementary Table 1).

The fourth qualitative theme concerned pathway fit. CT delivery was constrained by transfer logistics and tolerance of lying flat, rather than rejection of the trial concept or lack of equipoise. FAPP delivery required successful sputum collection, sample handling, access to equipment, testing ownership and communication of results to clinical teams. These accounts explain how rapid tests can remain operationally fragile despite short analytical turnaround and identify modifiable steps between allocation and treatment decisions. Staff valued the prospect of pathogen-directed treatment, and patients and consultees supported appropriate antibiotic use. However, practitioners described the work involved in obtaining, analysing, transferring and interpreting the results, suggesting that influence on prescribing depended on information being early, trusted and linked to clinical review.

### Candidate outcomes

Table 3 summarises candidate trial outcome measures. Protocol-defined clinical cure was assessable in only 1/220 participants; 219/220 (99.5%) lacked assessment, mainly because repeat imaging was unavailable at days 7-10. A post-hoc definition omitting radiology yielded cure in 15/109 (13.8%) CT and 12/111 (10.8%) CXR participants, and 9/31 (29.0%) FAPP and 7/31 (22.6%) no-FAPP participants. Substantial non-assessment remained: 73/109 CT and 67/111 CXR participants, and 8/31 FAPP and 15/31 no-FAPP participants, could not be assessed using the relaxed definition. Reasons for non-assessment including missing symptom or consent-related data are detailed in SAR tables 12-4 and 12–19.

**Table 3:** Potential outcome measures assessed for a future definitive HAP-FAST trial, by randomised comparison.

| Outcome | Statistic / time point | CT | CXR | FAPP | No-FAPP |
|---|---|---|---|---|---|
| **Clinical cure** |  |  |  |  |  |
| Protocol-defined clinical cure | Achieved clinical cure | 0/109 (0.0) | 0/111 (0.0) | 0/31 (0.0) | 0/31 (0.0) |
|  | Failed clinical cure | 0/109 (0.0) | 1/111 (0.9) | 0/31 (0.0) | 1/31 (3.2) |
|  | Could not be assessed | 109/109 (100.0) | 110/111 (99.1) | 31/31 (100.0) | 30/31 (96.8) |
| Relaxed clinical cure* | Achieved clinical cure | 15/109 (13.8) | 12/111 (10.8) | 9/31 (29.0) | 7/31 (22.6) |
|  | Failed clinical cure | 21/109 (19.3) | 32/111 (28.8) | 14/31 (45.2) | 9/31 (29.0) |
|  | Could not be assessed | 73/109 (67.0) | 67/111 (60.4) | 8/31 (25.8) | 15/31 (48.4) |
| **Antibiotic usage for the HAP episode in defined daily doses** |  |  |  |  |  |
| DDD | N | 109 | 111 | 31 | 31 |
|  | Mean (SD) | 4.0 (7.2) | 4.8 (9.1) | 7.1 (9.4) | 5.4 (7.9) |
|  | Median (IQR) | 0.0 (0.0, 6.1) | 0.0 (0.0, 6.2) | 5.1 (0.0, 10.0) | 2.0 (0.0, 7.5) |
|  | Missing | 21 (19.3) | 10 (9.0) | 7 (22.6) | 8 (25.8) |
| **Antibiotic usage for the HAP episode by UK-AWaRe classification** |  |  |  |  |  |
| UK-AWaRe classification usage§ | Any Access antibiotic, n (%) | 49 (45.0%) | 55 (49.5%) | 15 (48.4%) | 18 (58.1%) |
|  | Any Watch antibiotic, n (%) | 54 (49.5%) | 47 (42.3%) | 18 (58.1%) | 16 (51.6%) |
|  | Any Reserve antibiotic, n (%) | 6 (5.5%) | 9 (8.1%) | 8 (25.8%) | 0 (0.0%) |
|  | Any Watch or Reserve antibiotic, n (%) | 56 (51.4%) | 50 (45.0%) | 20 (64.5%) | 16 (51.6%) |
|  | Any meropenem, n (%) | 6 (5.5%) | 9 (8.1%) | 8 (25.8%) | 0 (0.0%) |
| **Quality of life: EQ-5D-5L visual analogue scale (VAS)** |  |  |  |  |  |
| VAS score | Baseline: N expected; n observed; mean (SD); n missing | 68; 45; 44.1 (29.0); 23 | 74; 56; 48.7 (26.8); 18 | 28; 23; 45.3 (24.3); 5 | 26; 24; 40.3 (26.4); 2 |
|  | Day 10: N expected; n observed; mean (SD); n missing | 61; 21; 52.5 (26.2); 40 | 71; 26; 56.6 (24.1); 45 | 27; 10; 46.7 (17.5); 17 | 23; 12; 49.4 (25.5); 11 |
|  | Day 28: N expected; n observed; mean (SD); n missing | 55; 21; 66.7 (19.4); 34 | 64; 28; 60.1 (28.3); 36 | 26; 11; 54.6 (28.2); 15 | 19; 10; 58.5 (26.7); 9 |
|  | Day 90: N expected; n observed; mean (SD); n missing | 51; 20; 59.7 (24.4); 31 | 59; 23; 59.3 (25.7); 36 | 26; 11; 66.4 (24.5); 15 | 17; 7; 50.0 (22.9); 10 |
|  | Change from baseline to day 90: N expected; n observed; mean (SD); missing | 51; 16; 7.7 (33.0); 35 | 59; 21; 5.4 (30.8); 38 | 26; 10; 25.9 (21.1); 16 | 17; 7; 15.7 (43.4); 10 |
| **Length of hospital stay after suspected HAP** |  |  |  |  |  |
| Length of stay (days)† | N | 109 | 111 | 31 | 31 |
|  | Median (IQR) | 19 (8, 52) | 13 (6, 42) | 9 (7, 16) | 17 (9, ‡) |
|  | Range | (0, 176) | (0, ‡) | (0, 45) | (2, ‡) |
| **Mortality and Kaplan-Meier survival** |  |  |  |  |  |
| Mortality | Day 14: deaths n (%);<br>KM survival (95% CI); censored | 5/109 (4.6);<br>93.3 (84.4, 97.2); 44 | 6/111 (5.4);<br>92.9 (84.8, 96.7); 32 | 0/31 (0.0);<br>100.0 (100.0, 100.0); 5 | 4/31 (12.9);<br>84.4 (63.7, 93.9); 6 |
|  | Day 28: deaths n (%);<br>KM survival (95% CI); censored | 8/109 (7.3);<br>88.6 (78.3, 94.2); 45 | 11/111 (9.9);<br>86.4 (76.7, 92.2); 34 | 0/31 (0.0);<br>100.0 (100.0, 100.0); 5 | 5/31 (16.1);<br>80.4 (59.1, 91.4); 7 |
|  | Day 90: deaths n (%);<br>KM survival (95% CI); censored | 13/109 (11.9);<br>80.7 (68.9, 88.4); 45 | 16/111 (14.4);<br>79.8 (69.1, 87.1); 35 | 0/31 (0.0);<br>100.0 (100.0, 100.0); 5 | 8/31 (25.8);<br>67.7 (45.6, 82.4); 7 |

\*Relaxed clinical cure omitted the repeat radiological-imaging requirement. †Length of stay was analysed as a time-to-event outcome; medians are reported as means are biased towards zero because the latest observation is censored. ‡Quantity cannot be estimated due to censoring of later observations. §Participants may have received antibiotics from more than one UK AWaRe class; class rows are not mutually exclusive. AWaRe=Access, Watch, Reserve; CT=computed tomography; CXR=chest x-ray; DDD=defined daily doses; FAPP=BIOFIRE FILMARRAY Pneumonia plus Panel; HAP=hospital-acquired pneumonia; IQR=interquartile range; KM=Kaplan-Meier; VAS=visual analogue scale. Censoring counts are cumulative to each time point and separate from participants classified as surviving at that time. Participants contributed follow-up until censoring; the SAP pre-specified censoring at withdrawal of consent. Death percentages use the randomised-group denominator and are not Kaplan–Meier estimates.

**Table 4:**
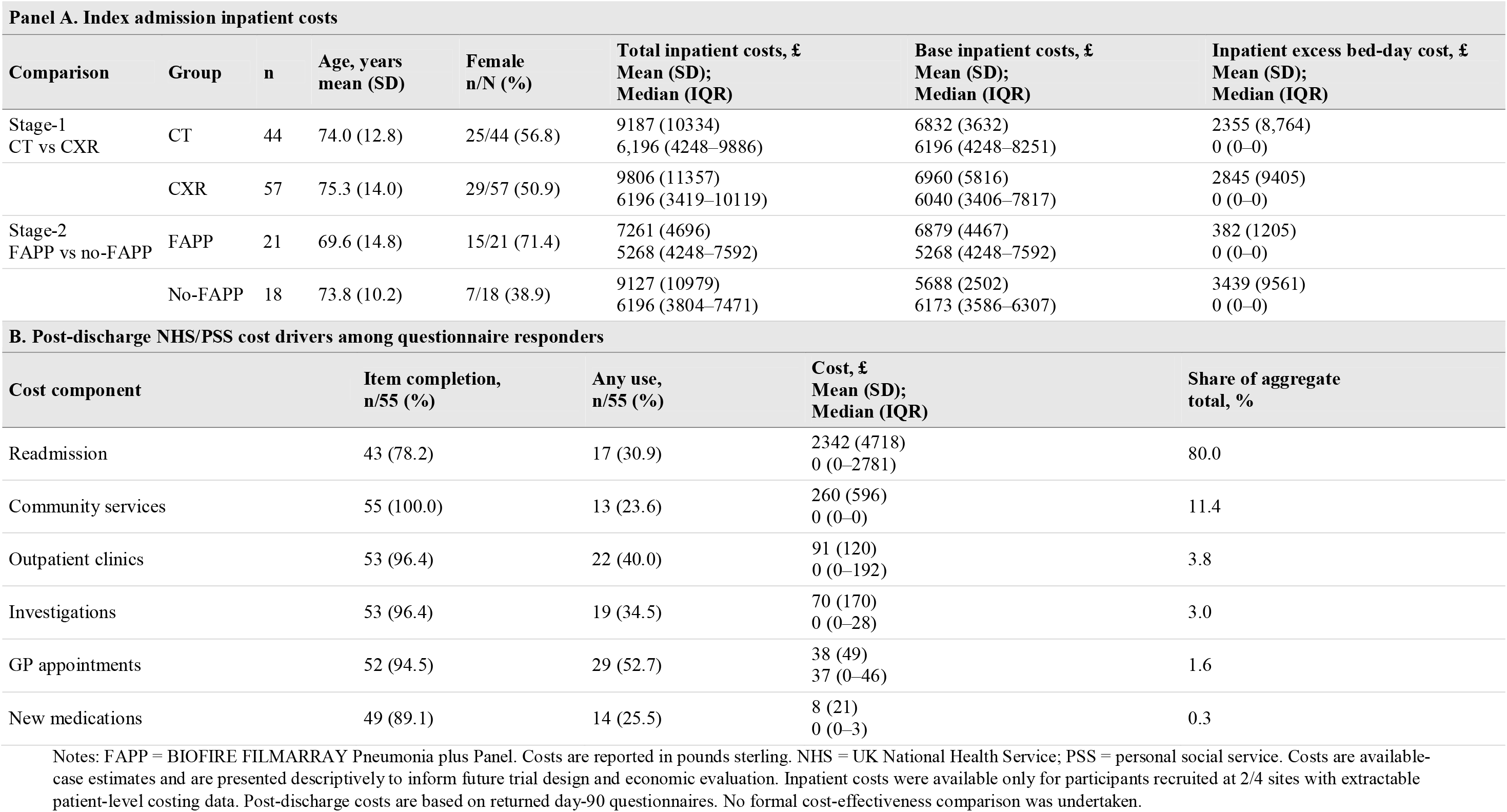
Costing feasibility: available-case inpatient costs, and post-discharge cost drivers.

| Panel A. Index admission inpatient costs |  |  |  |  |  |  |  |
|---|---|---|---|---|---|---|---|
| Comparison | Group | n | Age, years<br>mean (SD) | Female<br>n/N (%) | Total inpatient costs, £<br>Mean (SD);<br>Median (IQR) | Base inpatient costs, £<br>Mean (SD);<br>Median (IQR) | Inpatient excess bed-day cost, £<br>Mean (SD);<br>Median (IQR) |
| Stage-1<br>CT vs CXR | CT | 44 | 74.0 (12.8) | 25/44 (56.8) | 9187 (10334)<br>6,196 (4248–9886) | 6832 (3632)<br>6196 (4248–8251) | 2355 (8,764)<br>0 (0–0) |
|  | CXR | 57 | 75.3 (14.0) | 29/57 (50.9) | 9806 (11357)<br>6196 (3419–10119) | 6960 (5816)<br>6040 (3406–7817) | 2845 (9405)<br>0 (0–0) |
| Stage-2<br>FAPP vs no-FAPP | FAPP | 21 | 69.6 (14.8) | 15/21 (71.4) | 7261 (4696)<br>5268 (4248–7592) | 6879 (4467)<br>5268 (4248–7592) | 382 (1205)<br>0 (0–0) |
|  | No-FAPP | 18 | 73.8 (10.2) | 7/18 (38.9) | 9127 (10979)<br>6196 (3804–7471) | 5688 (2502)<br>6173 (3586–6307) | 3439 (9561)<br>0 (0–0) |
| **B. Post-discharge NHS/PSS cost drivers among questionnaire responders** |  |  |  |  |  |  |  |
| **Cost component** | **Item completion,<br>n/55 (%)** | **Any use,<br>n/55 (%)** | **Cost, £<br>Mean (SD);<br>Median (IQR)** | **Share of aggregate<br>total, %** |  |  |  |
| Readmission | 43 (78.2) | 17 (30.9) | 2342 (4718)<br>0 (0–2781) | 80.0 |  |  |  |
| Community services | 55 (100.0) | 13 (23.6) | 260 (596)<br>0 (0–0) | 11.4 |  |  |  |
| Outpatient clinics | 53 (96.4) | 22 (40.0) | 91 (120)<br>0 (0–192) | 3.8 |  |  |  |
| Investigations | 53 (96.4) | 19 (34.5) | 70 (170)<br>0 (0–28) | 3.0 |  |  |  |
| GP appointments | 52 (94.5) | 29 (52.7) | 38 (49)<br>37 (0–46) | 1.6 |  |  |  |
| New medications | 49 (89.1) | 14 (25.5) | 8 (21)<br>0 (0–3) | 0.3 |  |  |  |

Notes: FAPP = BIOFIRE FILMARRAY Pneumonia plus Panel. Costs are reported in pounds sterling. NHS = UK National Health Service; PSS = personal social service. Costs are available-case estimates and are presented descriptively to inform future trial design and economic evaluation. Inpatient costs were available only for participants recruited at 2/4 sites with extractable patient-level costing data. Post-discharge costs are based on returned day-90 questionnaires. No formal cost-effectiveness comparison was undertaken.

Mean antibiotic DDDs were 4.0 (SD 7.2) with CT, 4.8 (9.1) with CXR, 7.1 (9.4) with FAPP and 5.4 (7.9) without FAPP; missingness was substantial. DDD data were missing for 21/109 CT, 10/111 CXR, 7/31 FAPP and 8/31 no-FAPP participants (table 3). UK-AWaRe distributions were broadly similar at stage-1. Reserve use, exclusively meropenem, occurred in 6/109 (5.5%) CT and 9/111 (8.1%) CXR participants; at stage-2, it occurred in 8/31 (25.8%) FAPP participants and none without FAPP.

EQ-5D visual analogue scale (VAS) estimates were limited by incomplete follow-up. Mean baseline-to-day-90 changes were 7.7 points with CT and 5.4 with CXR; corresponding changes were 25.9 with FAPP and 15.7 without FAPP. Between-arm differences in VAS change, estimated using linear regression adjusted for baseline VAS score, were −0.9 points (95% CI −16.6 to 14.8) for CT versus CXR and 17.7 (−6.0 to 41.4) for FAPP versus no-FAPP.

Median post-HAP length of stay was 19 days (IQR 8-52) with CT and 13 (6-42) with CXR; corresponding stage-2 values were 9 (7-16) and 17 (9-not estimable). Long stays resulted in censoring.

By day 90, 29/220 stage-1 participants had died, with 80 censored (predominantly due to withdrawal of consent). Kaplan-Meier survival was 80.7% (95% CI 68.9-88.4) with CT and 79.8% (69.1-87.1) with CXR. No deaths were observed in the FAPP arm, with 5/31 censored, versus 8/31 deaths and 7/31 censored without FAPP. In the no-FAPP arm, four deaths were recorded by day 14 and five by day 28. Reported causes and timing of deaths are provided in SAR table 7-2.

### Costing feasibility

Inpatient costs were available for 101/151 participants at two sites; post-discharge NHS/personal social services data were available for 55/110 expected day-90 questionnaires (table 4). Available-case mean inpatient costs per participant were £9,187 with CT and £9,806 with CXR. In the smaller stage-2 subset, mean inpatient costs were higher without FAPP (£9,127 versus £7,261 with FAPP), largely reflecting excess bed days. Readmissions and community services accounted for 80.0% and 11.4% of aggregate recorded post-discharge costs, respectively.

## Discussion

HAP-FAST demonstrated successful recruitment to a sequential diagnostic trial in suspected nv-HAP but identified why a definitive trial should not simply enlarge this protocol. When managing suspected nv-HAP imaging and microbiology address linked decisions: whether nv-HAP explains a patient’s deterioration and which antibiotic should be chosen to target the responsible pathogen. The SMART design followed this sequence, while embedded qualitative and costing evidence explained implementation constraints that recruitment totals alone could obscure.^17, 18^ The central finding is therefore conditional feasibility, not evidence of clinical or economic effectiveness.

The recruitment target was reached early. Radiological tests performed at stage-1 successfully excluded a diagnosis of nv-HAP in 30% of participants thus after further excluding those who could not produce sputum, 28% of participants randomized at stage-1 entered stage-2. This proportion is essential for sample-size planning but limits generalisability of microbiological comparisons to the 40% of sputum-producing patients with radiologically confirmed nv-HAP. The qualitative finding of conditional acceptance helps distinguish willingness to enter an urgent diagnostic pathway from willingness to undertake repeated research procedures. Blood sampling and in-person follow-up discouraged continued participation; future designs should minimise non-essential sampling and use remote follow-up where feasible.

RWPC and clinician-led randomisation accommodated urgent treatment, ward dispersion and cognitive impairment without requiring specialist research staff on every ward continuously. Interviews supported RWPC but identified the need for staged explanations, later recap and consultee support.^28^ Acceptability should therefore be treated as an ongoing process rather than inferred from recruitment alone. The limited qualitative representation of people who did not consent further cautions against assuming universal acceptance. A definitive trial should distinguish agreement to routine-data follow-up from agreement to additional questionnaires, visits or samples wherever its consent arrangements permit.

Imaging findings illustrate the importance of separating delivery from documentation. Recorded CT receipt was incomplete, with missing forms contributing substantially to apparent non-delivery. Although recorded scans and reports were generally rapid, practitioners described scanner prioritisation, transfer logistics and tolerance as constraints. Future site-readiness assessments need explicit radiology agreements covering transfer, reporting and incidental findings, together with reliable delivery recording and allowance for crossover or non-receipt in design and analysis. The longer formal CXR reporting interval should not be equated with delayed interpretation, because managing clinicians interpreted CXR directly.

FAPP implementation was more fragile. Locating instruments near patients and training clinicians did not remove dependencies on sputum production, sample transfer, test ownership, result communication and antibiotic decision making. Qualitative accounts related recruitment decisions, sample losses and test non-completion to these practical steps. Future trials should implement systems for remote results access and antibiotic decision support, linking timely results to explicit stewardship review. Improved analytical speed alone does not ensure that diagnostic information reaches decision-makers or changes treatment.

Outcome measurement was a major feasibility result. The prespecified clinical-cure composite was effectively unusable because in clinical practice repeat imaging is rarely performed within days 7-10. Removing radiology post-hoc increased the proportion assessable but did not completely resolve missingness. Symptom data collected during days 0-10 may help characterise recovery and inform future assessment timepoints and thresholds, but this would require further evaluation rather than treating the exploratory definition as a validated endpoint. CAP-derived symptom and physiological criteria may not transfer readily to this population.^14, 15, 20, 22^ Cognitive impairment complicates self-report, while participants with an alternative diagnosis to pneumonia still require appropriate stage-1 outcomes. A future endpoint should therefore be diagnosis-agnostic, objective, routinely collected and suitable for an open-label trial; NEWS2 warrants evaluation.

Antibiotic stewardship outcomes may better match the mechanism of diagnostic intervention, but DDD alone does not distinguish appropriate escalation from avoidable exposure. The higher DDDs and ‘Reserve’ class antibiotic use seen with FAPP cannot establish benefit or harm: rapid microbiology results may prompt justified escalation as well as de-escalation. Related studies show these context-dependent prescribing effects and uncertainty around clinical endpoints.^9, 29^ A stewardship-focused outcome paired with safety or clinical response deserves evaluation.^30^ Outcome selection should precede definitive sample-size calculation, rather than relying on unstable effect estimates from this pilot.

Length of stay and quality-of-life measures provided planning information, but missingness, censoring and small stage-2 groups limit interpretation. Similar demographics in our comparator cohorts are reassuring, not proof of representativeness since HAP-FAST recruited when nv-HAP was suspected, whereas SUS and AQ capture coded and confirmed nv-HAP respectively. Apparent differences in outcomes across these sources may therefore reflect case definition as well as recruitment. Without screening denominators, recruitment may still depend on recognition timing, CT tolerance, clinician availability and sputum production. Mortality differences observed in these small, incompletely followed groups do not support effectiveness conclusions but are valuable for future trial planning.

The 50% post-discharge costing questionnaire return and available-case costing create potential selection biases, beyond item-level missingness so data inform future measurement rather than prove cost-effectiveness. Assistance with completion or routine-data extraction should be tested for acceptability and completeness, not assumed to solve missingness automatically.

The CAP-NEXT community-acquired pneumonia study also encountered incomplete delivery of low-dose CT and molecular testing, suggesting that diagnostic information must be integrated with workflow and prescribing decisions.^31^ That study stopped early during COVID-19 and did not collect reasons for non-adherence. HAP-FAST’s qualitative component addresses this explanatory gap for a different pneumonia population, linking observed failures to practical delivery conditions and participant experience. HAP-FAST adds nv-HAP-specific evidence by connecting quantitative delivery and retention measures with patient, consultee and practitioner accounts. Strengths include this integration, pragmatic sequential randomisation and evaluation of RWPC in a pneumonia trial outside critical care. Limitations include incomplete interventions and outcomes, available-case costing, absent screening logs and limited qualitative representation from non-consenters.

Progression is therefore conditional on redesign: enhanced RWPC, protected diagnostic pathways, enhanced sputum procedures, lower-burden follow-up, diagnosis-agnostic and stewardship-aligned outcomes, and robust economic data capture. A definitive trial should consider an internal pilot with prespecified stop-go criteria for recruitment, intervention delivery, outcome completeness and costing. Qualitative evaluation should remain integrated to explain whether refinements work, for whom and under which delivery conditions, informing implementation in clinical practice.

## Ethics Statement

The study was approved by Wales Research Ethics Committee 1, Health and Care Research Wales (HCRW) ref. 22/WA/0315

## Data sharing

De-identified data is available on request from LCTC by request to the corresponding author and following approval from the sponsor. De-identified qualitative data is available on request to the corresponding author and following approval from the sponsor.

## Supporting information

statistical analysis plan

Statistical Analysis Report

## Data Availability

De-identified data is available on request from Liverpool Clinical Trials Centre by request to the corresponding author and following approval from the sponsor, University of Liverpool. De-identified qualitative data is available on request to the corresponding author and following approval from the sponsor.

## Acknowledgements

The research team would also like to acknowledge the enthusiasm and support of all the clinicians, primarily resident grade doctors who randomized patients into HAP-FAST. DW would like to acknowledge the research mentorship provided by Professor Enitan Carrol, Professor Graham Devereux, Professor Richard Grieve and Professor John Norrie.

## Authors contributions

Conceptualization: DW

Funding acquisition: DW, ANN

Data curation: I.Powell, AJ, FS, FF, PE, ANG, FK, HC, TJ, CM, DW

Methodology: DW, AJ, FS, BY, SW, LT, SA, AAN, AH, JW

Project administration: SW, AT, DW, SJA, HS, RB, NS, LP, ST, FS, AC, J.Hare, J.Howells, JHS, KI, KE, BP, ET

Resources: AMC, AC, KE, NG, J.Hare, AH, J.Howells, JHS, KI, CM,

Investigation: DW, SJA, RB, HS, NS, EA, AMC, KC, AC, KE, NG, JHS, KI, KK, DM, CM, LP, ET, ST, SW, JW, FS

Supervision: DW, SJA, RB, HS, AT, BY, AJ, LT, AAN

Validation: I.Powell, AJ, FS and DW

Visualization: I.Powell and DW

Formal analysis: I.Powell, AJ, FS, FF, FK, HC, PE, ANG, TJ, DW Writing – original draft: DW

Writing – review & editing: All authors reviewed the manuscript and DW edited. All authors agreed the final manuscript.

---

**Supplementary Table 1:**
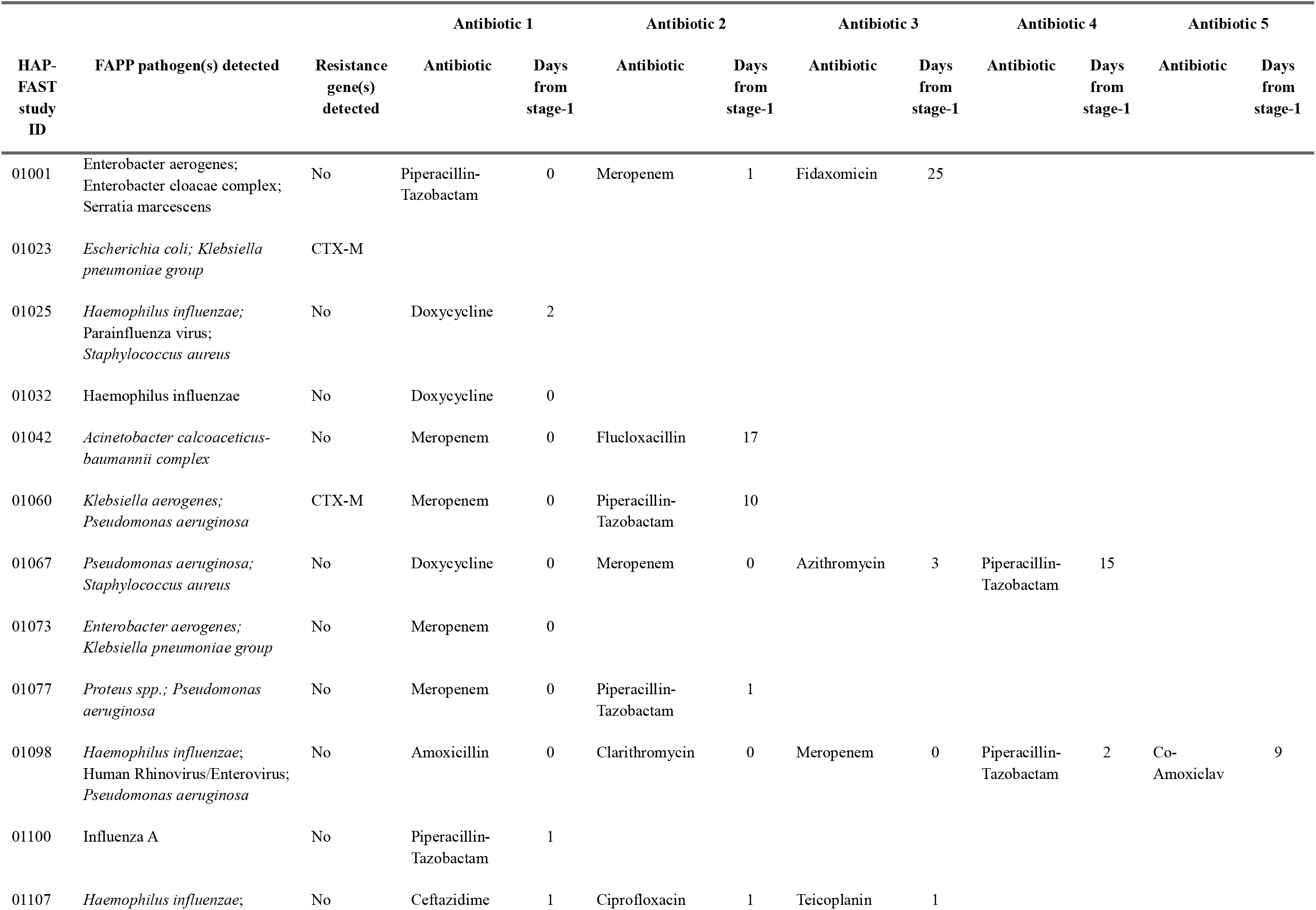
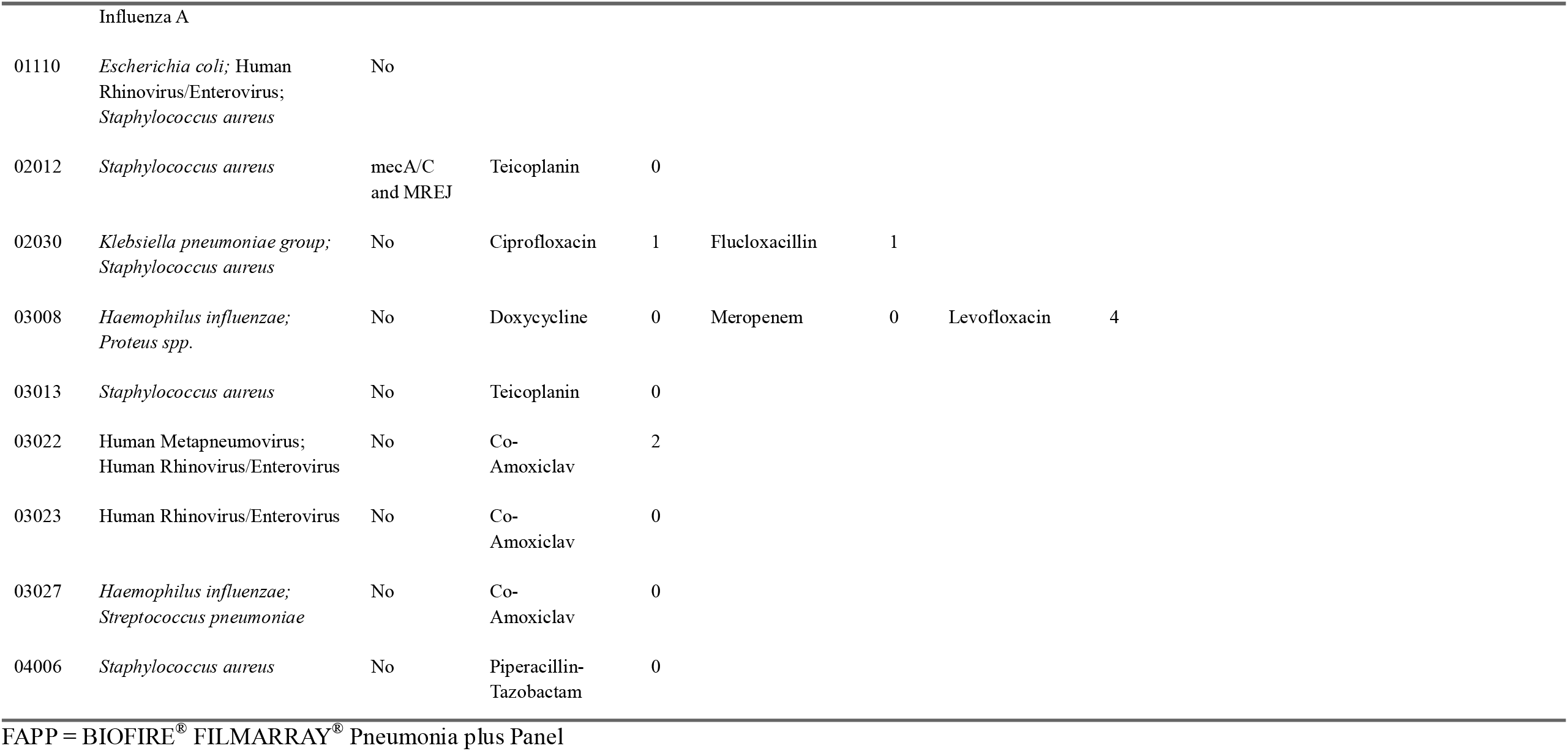
Participants with pathogen(s) detected by the FAPP, detected resistance genes, and antibiotics started on or after stage-1 randomisation.

|  |  |  | **Antibiotic 1** |  | **Antibiotic 2** |  | **Antibiotic 3** |  | **Antibiotic 4** |  | **Antibiotic 5** |  |
|---|---|---|---|---|---|---|---|---|---|---|---|---|
| **HAP-FAST study ID** | **FAPP pathogen(s) detected** | **Resistance gene(s) detected** | **Antibiotic** | **Days from stage-1** | **Antibiotic** | **Days from stage-1** | **Antibiotic** | **Days from stage-1** | **Antibiotic** | **Days from stage-1** | **Antibiotic** | **Days from stage-1** |
| 01001 | Enterobacter aerogenes; Enterobacter cloacae complex; Serratia marcescens | No | Piperacillin-Tazobactam | 0 | Meropenem | 1 | Fidaxomicin | 25 |  |  |  |  |
| 01023 | *Escherichia coli; Klebsiella pneumoniae group* | CTX-M |  |  |  |  |  |  |  |  |  |  |
| 01025 | *Haemophilus influenzae;* Parainfluenza virus; *Staphylococcus aureus* | No | Doxycycline | 2 |  |  |  |  |  |  |  |  |
| 01032 | Haemophilus influenzae | No | Doxycycline | 0 |  |  |  |  |  |  |  |  |
| 01042 | *Acinetobacter calcoaceticus-baumannii complex* | No | Meropenem | 0 | Flucloxacillin | 17 |  |  |  |  |  |  |
| 01060 | *Klebsiella aerogenes; Pseudomonas aeruginosa* | CTX-M | Meropenem | 0 | Piperacillin-Tazobactam | 10 |  |  |  |  |  |  |
| 01067 | *Pseudomonas aeruginosa; Staphylococcus aureus* | No | Doxycycline | 0 | Meropenem | 0 | Azithromycin | 3 | Piperacillin-Tazobactam | 15 |  |  |
| 01073 | *Enterobacter aerogenes; Klebsiella pneumoniae group* | No | Meropenem | 0 |  |  |  |  |  |  |  |  |
| 01077 | *Proteus spp.; Pseudomonas aeruginosa* | No | Meropenem | 0 | Piperacillin-Tazobactam | 1 |  |  |  |  |  |  |
| 01098 | *Haemophilus influenzae*; Human Rhinovirus/Enterovirus; *Pseudomonas aeruginosa* | No | Amoxicillin | 0 | Clarithromycin | 0 | Meropenem | 0 | Piperacillin-Tazobactam | 2 | Co-Amoxiclav | 9 |
| 01100 | Influenza A | No | Piperacillin-Tazobactam | 1 |  |  |  |  |  |  |  |  |
| 01107 | *Haemophilus influenzae*; Influenza A | No | Ceftazidime | 1 | Ciprofloxacin | 1 | Teicoplanin | 1 |  |  |  |  |
| 01110 | *Escherichia coli;* Human Rhinovirus/Enterovirus; *Staphylococcus aureus* | No |  |  |  |  |  |  |  |  |  |  |
| 02012 | *Staphylococcus aureus* | mecA/C and MREJ | Teicoplanin | 0 |  |  |  |  |  |  |  |  |
| 02030 | *Klebsiella pneumoniae group; Staphylococcus aureus* | No | Ciprofloxacin | 1 | Flucloxacillin | 1 |  |  |  |  |  |  |
| 03008 | *Haemophilus influenzae; Proteus spp.* | No | Doxycycline | 0 | Meropenem | 0 | Levofloxacin | 4 |  |  |  |  |
| 03013 | *Staphylococcus aureus* | No | Teicoplanin | 0 |  |  |  |  |  |  |  |  |
| 03022 | Human Metapneumovirus; Human Rhinovirus/Enterovirus | No | Co-Amoxiclav | 2 |  |  |  |  |  |  |  |  |
| 03023 | Human Rhinovirus/Enterovirus | No | Co-Amoxiclav | 0 |  |  |  |  |  |  |  |  |
| 03027 | *Haemophilus influenzae; Streptococcus pneumoniae* | No | Co-Amoxiclav | 0 |  |  |  |  |  |  |  |  |
| 04006 | *Staphylococcus aureus* | No | Piperacillin-Tazobactam | 0 |  |  |  |  |  |  |  |  |

FAPP = BIOFIRE<sup>®</sup> FILMARRAY<sup>®</sup> Pneumonia plus Panel

**Diagnostic strategies for suspected non-ventilator-associated hospital-acquired pneumonia: a pilot Sequential Multiple Assignment Randomised Trial with integrated qualitative and costing substudies**

**CONSORT 2025 main checklist**

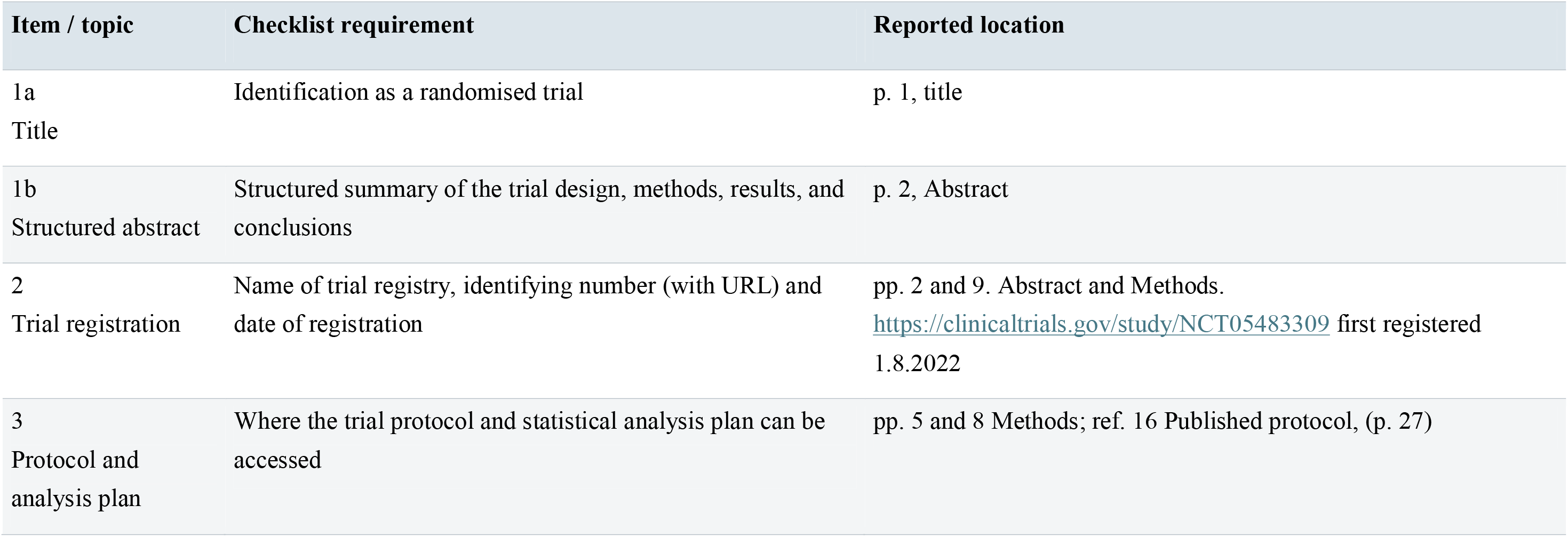
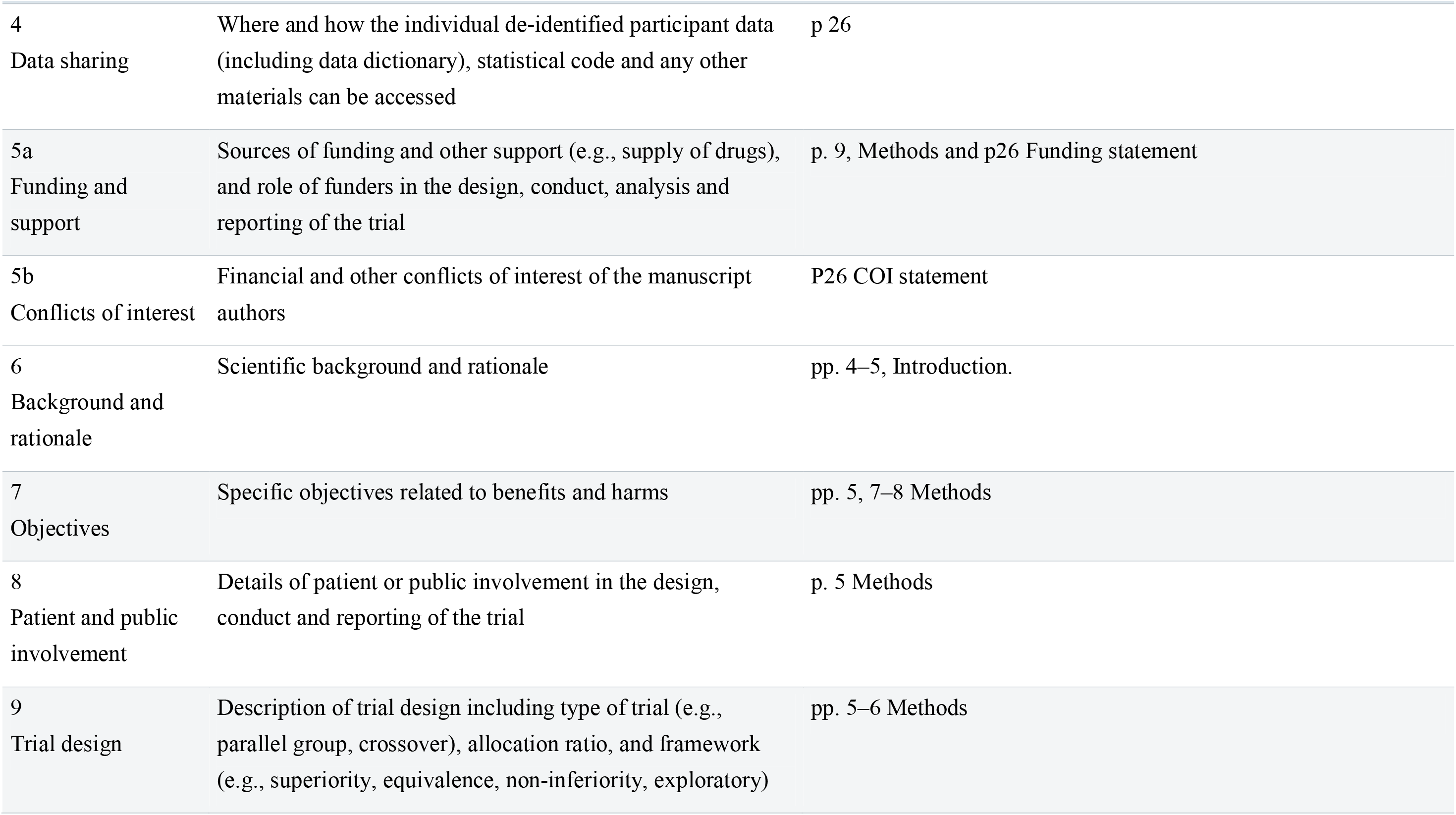
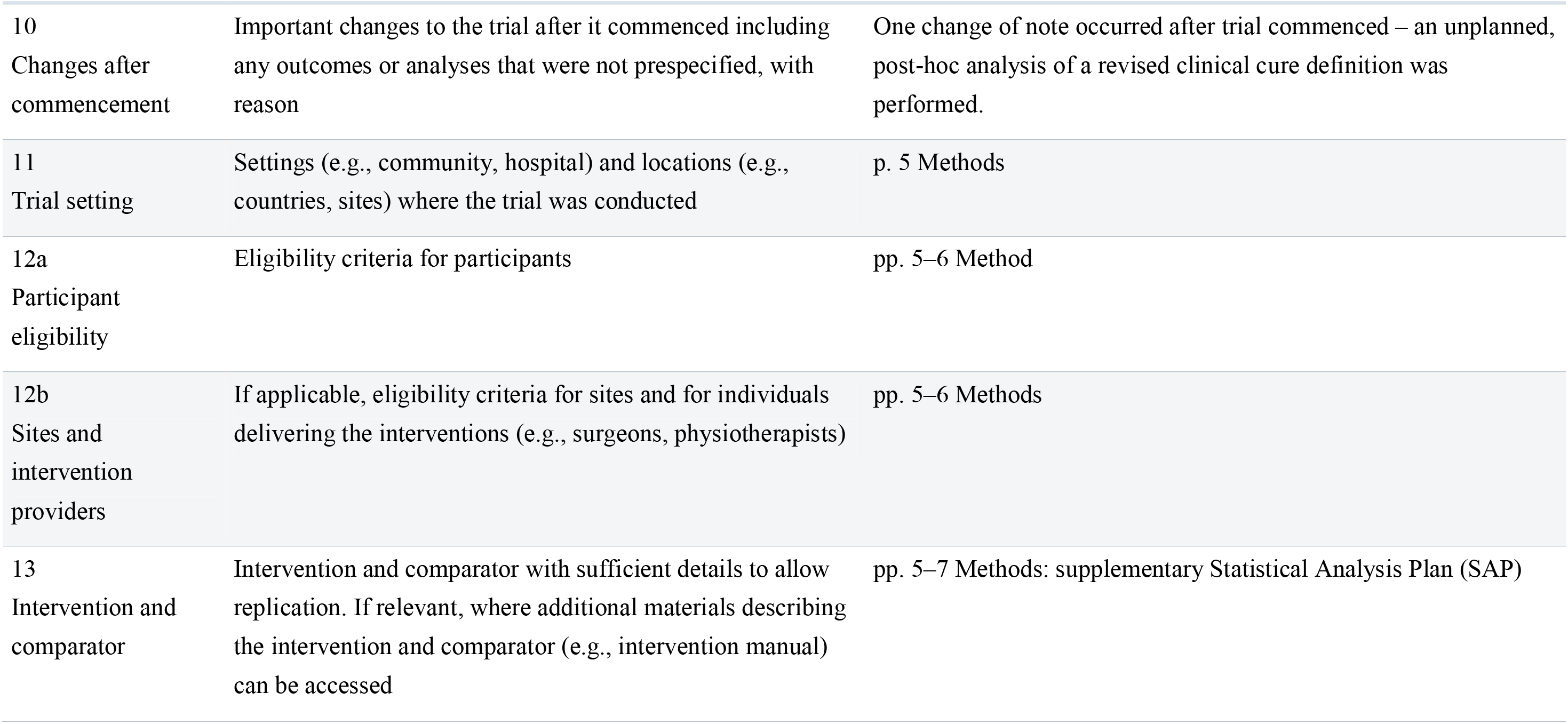
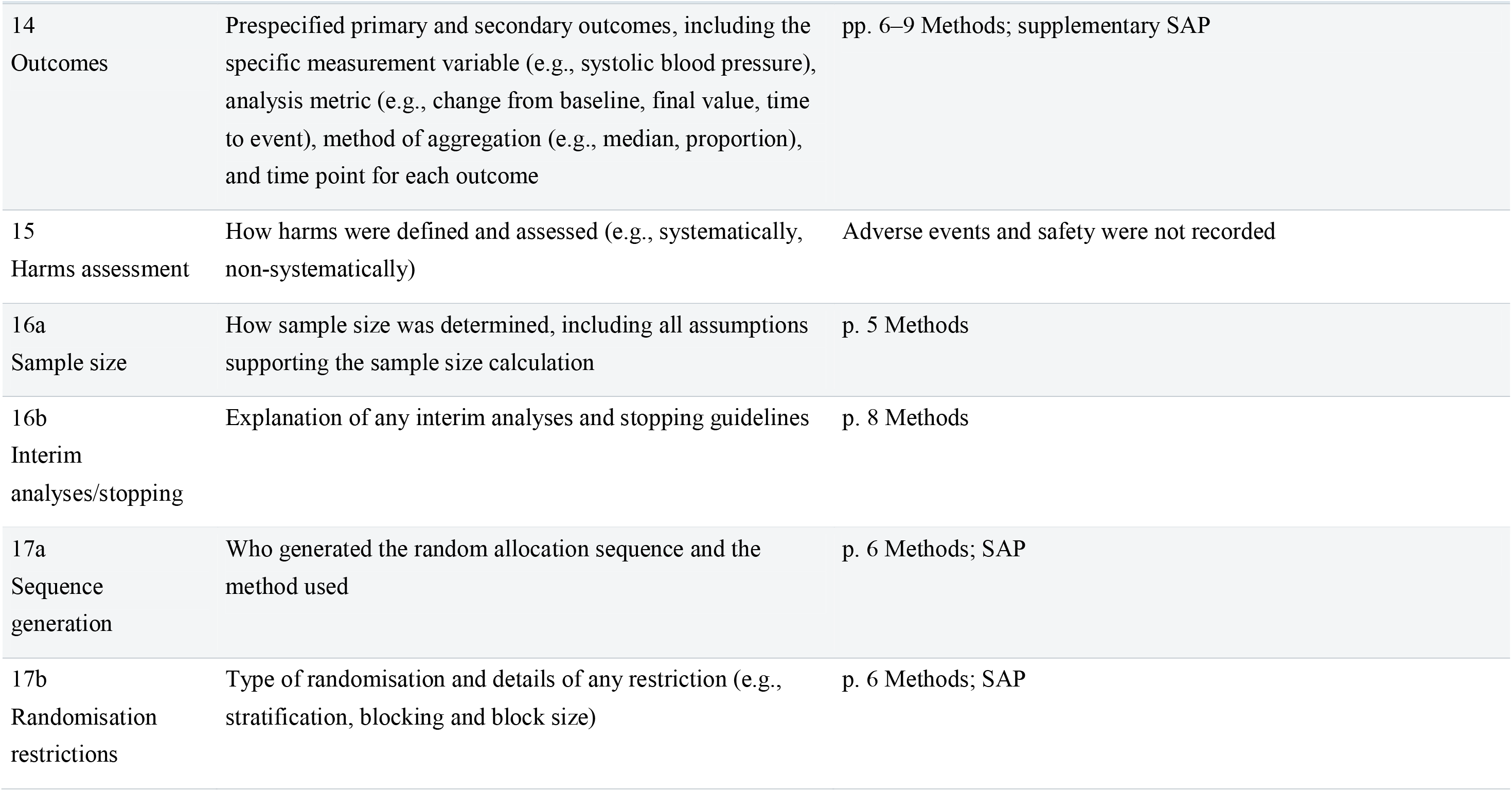
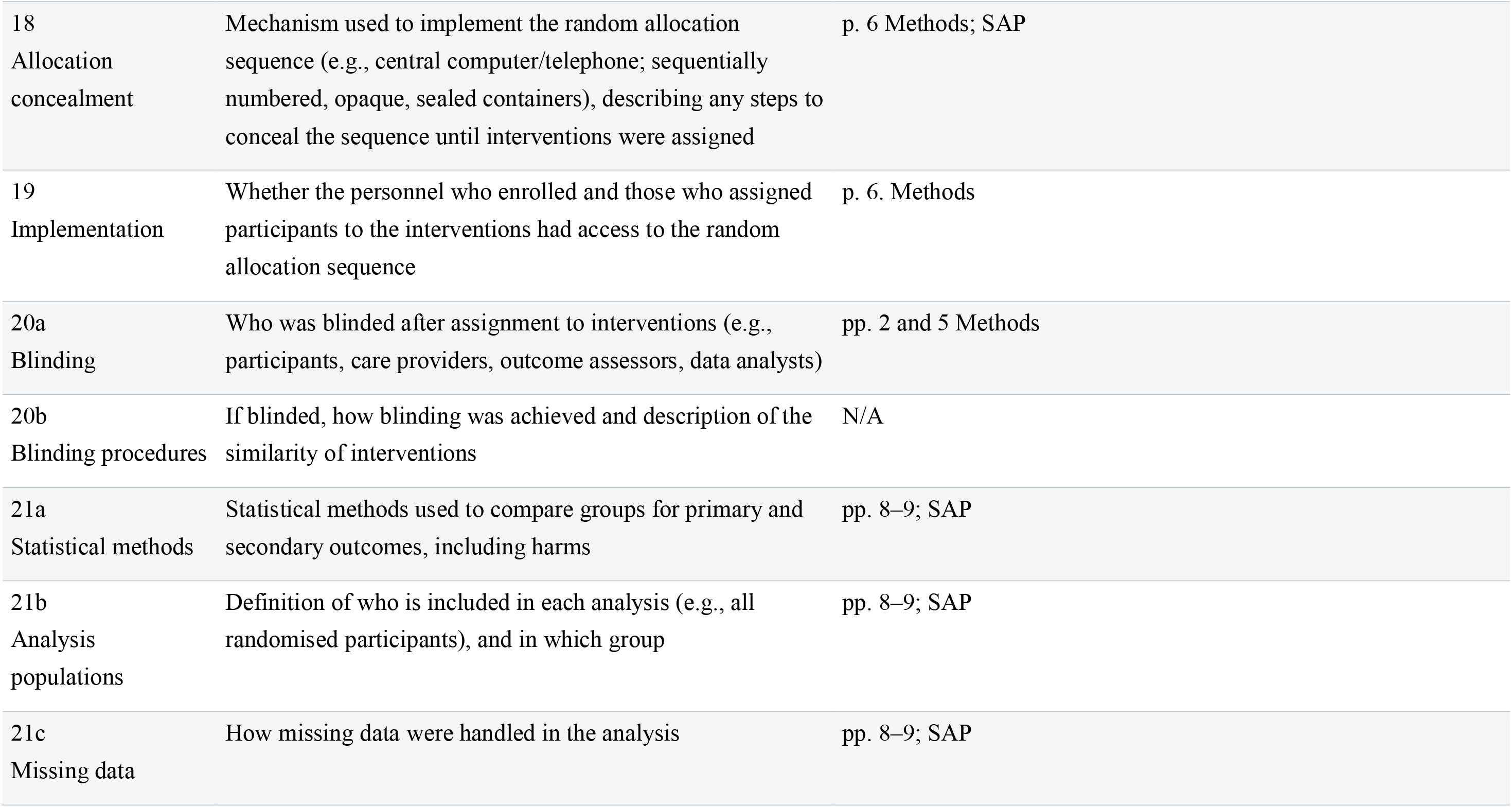
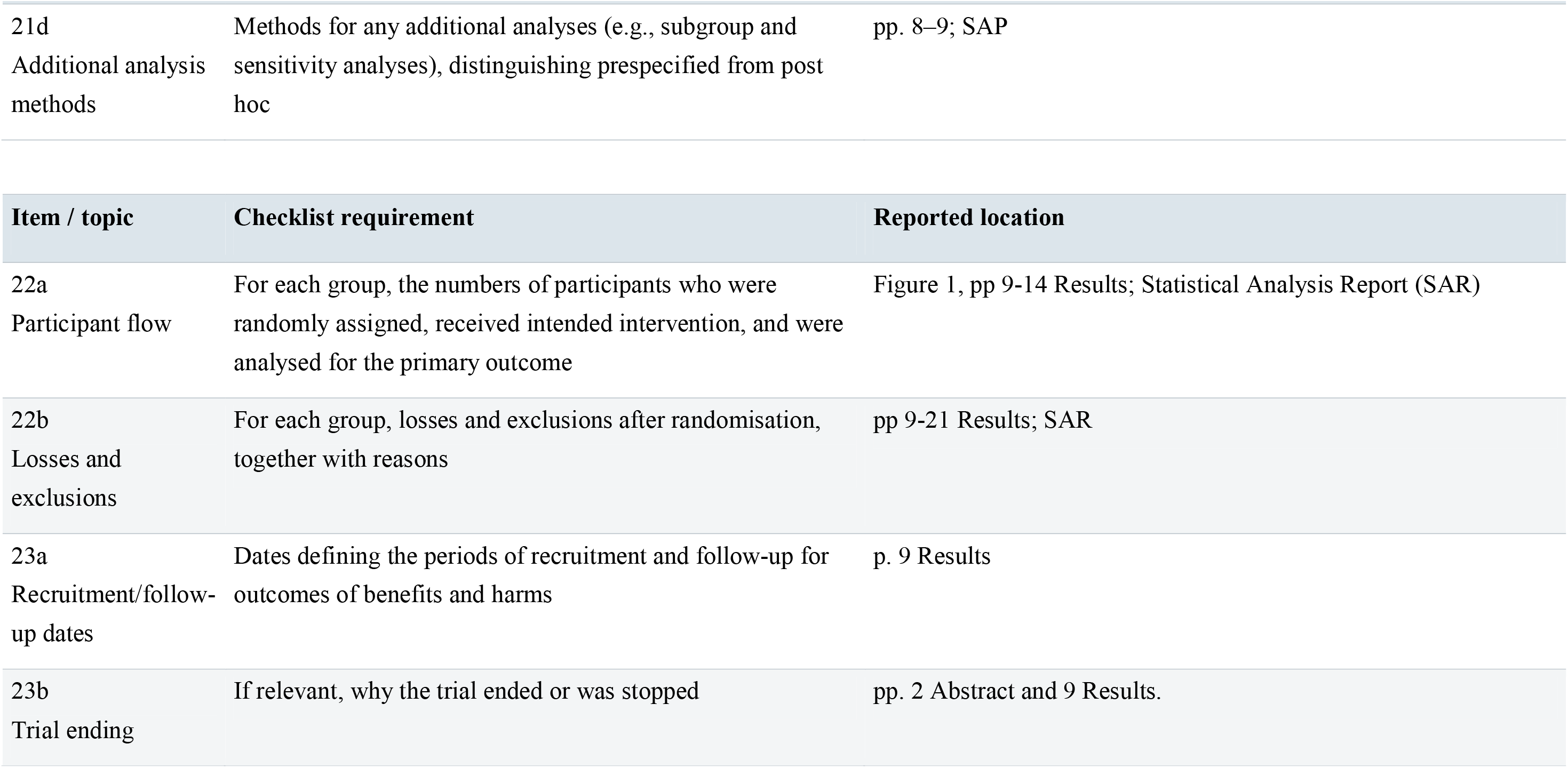
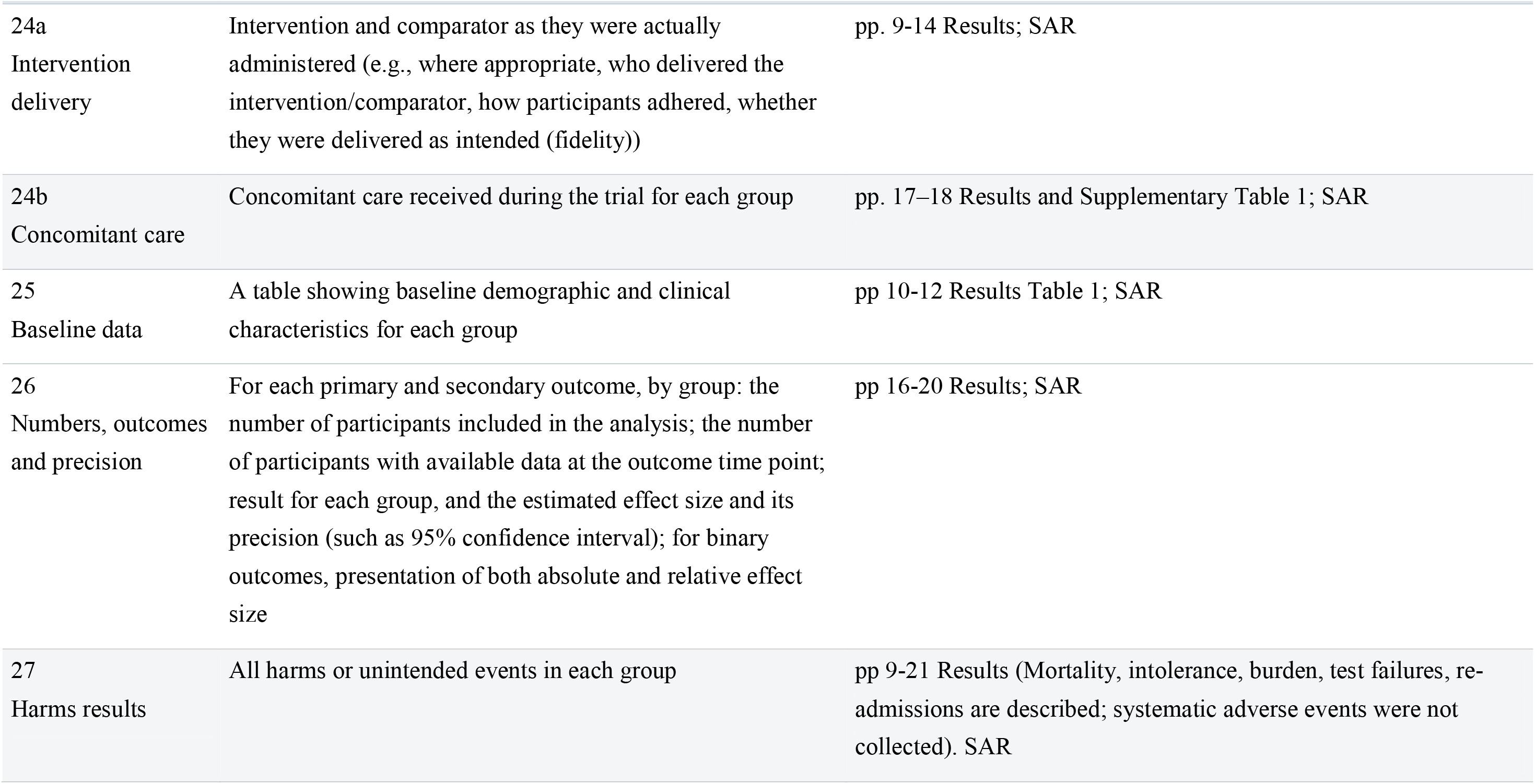
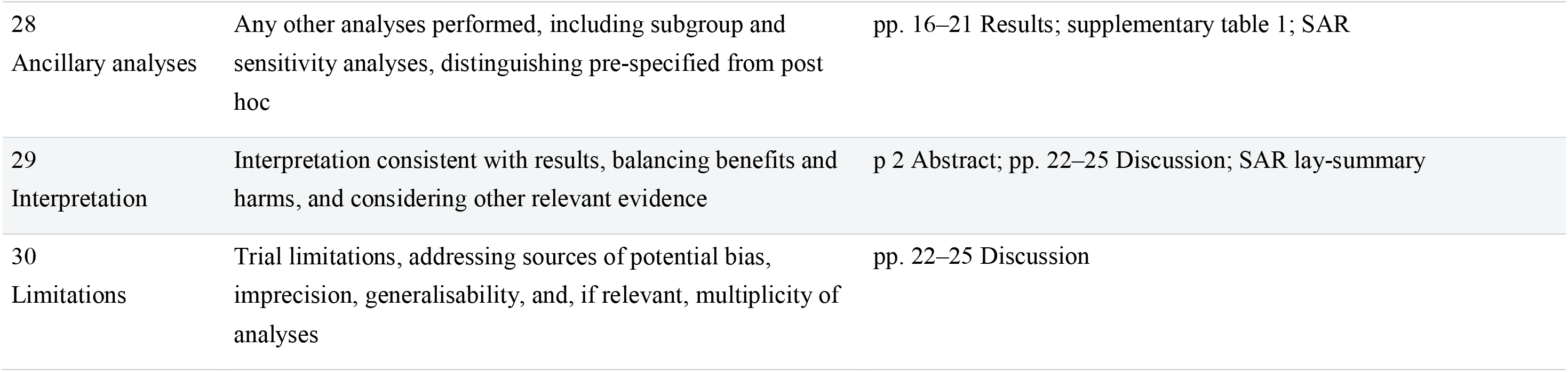

| **Item / topic** | **Checklist requirement** | **Reported location** |
|---|---|---|
| 1a<br>Title | Identification as a randomised trial | p. 1, title |
| 1b<br>Structured abstract | Structured summary of the trial design, methods, results, and conclusions | p. 2, Abstract |
| 2<br>Trial registration | Name of trial registry, identifying number (with URL) and date of registration | pp. 2 and 9. Abstract and Methods. https://clinicaltrials.gov/study/NCT05483309 first registered 1.8.2022 |
| 3<br>Protocol and analysis plan | Where the trial protocol and statistical analysis plan can be accessed | pp. 5 and 8 Methods; ref. 16 Published protocol, (p. 27) |
| 4<br>Data sharing | Where and how the individual de-identified participant data (including data dictionary), statistical code and any other materials can be accessed | p 26 |
| 5a<br>Funding and support | Sources of funding and other support (e.g., supply of drugs), and role of funders in the design, conduct, analysis and reporting of the trial | p. 9, Methods and p26 Funding statement |
| 5b<br>Conflicts of interest | Financial and other conflicts of interest of the manuscript authors | P26 COI statement |
| 6<br>Background and rationale | Scientific background and rationale | pp. 4–5, Introduction. |
| 7<br>Objectives | Specific objectives related to benefits and harms | pp. 5, 7–8 Methods |
| 8<br>Patient and public involvement | Details of patient or public involvement in the design, conduct and reporting of the trial | p. 5 Methods |
| 9<br>Trial design | Description of trial design including type of trial (e.g., parallel group, crossover), allocation ratio, and framework (e.g., superiority, equivalence, non-inferiority, exploratory) | pp. 5–6 Methods |
| 10<br>Changes after commencement | Important changes to the trial after it commenced including any outcomes or analyses that were not prespecified, with reason | One change of note occurred after trial commenced – an unplanned, post-hoc analysis of a revised clinical cure definition was performed. |
| 11<br>Trial setting | Settings (e.g., community, hospital) and locations (e.g., countries, sites) where the trial was conducted | p. 5 Methods |
| 12a<br>Participant eligibility | Eligibility criteria for participants | pp. 5–6 Method |
| 12b<br>Sites and intervention providers | If applicable, eligibility criteria for sites and for individuals delivering the interventions (e.g., surgeons, physiotherapists) | pp. 5–6 Methods |
| 13<br>Intervention and comparator | Intervention and comparator with sufficient details to allow replication. If relevant, where additional materials describing the intervention and comparator (e.g., intervention manual) can be accessed | pp. 5–7 Methods: supplementary Statistical Analysis Plan (SAP) |
| 14<br>Outcomes | Prespecified primary and secondary outcomes, including the specific measurement variable (e.g., systolic blood pressure), analysis metric (e.g., change from baseline, final value, time to event), method of aggregation (e.g., median, proportion), and time point for each outcome | pp. 6–9 Methods; supplementary SAP |
| 15<br>Harms assessment | How harms were defined and assessed (e.g., systematically, non-systematically) | Adverse events and safety were not recorded |
| 16a<br>Sample size | How sample size was determined, including all assumptions supporting the sample size calculation | p. 5 Methods |
| 16b<br>Interim analyses/stopping | Explanation of any interim analyses and stopping guidelines | p. 8 Methods |
| 17a<br>Sequence generation | Who generated the random allocation sequence and the method used | p. 6 Methods; SAP |
| 17b<br>Randomisation restrictions | Type of randomisation and details of any restriction (e.g., stratification, blocking and block size) | p. 6 Methods; SAP |
| 18<br>Allocation concealment | Mechanism used to implement the random allocation sequence (e.g., central computer/telephone; sequentially numbered, opaque, sealed containers), describing any steps to conceal the sequence until interventions were assigned | p. 6 Methods; SAP |
| 19<br>Implementation | Whether the personnel who enrolled and those who assigned participants to the interventions had access to the random allocation sequence | p. 6. Methods |
| 20a<br>Blinding | Who was blinded after assignment to interventions (e.g., participants, care providers, outcome assessors, data analysts) | pp. 2 and 5 Methods |
| 20b<br>Blinding procedures | If blinded, how blinding was achieved and description of the similarity of interventions | N/A |
| 21a<br>Statistical methods | Statistical methods used to compare groups for primary and secondary outcomes, including harms | pp. 8–9; SAP |
| 21b<br>Analysis populations | Definition of who is included in each analysis (e.g., all randomised participants), and in which group | pp. 8–9; SAP |
| 21c<br>Missing data | How missing data were handled in the analysis | pp. 8–9; SAP |
| 21d<br>Additional analysis methods | Methods for any additional analyses (e.g., subgroup and sensitivity analyses), distinguishing prespecified from post hoc | pp. 8–9; SAP |
| **Item / topic** | **Checklist requirement** | **Reported location** |
| 22a<br>Participant flow | For each group, the numbers of participants who were randomly assigned, received intended intervention, and were analysed for the primary outcome | Figure 1, pp 9-14 Results; Statistical Analysis Report (SAR) |
| 22b<br>Losses and exclusions | For each group, losses and exclusions after randomisation, together with reasons | pp 9-21 Results; SAR |
| 23a<br>Recruitment/follow-up dates | Dates defining the periods of recruitment and follow-up for outcomes of benefits and harms | p. 9 Results |
| 23b<br>Trial ending | If relevant, why the trial ended or was stopped | pp. 2 Abstract and 9 Results. |
| 24a<br>Intervention delivery | Intervention and comparator as they were actually administered (e.g., where appropriate, who delivered the intervention/comparator, how participants adhered, whether they were delivered as intended (fidelity)) | pp. 9-14 Results; SAR |
| 24b<br>Concomitant care | Concomitant care received during the trial for each group | pp. 17–18 Results and Supplementary Table 1; SAR |
| 25<br>Baseline data | A table showing baseline demographic and clinical characteristics for each group | pp 10-12 Results Table 1; SAR |
| 26<br>Numbers, outcomes and precision | For each primary and secondary outcome, by group: the number of participants included in the analysis; the number of participants with available data at the outcome time point; result for each group, and the estimated effect size and its precision (such as 95% confidence interval); for binary outcomes, presentation of both absolute and relative effect size | pp 16-20 Results; SAR |
| 27<br>Harms results | All harms or unintended events in each group | pp 9-21 Results (Mortality, intolerance, burden, test failures, re-admissions are described; systematic adverse events were not collected). SAR |
| 28<br>Ancillary analyses | Any other analyses performed, including subgroup and sensitivity analyses, distinguishing pre-specified from post hoc | pp. 16–21 Results; supplementary table 1; SAR |
| 29<br>Interpretation | Interpretation consistent with results, balancing benefits and harms, and considering other relevant evidence | p 2 Abstract; pp. 22–25 Discussion; SAR lay-summary |
| 30<br>Limitations | Trial limitations, addressing sources of potential bias, imprecision, generalisability, and, if relevant, multiplicity of analyses | pp. 22–25 Discussion |

**CONSORT pilot and feasibility extension**

| Item / topic | Checklist requirement | Reported location |
|---|---|---|
| 1a<br>Title | Identification as a pilot or feasibility randomised trial in the title | p. 1, title: explicitly a pilot randomised SMART; p2 Abstract; p 5 Methods |
| 1b<br>Abstract | Structured summary of pilot trial design, methods, results, and conclusions (for specific guidance see CONSORT abstract extension for pilot trials) | p. 2 Abstract, pp5-10 Methods; SAP; ref. 16 Published protocol |
| 2a<br>Background and objectives | Scientific background and explanation of rationale for future definitive trial, and reasons for randomised pilot trial | pp. 4–5, Introduction; ref. 16 Published protocol |
| 2b<br>Background and objectives | Specific objectives or research questions for pilot trial | p. 4-5 Introduction; SAP; ref. 16 Published protocol |
| 3a<br>Trial design | Description of pilot trial design (such as parallel, factorial) including allocation ratio | pp. 5–6 Methods; SAP; ref. 16 Published protocol |
| 3b<br>Trial design | Important changes to methods after pilot trial commencement (such as eligibility criteria), with reasons | N/A no significant changes to methods |
| 4a<br>Participants | Eligibility criteria for participants | pp. 5–6 Methods; ref. 16 Published protocol |
| 4b<br>Participants | Settings and locations where the data were collected | p. 5 Methods |
| 4c<br>Identification/consent | How participants were identified and consented | pp. 5–7 Methods; ref. 16 Published protocol |
| 5<br>Interventions | The interventions for each group with sufficient details to allow replication, including how and when they were actually administered | pp. 5–7 Methods and Figure 1 and Supplementary Table 1; SAP; ref. 16 Published protocol |
| 6a<br>Outcomes | Completely defined prespecified assessments or measurements to address each pilot trial objective specified in 2b, including how and when they were assessed | pp. 6–9 Methods; SAP; ref. 16 Published protocol |
| 6b<br>Outcomes | Any changes to pilot trial assessments or measurements after the pilot trial commenced, with reasons | pp. 16–17 Results – post hoc “relaxed” clinical cure assessment; SAR |
| 6c<br>Progression criteria | If applicable, prespecified criteria used to judge whether, or how, to proceed with future definitive trial | p. 8 Methods; ref. 16 Published protocol |
| 7a<br>Sample size | Rationale for numbers in the pilot trial | p. 5 Methods and SAP gives target 220, 12-month plan and audit-based eligibility recruitment assumption; ref. 16 Published protocol |
| 7b<br>Sample size | When applicable, explanation of any interim analyses and stopping guidelines | p. 8 Methods. |
| 8a<br>Sequence generation | Method used to generate the random allocation sequence | p. 6 Methods; SAP; ref. 16 Published protocol |
| 8b<br>Sequence generation | Type of randomisation(s); details of any restriction (such as blocking and block size) | p. 6 Methods; SAP; ref. 16 Published protocol |
| 9<br>Allocation concealment mechanism | Mechanism used to implement the random allocation sequence (such as sequentially numbered containers), describing any steps taken to conceal the sequence until interventions were assigned | p. 6 Methods; SAP; ref. 16 Published protocol |
| 10<br>Implementation | Who generated the random allocation sequence, who enrolled participants, and who assigned participants to interventions | p. 6 Methods; SAP; ref. 16 Published protocol |
| 11a<br>Blinding | If done, who was blinded after assignment to interventions (for example, participants, care providers, those assessing outcomes) and how | p. 6 Methods; SAP; ref. 16 Published protocol |
| 11b<br>Blinding | If relevant, description of the similarity of interventions | N/A open label |
| 12<br>Statistical methods | Methods used to address each pilot trial objective whether qualitative or quantitative | pp. 7–9 Methods; SAP; ref. 16 Published protocol |
| 13a<br>Participant flow (a diagram is strongly recommended) | For each group, the numbers of participants who were approached and/or assessed for eligibility, randomly assigned, received intended treatment, and were assessed for each objective | p. 9, Results and Figure 1; SAR |
| 13b<br>Participant flow (a diagram is strongly recommended) | For each group, losses and exclusions after randomisation, together with reasons | p. 9, Results and Figure 1; SAR |
| 14a<br>Recruitment | Dates defining the periods of recruitment and follow-up | p. 9 Results and SAR. |
| 14b<br>Recruitment | Why the pilot trial ended or was stopped | pp. 2 Abstract and 9 Results |
| 15<br>Baseline data | A table showing baseline demographic and clinical characteristics for each group | Pp 10-12 Table 1; SAR |
| 16<br>Numbers analysed | For each objective, number of participants (denominator) included in each analysis. If relevant, these numbers should be by randomised group | Pp 13-21 Results and supplementary table 1; SAR |
| 17<br>Outcomes and estimation | For each objective, results including expressions of uncertainty (such as 95% confidence interval) for any estimates. If relevant, these results should be by randomised group | pp. 13–21 Results and supplementary table 1; SAR |
| 18<br>Ancillary analyses | Results of any other analyses performed that could be used to inform the future definitive trial | pp. 16–21 Results, 23–25 Discussion; SAR |
| 19<br>Harms | All important harms or unintended effects in each group (for specific guidance see CONSORT for harms) | pp. 14–19 Results; SAR.<br>Burden, intolerance and mortality are described; no systematic safety-event collection. |
| 19a<br>Unintended consequences | If relevant, other important unintended consequences | pp. 14–16 Results and pp. 22–25 Discussion; SAR |
| 20<br>Limitations | Pilot trial limitations, addressing sources of potential bias and remaining uncertainty about feasibility | pp. 22–25 Discussion; SAR lay summary |
| 21<br>Generalisability | Generalisability (applicability) of pilot trial methods and findings to future definitive trial and other studies | pp. 22–25 Discussion |
| 22<br>Interpretation | Interpretation consistent with pilot trial objectives and findings, balancing potential benefits and harms, and considering other relevant evidence | pp. 22–25 Discussion; SAR with lay summary. |
| 22a<br>Progression implications | Implications for progression from pilot to future definitive trial, including any proposed amendments | pp. 22–25 Discussion |
| 23<br>Registration | Registration number for pilot trial and name of trial registry | pp. 2 Abstract and 9 Methods NCT05483309 |
| 24<br>Protocol | Where the pilot trial protocol can be accessed, if available | p. 5, protocol citation; reference 16, p. 27, identifies Shafiqa et al., BMJ Open 2024;14:e088490, DOI 10.1136/bmjopen-2024-088490. |
| 25<br>Funding | Sources of funding and other support (such as supply of drugs), role of funders | p. 9. NIHR award, bioMérieux equipment/panel support and role in design, conduct, analysis stated. |
| 26<br>Ethics | Ethical approval or approval by research review committee, confirmed with reference number | p. 9. Methods NHS REC/HRA approval reference 22/WA/0315; sponsor and independent steering oversight also stated. |

**CONSORT Abstract checklist**

| **Item / topic** | **Checklist requirement** | **Location** |
|---|---|---|
| 1<br>Title | Identification of study as randomised pilot or feasibility trial | p. 1, title |
| 2<br>Authors * | Contact details for the corresponding author | Not applicable. Conference-only checklist item. |
| 3<br>Trial design | Description of pilot trial design (eg, parallel, cluster) | p. 2, Methods |
| 4<br>Participants | Eligibility criteria for participants and the settings where the pilot trial was conducted | p. 2, Methods |
| 5<br>Interventions | Interventions intended for each group | p. 2, Methods |
| 6<br>Objective | Specific objectives of the pilot trial | p. 2, Background |
| 7<br>Outcome | Prespecified assessment or measurement to address the pilot trial objectives** | p. 2, Methods |
| 8<br>Randomization | How participants were allocated to interventions | p. 2, Methods |
| 9<br>Blinding (masking) | Whether or not participants, care givers, and those assessing the outcomes were blinded to group assignment | p. 2, Methods |
| **Item / topic** | **Checklist requirement** | **Reported location and current status** |
| 10<br>Numbers randomized | Number of participants screened and randomised to each group for the pilot trial objectives** | p. 2, Results |
| 11<br>Recruitment | Trial status† | Not applicable. conference specific. |
| 12<br>Numbers analysed | Number of participants analysed in each group for the pilot objectives** | p. 2, Results |
| 13<br>Outcome | Results for the pilot objectives, including any expressions of uncertainty** | p. 2, Results |
| 14<br>Harms | Important adverse events or side effects | p. 2, Methods |
| 15<br>Conclusions | General interpretation of the results of pilot trial and their implications for the future definitive trial | p. 2, Conclusions |
| 16<br>Trial registration | Registration number for pilot trial and name of trial register | p. 2, Methods |
| 17<br>Funding | Source of funding for pilot trial | p.2, Methods |

