## Supplementary material for "Low-dose computed tomography and rapid molecular sputum testing in adults with suspected non-ventilator-associated hospital-acquired pneumonia: the HAP-FAST pilot sequential multiple assignment randomised trial with embedded qualitative and costing studies": statistical analysis plan

FUNDED BY

**Feasibility study of the clinical and cost-effectiveness of contemporary diagnostics for patients with suspected Hospital-Acquired Pneumonia (HAP).**

**Trial registration No: NCT05483309**

### Statistical Analysis Plan

### Version 1.0 01/11/2024

| | ORIGINATED BY | QC PERFORMED BY | APPROVED BY |
|---|---|---|---|
| **Name** | Ian Powell | TBC | Dr Ashley Jones |
| **Title** | Trial Statistician | Trial Statistician | Senior Statistician |
| **Date** | 01/11/2024 | | |
| **Protocol Version and Date** | V4.0 12/08/2024 | | |

### 1 Approval and Agreement

**SAP Version Number being approved: V1.0**

**Trial Statistician**

Name Ian Powell

Signed* I Powell Date 4 Nov 2024

box SIGN 1J53LWZ2-17PP599Z

***Signature should be evidenced via physical signature (wet-ink or electronic).**

**Senior Statistician**

Name Dr Ashley Jones

Signed* AP Jones Date 5 Nov 2024

box SIGN 42YWJYX1-17PP599Z

***Signature should be evidenced via physical signature (wet-ink or electronic).**

**Chief Investigator/clinical lead**

Name Dr Daniel Wootton

Signed* Dr Dan Wootton Date Nov 4, 2024

box SIGN 4W9W39V4-17PP599Z

***Signature should be evidenced via physical signature (wet-ink or electronic).**

### 3 Glossary

| | |
|---|---|
| AQUA | Advancing Quality Alliance |
| CI | Chief Investigator |
| CXR | Chest X-Ray |
| CT | Low-dose Thoracic CT Scan |
| CTPA | CT Pulmonary Angiogram |
| CTU | Clinical Trials Unit |
| eCRF | Electronic Case Report Form |
| DDD | WHO defined daily doses |
| DTR | Dynamic Treatment Regimens |
| FAPP | FILMARRAY® Pneumonia Panel |
| HAP | Hospital-Acquired Pneumonia |
| IQR | Interquartile Range |
| ITT | Intention to Treat |
| LCTC | Liverpool Clinical Trials Centre |
| RCT | Randomised Controlled Trial |
| RTI | Hospital Acquired Respiratory Tract Infection |
| SAP | Statistical Analysis Plan |
| SMART | Sequential Multiple Assignment Randomised Trial |
| SOP | Standard Operating Procedure |
| SD | Standard Deviation |
| TSC | Trial Steering Committee |
| VAS | Visual Analogue Scale |

### 4 Roles and Responsibilities

| Name | Affiliations | Role |
|---|---|---|
| Ian Powell (IP) | Liverpool Clinical Trials Centre (LCTC), University of Liverpool | Trial Statistician |
| Dr Ashley Jones (AJ) | Liverpool Clinical Trials Centre (LCTC), University of Liverpool | Senior Statistician |
| Dr Daniel Wootton (DW) | Institute of Infection, Veterinary and Ecological Sciences, University of Liverpool | Chief Investigator |

IP and AJ proposed the statistical analysis plan. IP drafted the statistical analysis plan. AJ and DW read, amended and approved the statistical analysis plan. IP and AJ acknowledge Jessica Green for early development of this statistical analysis plan.

### 5 Statement of Compliance

This Statistical Analysis Plan (SAP) provides a detailed and comprehensive description of the pre-planned analyses for the HAP-FAST study. The planned statistical analyses described within this document are compliant with those specified in brief within the HAP-FAST protocol V4.0 (12/08/2024).

The purpose of the plan is to:

a. Ensure that all analyses are appropriate for the aims of the trial, reflect good statistical practice, and minimise bias by preventing inappropriate post hoc analyses.
b. Ensure that the analyses performed are consistent with the conditions of the protocol.
c. Explain in detail how the data will be handled, covariates derived and analysed to enable the analysis to be replicated, if necessary.

This study is carried out in accordance with the World Medical Association Declaration of Helsinki (1964) and the Tokyo (1975), Venice (1983), Hong Kong (1989) and South Africa (1996) amendments and will be conducted in compliance with the protocol, Liverpool Clinical Trials Centre (LCTC) Clinical Trials Unit (CTU) Standard Operating Procedures (SOPs) and EU Directive 2001/20/EC, transposed into UK law as the UK Statutory Instrument 2004 No 1031: Medicines for Human Use (Clinical Trials) Regulations 2004.

### 6 Introduction

Analyses of accumulating data will be described in a confidential independent oversight committee report. The Trial Steering Committee (TSC) is the only independent oversight committee for HAP-FAST, as the TSC is blinded an open report template will be used for TSC meetings.

The results of the final analysis described within this statistical analysis plan will be contained within a statistical analysis report. This report will be used as the basis of the primary research publications according to the study publication plan. Where analyses are presented which are not included in the SAP, justification as to their inclusion must be provided.

All analyses are performed with standard statistical software (SAS version 9.4 or later, R 4.4.1 or later, and/or other appropriate statistical software). The finalised analysis datasets, programs and outputs will be archived following Good Clinical Practice guidelines and SOP LCTC_GE018 Archiving Procedures.

### 7 Background and Rationale

Hospital-Acquired Pneumonia (HAP) refers to a type of severe lung infection that develops while a patient is in hospital or has been recently discharged. HAP is common, frequently fatal and there is sparse evidence to support its management. Recent guidelines have called for studies focused on diagnostics [1].

There are problems diagnosing the condition; HAP diagnosis relies on a chest X-ray (CXR) but misinterpretation leads to over-diagnosis [2]. There are also problems diagnosing the cause of HAP; sputum culture takes too long to meaningfully impact upon antibiotic decisions. Together, these diagnostic inadequacies contribute to poor clinical outcomes and inappropriate antibiotic usage [3]. CT scans are more accurate than chest X-rays at diagnosing pneumonia but there are no studies to demonstrate impact on outcome in HAP. The close to patient test, 'FILMARRAY® Pneumonia Panel' (FAPP) can identify 28 pneumonia pathogens from a respiratory sample in 75 minutes – but clinical and cost-effectiveness in an NHS setting has not been evaluated in the context of non-ventilator acquired HAP.

The HAP-FAST study will therefore investigate whether using CT scans or the FAPP, or both together, helps improve antibiotic use and patient recovery while being cost effective. Please refer to the "Background" and "Rationale" sections of the HAP-FAST protocol for more details.

### 8 Objectives

The primary objective is to determine the feasibility of a full-scale Randomised Controlled Trial (RCT) comparing different diagnostic dynamic treatment regimens (DTRs) in adult patients suspected of HAP. The primary objective will be determined on the basis of a range of secondary objectives. Please refer to the "Objectives" section of the HAP-FAST protocol for further details on the study objectives.

### 9 Study Design

#### 9.1 Overall study design

HAP-FAST is a feasibility study consisting of a pilot study, two qualitative studies, and a costing analysis. The study participants will also provide clinical samples to support exploratory analyses of the immune-pathophysiology of HAP. Please refer to the "Study Design" section of the HAP-FAST protocol for information on the designs of the qualitative, costing analysis and exploratory sub-studies.

The pilot study is designed as an open, sequential, multiple assignment, randomised trial (SMART) with a 1:1 allocation ratio. Participants are initially randomised between a chest X-ray (CXR) and low-dose thoracic CT scan (CT). Following the imaging, participants whose clinician decides to manage them as either hospital acquired pneumonia (HAP) or hospital acquired respiratory tract infection (RTI), and who are able to produce a sputum sample, are further randomised to 'FILMARRAY® Pneumonia Panel' (FAPP) or no FAPP. All other participants will be managed as per usual care. The randomisation results in 4 dynamic treatment regimens (DTRs), detailed in the table below. Please refer to the "Study Design" section of the HAP-FAST protocol for further details on the design of the pilot study.

| **Dynamic treatment regimen (DTR)** | **Phase 1 intervention** | **Phase 2 intervention** | |
|---|---|---|---|
| | | **Phase 1 indicates HAP/RTI and patient has sputum** | **Phase 1 indicates no HAP/RTI and/or patient has no sputum** |
| DTR 1 | CXR | FAPP | |
| DTR 2 | CXR | No FAPP | No FAPP |
| DTR 3 | CT | FAPP | |
| DTR 4 | CT | No FAPP | No FAPP |

A diagram of the pilot study, detailing the different groups is presented below:

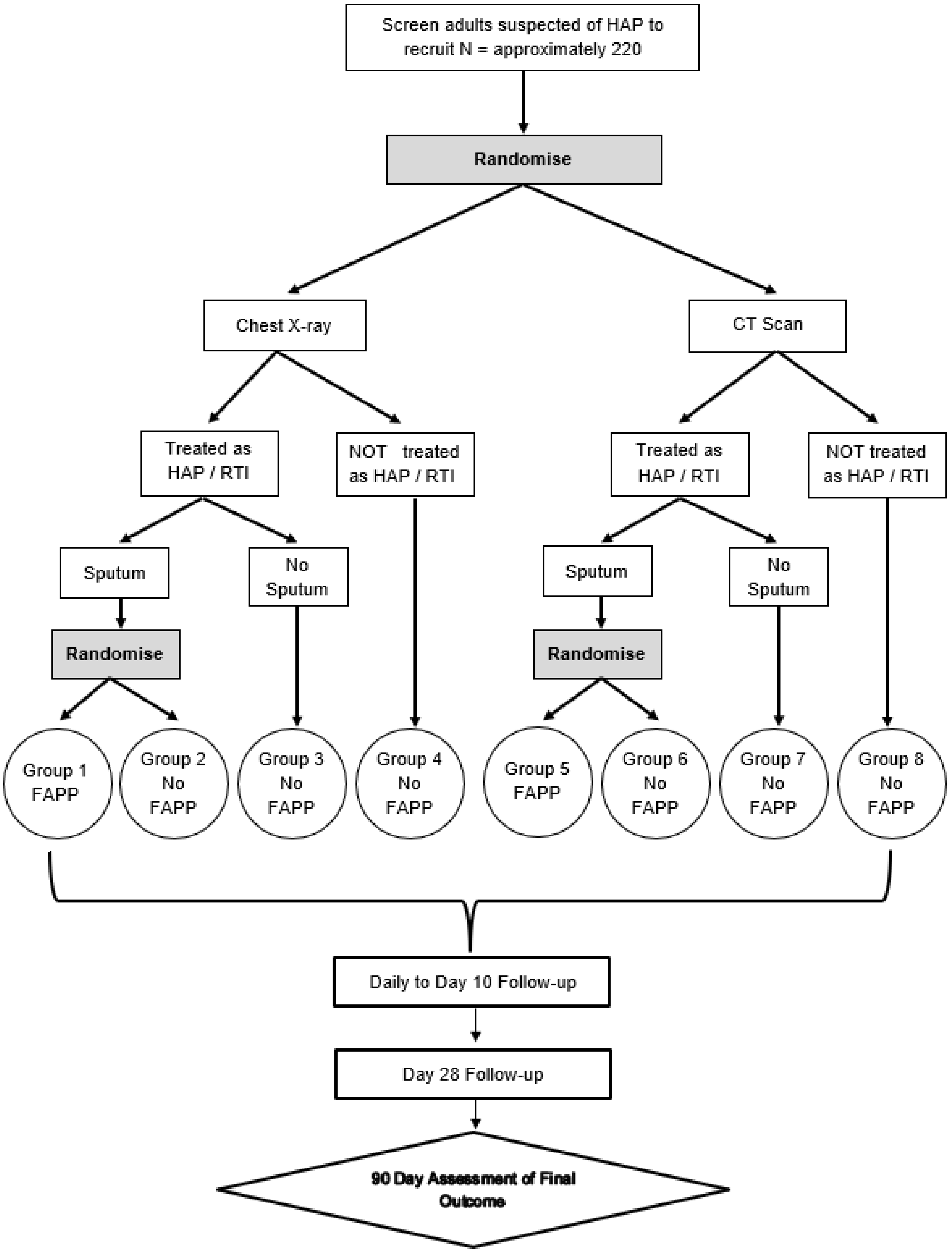

#### 9.2 Blinding

The study is open-label, and treating clinicians, researchers, and participants will know which treatment/intervention is being administered.

### 10 Consent process

Due to the potential severity of HAP, there is a short timeframe of eligibility between HAP being suspected and diagnostic tests being carried out. Moreover, eligible patients, as a consequence of their acute illness and or underlying comorbidities, may have impaired capacity to provide written informed consent and consequently require a consultee for assent. As a result, HAP-FAST will incorporate a deferred consent model as has been used in other emergency situations.

Patients who are randomised but decline consent to continue with the study will have still have all data captured up until they declined included in the analysis. Additionally, patients whose consent is unable to be obtained (e.g. if a patient is discharged from hospital or dies[1] before providing consent) will still have all data captured analysed. Please refer to the "Informed Consent" section of the protocol for further details on the consent process.

### 11 Study population

##### 11.1.1 Inclusion criteria

HAP-FAST has two stages of randomisation, with different inclusion and exclusion criteria. The inclusion criteria for each stage of randomisation can be found in the "Inclusion Criteria" sections of the HAP-FAST protocol.

##### 11.1.2 Exclusion criteria

The exclusion criteria for each stage of randomisation can be found in the "Exclusion Criteria" sections of the HAP-FAST protocol.

[1] If a participant dies before providing consent, their next of kin will be notified of their participation in the trial. Relatives and friends are not able to consent on behalf of the deceased participant. The data captured whilst they were alive will remain in the study unless the relatives express recollection of them having very strong negative views about research in which retention of data will be considered on a case by case basis.

#### 11.2 Removal of participants from intervention or follow-up

Removal of participants from intervention is not applicable. Randomisation for HAP-FAST pertains to diagnosis strategies - the participants' treatment will ultimately be at the discretion of the treating clinician.

Participants are free to withdraw from follow up at any time without providing a reason, though a reason should be recorded if one is given. There are no pre-determined reasons for withdrawal from follow-up. There are two types of withdrawal:

- Withdrawal from research samples only,
- Withdrawal of consent for ALL further follow up.

Those who wish to withdraw from further follow-up will have the data collected up to the point of that withdrawal included in the analyses.

Please refer to the "Intervention Discontinuation and Participant Discontinuation/Withdrawal" section of the "HAP-FAST" protocol for further details on withdrawal from follow-up.

### 12 Method of assignment to intervention

There are two stages of randomisation in the pilot study. Both will use a secure (24-hour) web-based randomisation system controlled centrally by the LCTC. Randomisation lists were generated with random variable block sizes and stratified by site.

**Randomisation 1: Choice of imaging**

Participants will be randomised to undergo either a CT scan or chest X-ray (in a ratio of 1:1).

**Randomisation 2: FAPP or No FAPP**

Once imaging has been completed, and a clinical judgement is made, participants who are:

- to be treated as HAP or a hospital acquired RTI and
- able to produce a sputum sample

will be randomised to FAPP or No FAPP (in a ratio of 1:1).

### 13 Schedule of assessments

Please refer to the "Schedule for Assessments and Follow-up" section of the HAP-FAST protocol for further details on the schedule of assessments. The study flow diagram can be found in the "Schematic of Study Design" section of the HAP-FAST protocol.

### 14 Interventions

The pilot study has a SMART design, where the randomisation pertains to diagnostic strategies which may or may not affect treatments received. In general, choice of treatment will be determined by the diagnostic information available to clinicians. There are eight distinct possible routes through the study. Participants' treatment will ultimately be at the discretion of the treating clinician. A summary of which approach to take based on the participant's flow through the study is given in the table below. Please refer to the "Trial Treatment/Interventions" section of the HAP-FAST protocol for more information.

| Result of Stage 1 Randomisation | Result of Imaging | Sputum Available? | Result of Stage 2 Randomisation | Treatment | Group |
|---|---|---|---|---|---|
| CXR | Clinician decides to treat for HAP /hospital acquired RTI | YES | FAPP | • Use an aliquot of respiratory specimen in the FAPP<br>• Send remainder of specimen to microbiology for standard tests<br>• Prescribe antibiotics with reference to the FAPP antibiotic guideline | 1 |
| | | | No FAPP | • Prescribe empirical antibiotics based on local guidelines | 2 |
| | | NO | N/A | • Prescribe empirical antibiotics based on local guidelines | 3 |
| | Clinical diagnosis is **not** HAP/RTI | N/A | N/A | • Patient receives usual care and is followed up as per the study schedule | 4 |
| CT Scan* | Clinician decides to treat for HAP/ | YES | FAPP | • Use an aliquot of respiratory specimen in the FAPP | 5 |

| | | | | | |
|---|---|---|---|---|---|
| | hospital acquired RTI | | | • Send remainder of specimen to microbiology for standard tests<br>• Prescribe antibiotics with reference to the FAPP antibiotic guideline | |
| | | | No FAPP | • Prescribe empirical antibiotics based on local guidelines | 6 |
| | | NO | N/A | • Prescribe empirical antibiotics based on local guidelines | 7 |
| | Clinical diagnosis is **not** HAP /RTI | N/A | N/A | • Patient receives usual care and is followed up as per the study schedule | 8 |

* Low-dose, non-contrast, CT scan of the thorax “hot reported”.

### 15 Listing of Outcomes

#### 15.1 Primary outcome(s)

The primary objective is determining the feasibility of a future definitive RCT. This will be answered by assessing the secondary outcomes.

#### 15.2 Secondary outcome

The secondary outcomes in the table below will be used to determine the final decision as to whether a definitive study is feasible:

| Objective | Outcome |
|---|---|
| Inform the sample size of a definitive study | (1) Time to clinical cure:<br>Defined as the number of days from baseline when there is a combination of resolution of signs and symptoms present at enrolment and improvement or lack of progression of radiological signs |
| | (2) Antibiotic usage for the HAP episode |
| | (3) EQ-5D-5L |
| | (4) Length of hospital stay post HAP diagnosis |
| | (5) Mortality:<br>Day 14, 28 and 90 |
| To measure key outcome measures (completion rates, missing data, estimates and dispersion) | (6) Outcome data completion:<br>Estimate rates of completion of questionnaires – EQ-5D-5L, CAP-sym, economic evaluation |

| | |
|---|---|
| | Summary statistics and proportion of missing data for time to clinical care, antibiotic usage for HAP diagnosis, EQ-5D-5L, length of hospital stay post HAP diagnosis, mortality |
| To estimate eligibility, recruitment and consent rates | (7) Eligibility, recruitment and consent rates:<br>• Rate of recruitment,<br>• Proportion screened that meet eligibility criteria**,<br>• Proportion eligible that consent and where they present**.<br>(8) Rates of successful follow-up:<br>• Proportion consented and randomised that complete study pathway as per protocol,<br>• Proportion consented and randomised that withdraw from study intervention or follow up**. |
| Estimate rates of successful follow up | (8) Rates of successful follow up:<br>• Proportion consented and randomised that complete study pathway as per protocol,<br>• Proportion consented and randomised that withdraw from study intervention or follow up** |
| Evaluate adherence to antibiotic guidelines and study protocol | (9) Summary statistics relating to antibiotic use in the pilot study with a comparison between the DTRs |
| Assess the web-based randomisation process and incorporate clinical and researcher feedback | (10) Qualitative conclusions based on staff focus groups |
| Assess human factors involved in delivery of the study and how the different diagnostic tests influence clinical decision making by conducting qualitative interviews and focus groups with healthcare workers and researchers | (10) Qualitative conclusions based on staff focus groups |
| Evaluate willingness of clinicians to recruit to the study | (10) Qualitative conclusions based on staff focus groups |
| Evaluate willingness of potential participants or their consultees to be recruited | (11) Qualitative conclusions based on participant and carer interviews |
| Assess the study participant and carer experience of participating in the study | (12) Qualitative interviews |
| Perform a costing analysis of HAP to inform the cost-effectiveness analysis for any definitive study | (13) Summary statistics for numbers and types of costs with comparison between DTRs |

** Reasons why, and stage will be collected to inform future trial design.

#### 16 Sample size calculation

Since HAP-FAST is a feasibility study, there is no sample size calculation. A justification for the sample size can be found in the “Sample Size” section of the HAP-FAST protocol.

### 17 Study Framework

The overall objective of the study is to determine the feasibility of a future definitive RCT, as measured by the secondary outcomes.

### 18 Timing and Objectives of Analyses

#### 18.1 Interim Reporting

##### 18.1.1 Reports to Independent Oversight Committees

Analyses of the accumulating data will be performed at regular intervals (at least biannually) for review by the TSC.

##### 18.1.2 Assessments of progression criteria

There are no progression criteria to assess.

##### 18.1.3 Formal Interim Analyses

There are no formal interim analyses planned.

#### 18.2 Final Analysis

The final analysis should take place once all participants have completed their 90-day follow-up. However, the trial may be closed prematurely by the TSC. All outcomes will be analysed once the end of study has been reached - the end of study is defined to be the date on which data for all participants is frozen and data entry privileges are withdrawn from the study database.

### 19 Disposition of Participants

Due to the nature of the study and the urgent need to treat, standard screening logs will not be maintained. As soon as a patient is identified as having suspected HAP, they will be assessed for eligibility and included in the study (if eligible). For participants who are assessed for eligibility but not randomised at stage one, ineligibility reason(s) will be recorded by the online randomisation system.

A CONSORT flow diagram [5] [6] will be used to summarise the number of patients who were:

Note: Anything in italics will only be presented in the final statistical analysis report and not in TSC reports.

- Assessed for eligibility,
- Ineligible at screening,

- Randomised to stage 1,
- *Received allocated stage 1 intervention (split by stage 1 randomisation),*
- Assessed for stage 2,
- Assessment for stage 2 not recorded*,
- Ineligible for stage 2,
- Randomised to stage 2,
- *Received allocated stage 2 intervention (split by stage 2 randomisation),*

*For the final statistical analysis report, the following will be split by allocation pathway[2]:*

- Consent not obtained,
- Did not provide consent,
- Consent provided,
- Lost to follow-up,
- Withdrew from follow-up,
- Died,
- Included in the analysis for each of secondary outcomes 1-5.

∗ The number of patients whose assessment for eligibility for stage 2 was not recorded and who were not able to produce a sputum sample before their second dose of antibiotic will be presented.

#### 19.1 Screening, eligibility and recruitment

Screening data will be summarised overall and by site in two tables detailing:

Note: Anything in italics will only be presented in the final statistical analysis report and not in TSC reports.

**Stage 1**

i. Those who were assessed for eligibility, expressed as a frequency,
ii. Those who met the eligibility criteria at screening for stage 1, expressed as a frequency and a percentage with the denominator being all patients assessed for eligibility (ii),
iii. Those who did not meet the eligibility criteria at screening for stage 1, expressed as a frequency and a percentage with the denominator being all patients assessed for eligibility (ii),
iv. Those who were eligible and randomised to stage 1, expressed as a frequency and a percentage with the denominator being all eligible patients for stage 1 (iii),

---

[2] Allocation pathways: (1) CT only, (2) CXR only, (3) CT/ FAPP, (4) CXR/ FAPP, (5) CT/ No FAPP and (6) CXR/ No FAPP.

v. Those who were randomised to stage 1 and assessed for eligibility for stage 2, expressed as a frequency and a percentage with the denominator being all patients randomised to stage 1 (v),

vi. Those who were randomised to stage 1 but their assessment for eligibility for stage 2 was not recorded, expressed as a frequency and a percentage with the denominator being all patients randomised to stage 1 (v),

vii. Those whose eligibility assessment for stage 2 was not recorded and provided consent, expressed as a frequency and percentage with the denominator being all patients whose eligibility assessment for stage 2 was not recorded (vii),

viii. Those whose eligibility assessment for stage 2 was not recorded and consent was not able to be obtained from them, expressed as a frequency and percentage with the denominator being all patients whose eligibility assessment for stage 2 was not recorded (vii),

ix. Those whose eligibility assessment for stage 2 was not recorded and did not provide consent, expressed as a frequency and percentage with the denominator being all patients whose eligibility assessment for stage 2 was not recorded (vii).

**Stage 2**

x. Those who were assessed for eligibility for stage 2,

xi. Those who met the eligibility criteria for stage 2, expressed as a frequency and a percentage with the denominator being all patients assessed for eligibility for stage 2 (xi),

xii. Those who did not meet the eligibility criteria for stage 2, expressed as a frequency and a percentage with the denominator being all patients assessed for eligibility for stage 2 (xi),

xiii. Those who did not meet the eligibility criteria for stage 2 and provided consent, expressed as a frequency and a percentage with the denominator being all ineligible patients for stage 2 (xiii),

xiv. Those who did not meet the eligibility criteria for stage 2 and consent was not able to be obtained from them, expressed as a frequency and a percentage with the denominator being all ineligible patients for stage 2 (xiii),

xv. Those who did not meet the eligibility criteria for stage 2 and did not provide consent, expressed as a frequency and a percentage with the denominator being all ineligible patients for stage 2 (xiii),

xvi. Those who were eligible and randomised to stage 2, expressed as a frequency and a percentage with the denominator being all eligible patients for stage 2 (xii),

xvii. Those who were randomised to stage 2 and provided consent, expressed as a frequency and a percentage with the denominator being all patients randomised to stage 2 (xvii),

xviii. Those who were randomised to stage 2 and consent was not able to be obtained from them, expressed as a frequency and a percentage with the denominator being all patients randomised to stage 2 (xvii),

xix. Those who were randomised to stage 2 and did not provide consent, expressed as a frequency and a percentage with the denominator being all patients randomised to stage 2 (xvii).

Reasons for ineligibility for stage 1 will be summarised overall and by site in a table with the following categories:

- Aged less than 18 years,
- Does not have suspected HAP,
- Already received a chest X-ray to confirm suspected HAP diagnosis,
- Diagnosis or suspected diagnosis of ventilator acquired pneumonia,
- Intention to palliate rather than cure,
- Interventions cannot be completed before administration of second antibiotic dose,
- Cannot be randomised to low-dose, non-contrast CT scan on clinical grounds e.g. strong suspicion of PE,
- Pregnant,
- Previous study participation (patients with second of third episodes of HAP will not be re-recruited).

Frequencies will be presented along with percentages using all patients ineligible for stage 1 (iv) as the denominator.

Reasons for ineligibility for stage 2 will be overall and by site in a table with the following categories:

- Following the CXR or CT the clinician does not intend to treat the patient for HAP or a hospital acquired RTI,
- A sputum sample cannot be obtained before the second dose of antibiotic,
- Following the CXR or CT the clinician decides not to treat with antibiotics for either HAP or hospital acquired RTI.

Frequencies will be presented along with percentages using all patients ineligible for stage 2 (xiii) as the denominator.

Reasons for patients who were only randomised to stage 1 declining consent will be grouped into similar terms and the grouped reasons summarised overall and by site in a table. Frequencies will be presented along with percentages using all patients who were only randomised to stage 1 and did not provide consent (x + xvi) as the denominator.

Reasons for patients who were randomised to stages 1 and 2 declining consent will be grouped into similar terms and the grouped reasons summarised overall and by site in a table. Frequencies will be presented along with percentages using all patients who were randomised to stages 1 and 2 and did not provide consent (xx) as the denominator.

A recruitment summary table will be presented showing the following for each site: site code, site name, date site opened to recruitment, dates of first/last randomisations, total number randomised (to stages 1 and 2) and total number did not provide consent. The percentage of participants randomised to stage 1 who were randomised to stage 2 and the percentage who did not provide consent will be presented using the total number randomised to stage 1 as the denominator.

A recruitment graph detailing the expected and actual number of sites opened and the expected (stage 1) and actual (stages 1 and 2) number of participants randomised each month will be presented.

Additionally, for TSC reports the number randomised to each stage by calendar month will be presented.

#### 19.2 Post randomisation discontinuations

A summary sentence detailing the number and percentage of randomised patients who did not provide consent and who consent was unable to be obtained from will be provided. The total number of participants lost to follow-up will be reported and any available reasons provided in the CONSORT diagram.

Any deaths will be reported in a line listing detailing site code, site, stage 1 allocation (CT or X-ray), timing of death (in days since stage 1 randomisation) and cause of death.

Other withdrawals will be presented as line listings detailing site code, site, stage 1 allocation, timing of withdrawal (in days since stage 1 randomisation), who made the decision to withdraw the participant, the reason for withdrawal and for participant/consultee withdrawals, the type of withdrawal.

### 20 Protocol Deviations

Possible protocol deviations (that are considered to negatively impact on generalisability, participant safety or the integrity of trial results) are specified in the HAP-FAST Monitoring Plan V1.0 (16/05/2023). The number (and percentage) of participants experiencing each protocol deviation will be presented overall and by stage 1 allocation, along with the number of participants with at least one protocol deviation. The participants that are included in the intention to treat (ITT) analysis data set will be used as the denominator to calculate the percentages. No formal statistical testing will be undertaken.

### 21 Unblinding

Not applicable.

### 22 Analysis Datasets

#### 22.1 Efficacy Analyses

The principle of intention-to-treat, as far as practically possible, will be the main strategy of the analysis adopted for all of the outcomes. These analyses will be conducted on all randomised participants (or all consenting participants for outcomes incorporating questionnaires), in the group to which they were allocated, and for whom the outcome(s) of interest have been observed/measured.

#### 22.2 Safety Analyses

Not applicable. Since HAP-FAST only incorporates well-established and non-invasive diagnostic investigations that would normally be carried out as standard of care, safety events will not be recorded as part of this study.

### 23 Baseline Characteristics

Descriptive statistics will be reported split by stage 1 allocation. Categorical data will be summarised by counts and percentages. Continuous data will be summarised by mean, standard deviation (SD), median, interquartile range (IQR) and range. Percentages will be reported to one decimal place; continuous summaries will be presented to one decimal place or one significant figure (if there are no significant figures before two decimal places). Variables to be included in the table are as follows:

| **Variable** | **Electronic case report form (eCRF)** |
|---|---|
| Number randomised | Randomisation Stage 1 eCRF |
| Number consent provided | Informed Consent eCRF |
| Number consent not obtained | |
| Number did not provide consent | |
| **Demographic details** | |
| Age in years (Continuous) | Day 0 Baseline eCRF |
| Sex<br>(Male, Female) | Randomisation Stage 1 eCRF |
| Height in cm (Continuous) | Day 0 Baseline eCRF |
| Weight in kg (Continuous) | |
| Calculated BMI (Continuous) | |
| Smoking status<br>(Current smoker, Ex-smoker, Never smoked, Not known) | Day 0 Admission Related Data eCRF |
| **Vital signs (at time of suspected HAP)**[3] | |
| Temperature in degrees Celsius | Day 0 Baseline eCRF |
| Systolic blood pressure in mmHg | |
| Diastolic blood pressure in mmHg | |
| Pulse rate in bpm | |
| Respiratory rate in bpm | |
| Oxygen saturation on room air in % | |
| NEWS2 Score | |
| **Inpatient details and HAP signs & symptoms** | |
| Ward type<br>(Medical, Surgical, A&E, Acute medical unit, Other, Not known) | Day 0 Baseline eCRF |
| Clinical symptoms of HAP<br>(New/worsening shortness of breath, Cough, Fever, Chest pain, New/worsening sputum, Confusion, Other) | |
| Clinical description of respiratory finding of HAP<br>(New chest crackles, Reduced air entry, Respiratory distress, Other) | |
| 'Medically fit for discharge' prior to HAP suspicion<br>(Yes, No, Not known) | |
| **Concomitant medications and immunosuppressant medication use** | |
| Antibiotics taken/received in last 7 days<br>(Yes, No) | Day 0 Baseline eCRF |
| Received immune suppressant mediation (excluding cancer treatment) in the 3 months prior to suspected HAP | |

[3] For each vital sign, the number of participants with the vital sign recorded as 'Not known' or 'Not collected' will also be presented.

| | |
|---|---|
| (Yes, No, Not known) | |
| Received chemotherapy for a solid organ or haematological cancer in the 3 months prior to suspected HAP<br>(Yes, No, Not known) | |
| Taken or received the following in the last 7 days:<br>Steroids (oral, IV, inhaled)<br>(Yes, No)<br>Antipsychotics<br>(Yes, No)<br>Proton pump inhibitors<br>(Yes, No)<br>Statins<br>(Yes, No)<br>None of the above<br>(Yes, No) | |
| **Admission details** | |
| Time from admission to HAP suspected (Continuous) | Day 0 Admission Related Data eCRF and Randomisation Stage 1 eCRF |
| Reason for admission<br>(Medical, Surgical) | Day 0 Admission Related Data eCRF |
| Had the following conditions at the point of admission<br>(None of the below, Congestive cardiac failure, Dementia, Other form of cognitive impairment, Myocardial Infarction, Chronic pulmonary disease, Rheumatic disease, AIDS (the syndrome not just HIV positive), Hemiplegia or paraplegia, Renal disease, Liver disease, Diabetes, Any malignancy) | |
| Oxygen saturation on room air in % at time of admission (Continuous) – the number of participants with 'Not known' for oxygen saturation will also be presented | |
| Could eat and drink independently prior to admission<br>(Yes, No, Not known) | |
| Clinical frailty score for ages 65 and over only)<br>(1, 2, 3, 4, 5, 6, 7, 8, 9, N/A (under 65), Not known) | |

### 24 Compliance with Interventions

The time from stage 1 randomisation to scan and time from stage 1 randomisation to scan report will be presented in hours split by stage 1 allocation (CT or CXR) and site using the following summaries: mean, SD, median and IQR. Additionally, a summary table detailing the number and percentage of participants who did not receive their stage 1 allocated intervention will be presented. The summary table will further break this down by the reasons for not receiving their allocated intervention.

A summary table detailing the number and percentage of participants allocated to FAPP who did not receive FAPP will also be presented. The summary table will further break this down by the reasons for FAPP not being received.

### 25 Analysis of Outcomes

#### 25.1 Interim Reporting

##### 25.1.1 For reports to independent oversight committees

The number (and percentage) of participants with complete outcome data (out of those expected to have outcome data at a given timepoint) will be reported overall.

##### 25.1.2 Assessment of progression criteria

There a no progression criteria to assess.

##### 25.1.3 Formal Interim Analysis

There are no formal interim analyses planned.

#### 25.2 Final Analysis

For each outcome, the following treatment arm comparisons will be made: CXR vs CT (groups 1-4 vs groups 5-8 in the table in section 14) and FAPP vs no FAPP (groups 1 and 5 vs groups 3 and 6).

Percentages will be reported to one decimal place throughout; mean, median, SD and IQR will be presented to one decimal place or one significant figure (if there are no significant figures before two decimal places); all other values will be presented to two decimal places or one significant figure (if there are no significant figures before three decimal places).

##### 25.2.1 Levels of significance and multiplicity

Not applicable. There are no formal hypotheses being tested. For comparisons between treatment arms, 95% confidence intervals will be presented but no inference will be drawn – all results will be treated as hypothesis generating.

##### 25.2.2 Primary Outcome

The key objective and primary outcome of this trial is determining the feasibility of a future definitive randomised controlled trial. This will be answered by assessing the secondary outcomes.

###### 25.2.2.1 Derivation

Not applicable.

###### 25.2.2.2 Analysis

Not applicable.

##### 25.2.3 Secondary Outcomes

###### 25.2.3.1 Secondary outcome 1: Time to clinical cure

###### 25.2.3.1.1 Derivation

If a participant has the question 'Report findings: Were features consistent with Pneumonia?' answered with 'Yes' on the 'CT/X-Ray Details' eCRF, they will be classed as being diagnosed with HAP.

Time to clinical cure is defined as the number of days from day 0 (date of suspected HAP) till there is a combination of:

- Resolution of signs and symptoms present at enrolment,
- And improvement or lack of progression of radiologic signs.

The table below details the criteria that must be met for clinical cure:

| **Sign/symptom/radiological sign** | **Criteria required for clinical cure** | **eCRF** |
|---|---|---|
| Temperature | ≤ 37.2 °C | Day 0 baseline eCRF<br>Daily (Day 1-10) eCRF |
| Heart rate | ≤ 100 beats per minute | |
| Respiratory rate | ≤ 24 breaths per minute | |
| Systolic blood pressure | ≥ 90 mmHg | |
| Oxygen saturation on room air | ≥ 90%<br>or<br>Return to pre-HAP state | Day 0 admission related data eCRF<br>Daily (Day 1-10) eCRF |
| Ability to maintain oral intake (eat and drink independently) | Ability to eat and drink independently (answered with 'Yes')<br>or<br>Return to pre-HAP state | |

| | | |
|---|---|---|
| Cognitive function | Regained pre-HAP level of cognitive function | Daily (Day 1-10) eCRF |
| Radiological scan result (CT, CXR or CTPA) up to day 10 | Findings of any repeat chest imaging are not 'Progression of pneumonia' | Day 90 visit eCRF (only interested in repeat scans that occur up to day 10) |
| CAP-Sym[a] | Percentage improvement in CAP-Sym score from baseline* of 35% or more. The percentage change is calculated as follows: $100 \times \frac{Score_{D7-10} - Score_{D0-3}}{Score_{D0-3}}$ <br> * Only baseline questionnaires completed between days 0 and 3 will be considered analysable for this outcome | Baseline Questionnaire<br>Daily CAP-SYM<br>Day 10 or Discharge Questionnaire |

[a] The CAP-Sym consists of 18 items, each item is scored on a six-point Likert scale (0-5), with lower scores indicating the symptom/problem is less bothersome for the participant. All questions must be completed for the questionnaire to be able to be included in the analysis. The total CAP-Sym score is calculated by adding the score for each item together.

If a participant meets all of the criteria in the table above, they will be classed as achieving clinical cure. All participants may not achieve clinical cure, and participants that do not achieve clinical cure will be classed as censored. The time to clinical cure/censoring time will be computed as the number of days from stage 1 randomisation (day 0) to the date of one of the following scenarios:

| **Scenario** | **Date** |
|---|---|
| (1) Participant achieves clinical cure | Date of daily visit they achieved clinical cure |
| (2) Participant is discharged before their day 10 daily visit without achieving clinical cure | Date of discharge |
| (3) Participant dies before they are discharged and their day 10 daily visit without achieving clinical cure | Date their day 10 daily visit would have been |
| (4) Participant withdraws/is withdrawn before they are discharged and their day 10 daily visit without achieving clinical cure | Date of withdrawal |
| (5) Participant does not achieve clinical cure by their day 10 daily visit | Date of their day 10 daily visit |

A censoring indicator will be calculated as follows:

$$\text{censor} = \begin{cases} 1 \text{ if scenario } 1, \\ 0 \text{ if scenario } 2, 3, 4 \text{ or } 5. \end{cases}$$

###### 25.2.3.1.2 Analysis

The analysis for this outcome will include all randomised participants. Times to clinical cure will be summarised for each treatment arm using Kaplan-Meier plots with separate curves for each treatment arm. Two plots will be constructed: (1) FAPP vs no FAPP and (2) CXR vs CT. Additionally, the Kaplan-Meier method will be used to estimate the median time to clinical cure and 95% confidence interval (if possible) for each treatment arm.

To assess the feasibility of this outcome:

- For those who meet the criteria for clinical cure, the number and percentage who did and did not fail clinical cure at a later visit will be presented for each treatment arm. The denominator will be all participants who were randomised and achieved clinical cure during 10-day follow-up.
- For each treatment arm, the percentage change in CAP-Sym score from baseline to time of clinical cure will be presented using the following summaries: mean, SD, median and IQR.
- The number and percentage of participants who (1) failed clinical cure and (2) could not be assessed for clinical cure will be presented for each treatment arm. The denominator will be all those who did not experience clinical cure for that treatment arm.
    (1) Those who failed clinical cure will be further broken down[4] into those who failed on the:
        - Clinical stability component (temperature - cognitive function in the table above),
        - Radiological component,
        - CAP-Sym component.

    (2) Those who could not be assessed for clinical cure will be further broken down into those who:
        - Were discharged without repeat chest imaging,
        - Died without repeat chest imaging,
        - Did not consent by day 3,
        - Declined consent between days 4 and 6,
        - Withdrew consent before day 7,

[4] Participants may be assessed for clinical cure multiple times (up to day 10). Only the participant's best result up to day 10, i.e. the result from the day they failed on the least components, will be summarised. Participants may have failed on more than one component.

  - Had no repeat chest imaging up to day 10,
  - Had incomplete data[5].

- A sensitivity analysis may be conducted to analyse time to cure excluding the *Radiological scan result* criterion. This is to explore if the estimated time to cure is subject to selection bias, as patients with suspected HAP who are not formally treated as HAP/RTI patients may not be indicated for repeat chest imaging.

Comparisons between treatment arms (FAPP vs no FAPP and CXR vs CT) will be conducted using Cox Proportional Hazards models. For each comparison, the hazard ratio and 95% confidence interval will be presented. Cox Proportional Hazards models rely on the proportional hazards assumption. This assumption will be checked by inspection of the Kaplan-Meier plots.

###### 25.2.3.2 Secondary outcome 2: Antibiotic usage for the HAP episode

###### 25.2.3.2.1 Derivation

Antibiotics received by each participant will be recorded on the ‘Antibiotic Use’ eCRF, only antibiotics provided:

- Between the date of stage 1 randomisation and the end of antibiotic treatment for the HAP episode that made the patient eligible for the trial, and
- Indicated for ‘HAP/RTI’

will be considered as antibiotics used for the HAP episode.

Antibiotic usage will be reported in terms of WHO defined daily doses (DDD). The DDD is the assumed average maintenance dose per day for a drug used for its main indication in adults. For each antibiotic, the antibiotic usage in DDDs will be calculated as follows:

$$\text{Antibiotic usage (in DDDs)} = \frac{\text{Doses received} \times \text{Amount per dose (in same units as DDD)}}{\text{DDD}^{6}}.$$

For each participant with suspected HAP, the antibiotic usage in DDDs for each antibiotic they received for the HAP episode will be added together to compute their total antibiotic usage in DDDs.

[5] Participants may be assessed for clinical cure multiple times (up to day 10). If they never had complete data to assess clinical cure on any of these days, they are classed as having incomplete data.
[6] An antibiotic may have a different DDD depending on the route of administration.

###### 25.2.3.2.2 Analysis

The analysis for this outcome will include all randomised participants. For analysis of CXR versus CT scan, this means all patients who were randomised, including those where HAP was not confirmed. For analysis of FAPP versus no FAPP, all patients randomised in the second arm will be analysed.

For each treatment arm, total antibiotic usage (in DDDs) will be presented using the following summaries: mean, SD, median and IQR. Additionally, boxplots summarising total antibiotic usage (in DDDs) will be presented for each treatment arm.

Comparisons between treatment arms (FAPP vs no FAPP and CXR vs CT) will be conducted using Poisson regression. For each comparison, the incidence rate ratio and 95% confidence interval will be presented. Models will be checked for violations in the assumptions (especially integer response, zero-inflation and overdispersion), and an appropriate alternative model will be selected if violations are present.

##### 25.2.3.3 Secondary outcome 3: EQ-5D-5L

###### 25.2.3.3.1 Derivation

The EQ-5D-5L is completed at baseline, day 10[7], day 28 and day 90. The EQ-5D-5L consists of five dimensions (mobility, self-care, usual activities, pain/discomfort and anxiety/depression), each of the five dimensions consists of five levels of severity. Participants will indicate their health state by checking the box next to the most appropriate response level of each of the five dimensions. Additionally, the EQ-5D-5L contains a visual analogue scale (VAS) score that records the participants' self-rated health [6] [8].

###### 25.2.3.3.2 Analysis

The analysis for this outcome will include all randomised participants who provided consent.

For each timepoint (baseline, day 10, day 28 and day 90), the number and percentage of participants in each severity level for each dimension will be presented split by treatment arm. The denominator will be all participants who were randomised to that treatment arm and provided consent.

---

[7] The first post-baseline EQ-5D-5L is completed at discharge or day 10 if participants have not been discharged before then.

For each timepoint (baseline, day 10, day 28 and day 90), the VAS score will be presented using the following summaries: mean, SD, median and IQR split by treatment arm. Additionally, boxplots summarising the VAS score will be presented for each timepoint and treatment arm.

Change from baseline VAS score summaries (mean, SD, median and IQR) and boxplots will be presented for each timepoint (day 10, day 28 and day 90) and treatment arm. Comparisons between treatment arms (FAPP vs no FAPP and CXR vs CT) will be conducted using linear regression, adjusted for baseline VAS score. For each comparison, the mean difference between treatment arms and 95% confidence interval will be presented.

###### 25.2.3.4 Secondary outcome 4: Length of hospital stay post HAP diagnosis

###### 25.2.3.4.1 Derivation

If a participant has a 'Discharge CRF' they will be classed as being discharged from hospital. Length of hospital stay post HAP diagnosis will be analysed separately as both time to event and continuous.

**Time to event**

Patients with suspected HAP may not be discharged from hospital before their 90-day follow-up; participants that are not discharged will be classed as censored. The length of hospital stay/censoring time will be computed as the number of days from suspected HAP to the date of one of the following scenarios:

| **Scenario** | **Date** |
|---|---|
| (1) Participant is discharged from hospital | Date of discharge from hospital |
| (2) Participant dies before they are discharged | Date of the end of the visit window for what would have been their 90-day follow-up (90 days + 14) |
| (3) Participant withdraws/is withdrawn before they are discharged | Date of their withdrawal |
| (4) Participant is not discharged by their 90-day follow-up | Date of their 90-day follow-up |

A censoring indicator will be calculated as follows:

$$\text{censor} = \begin{cases} 1 \text{ if scenario } 1, \\ 0 \text{ if scenario } 2, 3 \text{ or } 4. \end{cases}$$

###### 25.2.3.4.2 Analysis

The analysis for this outcome will include all randomised participants.

For the time to event analysis, length of hospital stay post stage 1 randomisation will be analysed in the same way as time to clinical cure in section 25.2.3.1.2. However, the denominator will be all participants who were randomised and provided consent. Length of hospital stay post HAP diagnosis will also be presented using the following summaries: mean, SD, median and quartiles split by treatment arm. Kaplan-Meier estimates for cumulative probability of discharge will be presented to inform about differences in distribution of length of stay.

###### 25.2.3.5 Secondary outcome 5: Mortality

###### 25.2.3.5.1 Derivation

Deaths will be recorded on the 'Death Reporting' CRF. If a participant dies (of any cause) on or before day 14 (where day 0 is the date of stage 1 randomisation), they will be classed as a mortality by day 14. If participants withdraw on or before day 14 they will not be classed as a mortality. Mortalities by day 28 and day 90 will be determined using the same method as for day 14.

###### 25.2.3.5.2 Analysis

The analysis for this outcome will include all randomised participants.

For each timepoint (day 14, day 28 and day 90) and treatment arm, the number and percentage of participants as calculated by Kaplan-Meier estimates will be presented. Patients who withdrew consent will be included up to their withdrawal date, at which point they will be censored. The risk difference of mortality and 95% confidence interval will be presented for each treatment arm comparison (FAPP vs no FAPP and CXR vs CT) at each timepoint.

###### 25.2.3.6 Secondary outcome 6: Completion of outcomes

###### 25.2.3.6.1 Derivation

For secondary outcomes 2, 4 and 5, participants will only have the eCRFs used in the derivation ('Antibiotic Use', 'Discharge' and 'Death Reporting') of the outcomes if they had antibiotics, were discharged from hospital or died. Therefore, completion of outcomes is not applicable for these secondary outcomes.

**Time to clinical cure**

A participant will be classed as not being able to be assessed for clinical cure if they:

- Were discharged without repeat chest imaging,
- Died without repeat chest imaging,

- Did not consent by day 3,
- Declined consent between days 4 and 6,
- Withdrew consent before day 7,
- Had no repeat chest imaging up to day 10,
- Had incomplete data – participants may be assessed for clinical cure multiple times (up to day 10). If they never had complete data to assess clinical cure on any of these days, they are classed as having incomplete data.

**Questionnaires**

For the following questionnaires and timepoints, the number of questionnaires expected, returned and completed will be computed:

| Questionnaire | Timepoint |
|---|---|
| EQ-5D-5L | Baseline, day 10 or discharge (if discharged before day 10), day 28 and day 90 |
| CAP-Sym | Baseline, day 1 to day 10 or discharge (if discharged before day 10), day 28 and day 90 |
| Post-discharge indirect cost survey | Day 90 |

- Questionnaires are expected if the participant has provided consent prior to that timepoint. For the daily CAP-Sym questionnaires, if the participant is discharged before the timepoint the questionnaire is not expected.
- Questionnaires will be classed as returned if at least one question has been completed.
- Questionnaires will be classed as completed if they have been completed to an analysable standard and within the visit window (for day 28 this is +/- 7 days and for day 90 this is +/- 14 days):
    - The CAP-Sym is analysable if all 18 items have been answered.
    - For the EQ-5D-5L, separate completion rates will be computed for each of the five dimensions and VAS score.
    - For the Post-discharge indirect cost survey, the completion rate for each question will be computed.

The denominator for the return and completions rates will be the number of questionnaires expected.

###### 25.2.3.6.2 Analysis

**Time to clinical cure**

A summary sentence detailing the number and percentage of participants who could not be assessed for clinical cure will be provided and will be broken down by the reasons clinical cure could not be assessed. The denominator will be all participants who were diagnosed with HAP, i.e. the question 'Report findings: Were features consistent with Pneumonia?' is answered with 'Yes' on their 'CT/X-Ray Details' eCRF.

Incomplete data will be presented in a table detailing the number and percentage of participants with incomplete data who could not be assessed because they are missing the following[8]:

- Temperature,
- Heart rate,
- Respiratory rate,
- Systolic blood pressure,
- Oxygen saturation on room air,
- Oxygen saturation on room air (pre-HAP state)*,
- Ability to eat and drink independently,
- Ability to eat and drink independently (pre-HAP state)*,
- Cognitive function,
- Radiological scan result^,
- CAP-Sym,
- CAP-Sym (baseline not completed within 3 days).

* For some participants this data may be not known at admission. The clinical cure criteria for oxygen saturation (≥ 90% on room air or return to pre-HAP state) and ability to eat and drink independently (yes or return to pre-HAP state) are not fully dependent on the pre-HAP state being collected so participants may still meet the clinical cure criteria without admission data for oxygen saturation and ability to eat and drink independently.

^ Further imaging and findings are collected on the Day 90 visit eCRF. Only imaging carried out up to day 10 is applicable to this outcome. If the participant does not have a Day 90 eCRF or their eCRF indicates that they had further imaging up to day 10 and the finding(s) are not recorded on the eCRF they will be considered as missing their radiological scan result for that day.

---

[8] Only the participant's least missing assessment up to day 10, i.e. the day that has the least components missing, will be included in this table. Participants may have more than one sign/ symptom/ radiological sign missing.

**Questionnaires**

For each questionnaire and timepoint, the number of questionnaires expected/returned/completed and the return/completion rates will be presented (completion rates will be presented split by item for the EQ-5D-5L and Post-discharge indirect cost survey). Additionally, for the CAP-Sym questionnaire the number of missing items (on returned questionnaires) for each timepoint will be presented using the following summaries: mean, SD, median and IQR.

###### 25.2.3.7 Secondary outcome 7: Eligibility, recruitment and consent rates

###### 25.2.3.7.1 Derivation

**Rate of recruitment**

The overall recruitment rate/month (R) will be calculated as follows:

$$R = \frac{Total\ number\ of\ patients\ recruited}{Total\ number\ of\ months\ with\ a\ site\ open\ to\ recruitment*}.$$

For each site, the recruitment rate/month ($R_{Site}$) will be calculated as follows:

$$R_{Site} = \frac{Total\ number\ of\ patients\ recruited\ at\ site}{Total\ number\ of\ months\ site\ was\ open\ to\ recruitment*}.$$

* The exact number of months sites were open for will be used. Sites will be classed as open and active once they have completed the study greenlight checklist.

**Proportion screened that meet eligibility criteria**

For stage 1, the number of patients screened will be the estimated total number of HAP cases during the recruiting period for all recruiting centres. This will be calculated using data provided by AQUA. The number of patients that meet the eligibility criteria will be all participants who have an 'Eligibility Stage 1' eCRF with 'Were all inclusion/exclusion criteria passed?' answered with 'Yes'.

For stage 2, the number of patients screened will be the total number randomised to stage 1. The number of patients that meet the eligibility criteria will be all participants who have an 'Eligibility Stage 2' eCRF with 'Were all inclusion/exclusion criteria passed?' answered with 'Yes'.

For each stage, the proportion screened that meet the eligibility criteria will be calculated by dividing the number of patients eligible by the number of patients screened.

**Proportion eligible that consent and where they present**

The number of patients eligible will be all patients who have an 'Eligibility Stage 1' eCRF with 'Were all inclusion/exclusion criteria passed?' answered with 'Yes'. The number of patients who consented will be all patients who have an 'Informed Consent' eCRF with 'Was informed consent obtained from the patient or a personal nominated consultee?' answered with 'Yes'.

The proportion eligible that consent will be calculated by dividing the number of patients who consented by the number of eligible patients. Proportions will also be calculated for each ward type patients where on as inpatients at the time of their stage 1 randomisation (medical, surgical, A&E, acute medical unit, other, not known). The ward type is collected on the 'Day 0 Baseline' eCRF.

###### 25.2.3.7.2 Analysis

**Rate of recruitment**

- The total number of patients recruited, the total number of months with a site open to recruitment and the recruitment rate/month will be presented.
- For each site, the total number of patients recruited, the total numbers of months the site was open and the recruitment rate/month will be presented.

**Proportion screened that meet eligibility criteria**

For each stage, the number of patients screened/eligible and the proportion will be presented.

**Proportion eligible that consent and where they present**

For each ward type and overall, the number of patients eligible/consented and the proportion will be presented.

##### 25.2.3.8 Secondary outcome 8: Rates of successful follow-up

###### 25.2.3.8.1 Derivation

For each treatment arm, the number of participants consented and randomised will be the total number of participants randomised to that treatment arm who provided consent.

**Proportion consented and randomised that complete the full study pathway as per protocol**

Participants who are consented and randomised will be classed as completing the full study pathway if they have completed inpatient data and have their day 28 and 90 visits (completed within the visit window), i.e. if they have completed the following:

| | **eCRFs** | **Visit window** |
|---|---|---|

| | | |
|---|---|---|
| Inpatient data | | |
| Day 28 | Day 28 questionnaires<br>Day 28 Visit | Day 28 (+/- 7 days) |
| Day 90 | Day 90 questionnaires<br>Day 90 Visit | Day 90 (+/- 14 days) |

For each treatment arm, the proportion consented and randomised that complete the study pathway will be calculated by dividing the number of participants who completed the study pathway by the number of participants consented and randomised.

**Proportion consented and randomised that withdraw from study intervention or follow up**

A consented and randomised participant is classed as a withdrawal from follow up if they have a 'Withdrawal from Study' eCRF and if the participant made the decision to withdraw, their withdrawal type is not 'Withdrawal from research samples only'. While the nature of the intervention(s) mean that they are delivered at a single time point, if a participant does not receive their allocated intervention, e.g. if they were allocated to FAPP in stage 2 but their 'FAPP Report' eCRF has the question 'Was the FAPP carried out?' answered with 'No', they will be classed as a withdrawal from study intervention and similarly for CT and CXR. For each treatment arm, the number of participants who withdrew from study intervention or follow-up will be all participants who are classed as withdrawals from follow up and/or the intervention.

The proportion consented and randomised that withdraw from study intervention or follow up will be calculated by dividing the number of participants who withdrew from study intervention or follow-up by the number of participants consented and randomised.

The proportion consented and randomised that withdraw from study intervention or follow up (not due to death) will be calculated by dividing the number of participants who withdrew from study intervention or follow-up (excluding those whose reason for withdrawal was 'Death' and did not withdraw from intervention) by the number of participants consented and randomised.

**25.2.3.8.2 Analysis**

**Proportion consented and randomised that complete the full study pathway as per protocol**

For each treatment arm, the number of participants consented and randomised/completed the study pathway (as per the protocol) and the proportion will be presented.

**Proportion consented and randomised that withdraw from study intervention or follow up**

For each treatment arm, the following will be presented:

- The number of participants consented and randomised/withdrew from study intervention or follow up and the proportion,
- The number of participants consented and randomised/withdrew from study intervention or follow up (not due to death) and the proportion.

###### 25.2.3.9 Secondary outcome 9: Summary statistics relating to antibiotic use in the pilot study with a comparison between the DTRs

###### 25.2.3.9.1 Derivation

For each participant, the antibiotic usage in DDDs (calculated using the method described in section 25.2.3.2.1) for each antibiotic they have recorded on the 'Antibiotic Usage' eCRF will be added together to compute their total antibiotic usage in DDDs.

###### 25.2.3.9.2 Analysis

For each DTR, total antibiotic usage (in DDDs) will be presented using the following summaries: mean, SD, median and IQR. Additionally, boxplots summarising total antibiotic usage (in DDDs) will be presented for each DTR.

###### 25.2.3.10 Qualitative outcomes

The qualitative outcomes listed in section 15.2 will not be reported in the statistical analysis report.

###### 25.2.3.11 Costing analysis outcome

The costing analysis outcome listed in section 15.2 will not be reported in the statistical analysis report.

### 26 Safety Evaluations

Not applicable. Since HAP-FAST only incorporates well-established and non-invasive diagnostic investigations that would normally be carried out as standard of care, safety events will not be recorded as part of this study.

### 27 Additional Analyses

Depending on the hypotheses generated from this study, some additional analyses may be carried out post-hoc. Any extra analyses will be clearly marked as post-hoc.

### 28 Document History

| Statistical Analysis Plan Version | Protocol Version | Section number(s) changed | Description of changes | Justification for changes | Date Implemented |
|---|---|---|---|---|---|

### 29 References

#### 29.1 Non-standard statistical methods

Not applicable.

#### 29.2 Data Management Plan

HAP-FAST Data Management Plan V1.0 (dated 25/01/2023, effective 27/02/2023)

#### 29.3 Trial Master File and Trial Statistical File

HAP-FAST Trial Master File

HAP-FAST Trial Statistical File

#### 29.4 Other Standard Operating Procedures to be adhered to

LCTC_ST001: Sample Size Calculation

LCTC_ST002: Generation of a Random Allocation

LCTC_ST003: Production of Statistical Analysis Plans and Reports

LCTC_ST004: Quality control of Statistical Programming

LCTC_ST005: Statistical Programmed Data Checks

LCTC_ST006: Creating and Maintaining a Trial Statistical File

#### 29.5 Other references

[1] *Woodhead M. Pneumonia: Diagnosis and management of community and hospital-acquired pneumonia in adults. 2014.*

[2] *Burton LA, Price R, Barr KE, McAuley SM, Allen JB, Clinton AM, et al. Hospital-acquired pneumonia incidence and diagnosis in older patients. Age and Ageing. 2015;45(1):171-4.*

[3] *Felton T, Wootton D. Antimicrobial prescribing for hospital-acquired pneumonia. Br J Hosp Med (Lond). 2020;81(1):1-3.*

[4] *Vock DM, Almirall D. Sequential Multiple Assignment Randomized Trial (SMART). Wiley StatsRef: Statistics Reference Online. p. 1-11.*

[5] *Moher D, Hopewell S, Schulz KF, Montori V, Gøtzsche PC, Devereaux PJ, et al. CONSORT 2010 Explanation and Elaboration: updated guidelines for reporting parallel group randomised trials. BMJ. 2010;340:c869.*

[6] *Eldridge SM, Chan CL, Campbell MJ, et al. CONSORT 2010 statement: extension to randomised pilot and feasibility trials. Pilot and Feasibility Studies. 2016;2:64.*

[7] *Herdman M, Gudex C, Lloyd A, Janssen M, Kind P, Parkin D, et al. Development and preliminary testing of the new five-level version of EQ-5D (EQ-5D-5L). Qual Life Res. 2011;20(10):1727-36.*

[8] *Foundation ER. EQ-5D-5L user guide. EuroQol Research Foundation Rotterdam, The Netherlands; 2019.*
