## Supplementary material for "Low-dose computed tomography and rapid molecular sputum testing in adults with suspected non-ventilator-associated hospital-acquired pneumonia: the HAP-FAST pilot sequential multiple assignment randomised trial with embedded qualitative and costing studies": Statistical Analysis Report

**Feasibility study of the clinical and cost-effectiveness of contemporary diagnostics for patients with suspected Hospital-Acquired Pneumonia (HAP).**

**Trial registration No: NCT05483309**

### Final Statistical Analysis Report

### Version 1.0 30/04/2025

| | ORIGINATED BY | QC PERFORMED BY | APPROVED BY |
|---|---|---|---|
| **Name** | Ian Powell | Liam Whittle | Dr Ashley Jones |
| **Title** | Trial Statistician | Trial Statistician | Head of Statistics |
| **Date** | 30/04/2025 | | |
| **Protocol Version and Date** | V4.0 12/08/2024 | | |
| **Statistical Analysis Plan Version and Date** | V1.0 01/11/2024 | | |
| **Report Shell Version and Date** | V1.0 04/11/2024 | | |

14 Lay Summary of Study Results ... 60
14.1 Trial delivery and processes ... 60
14.2 Clinical effects (outcomes) of the trial ... 60
14.2.1 Cure ... 60
14.2.2 Antibiotic use ... 61
14.2.3 Quality of life ... 61
14.2.4 Length of hospital stay ... 62
14.2.5 Deaths ... 62
14.3 Feasibility of a future trial or trials ... 62
15 Mapping between report shell and SAP ... 63
16 Version history ... 64

### 2 List of Tables and Figures

Table 4-1 Screening summary by centre (stage 1) ... 9
Table 4-2 Calculating percentages for Table 4-1 ... 9
Table 4-3 Screening summary by centre (stage 2) ... 10
Table 4-4 Calculating percentages for Table 4-3 ... 10
Table 4-5 Reasons for ineligibility (stage 1) ... 11
Table 4-6 Reasons for ineligibility (stage 2) ... 11
Table 4-7 Reasons for not providing consent (randomised to stage 1 only) ... 12
Table 4-8 Reasons for not providing consent (randomised to stages 1 and 2) ... 13
Table 4-9 Randomisation details for recruiting centres ... 14
Table 4-10 Calculating percentages for Table 4-9 ... 14
Table 5-1 Output from randomisation checks ... 16
Table 7-1 Reasons for withdrawal from follow-up, among patients who consented. ... 18
Table 7-2 Deaths (all causes), all patients including those who died without providing consent ... 19
Table 8-1 Demographic details ... 21
Table 8-2 Vital signs (at time of suspected HAP) ... 22
Table 8-3 Inpatient details and HAP signs & symptoms ... 24
Table 8-4 Concomitant medications and immunosuppressant medication use (at time of suspected HAP) ... 25
Table 8-5 Admission details ... 26
Table 9-1 Data sets analysed ... 33
Table 10-1 Protocol deviations ... 33
Table 11-1 Summary statistics – time from stage 1 randomisation to scan and to scan report ... 34
Table 11-2 Reasons for not receiving allocated intervention (CT scan or chest X-ray) ... 34
Table 11-3 Reasons for not receiving FAPP (if allocated to FAPP) ... 34
Table 12-1 Summary statistics of time to clinical cure. ... 35
Table 12-2 Number and percentage of participants experiencing clinical cure who went onto fail clinical cure at a later visit (days 1-10) ... 35
Table 12-3 Summary statistics – percentage change in CAP-Sym score from baseline to time of clinical cure ... 35
Table 12-4 Number and percentage of participants who could not be assessed for clinical cure and failed clinical cure ... 36
Table 12-5 Kaplan-Meier estimates and Cox Proportional Hazards models estimates – time to clinical cure ... 36
Table 12-6 Summary statistics – antibiotic usage in DDD for the HAP episode ... 39

### 3 CONSORT flow diagram

Figure 3-1 CONSORT flow diagram (enrolment and allocation)

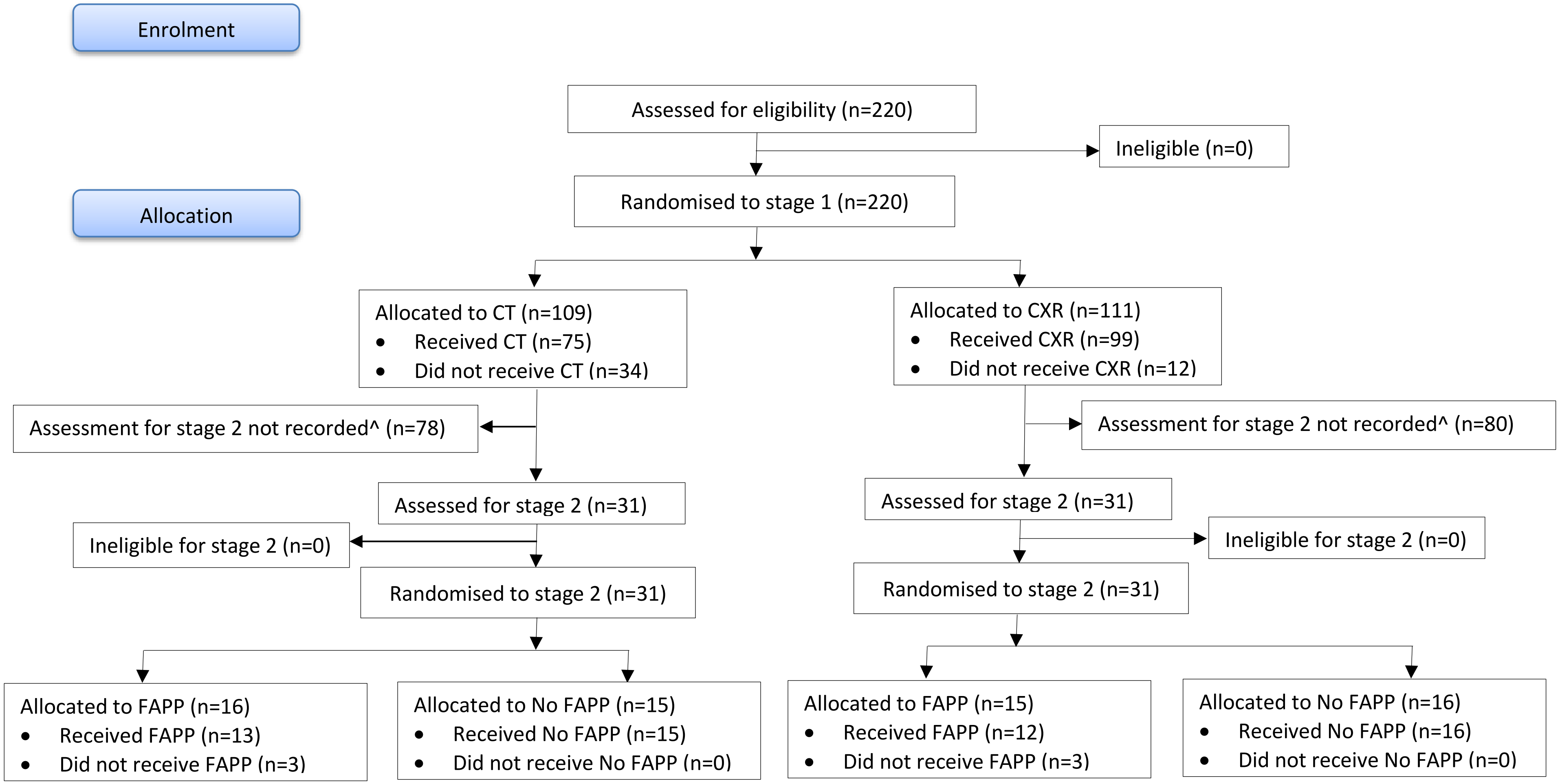

*Created using: X:\HAP-FAST\Statistical Analysis - Blinded\Final Analysis\Version 1.0\Programs\Section03_CONSORT.sas*

^ 150 patients whose assessment for eligibility for stage 2 was not recorded, were not able to produce a sputum sample before their second dose of antibiotic and were therefore not eligible for stage 2.

Figure 3-2 CONSORT flow diagram (consent, follow-up and analysis)

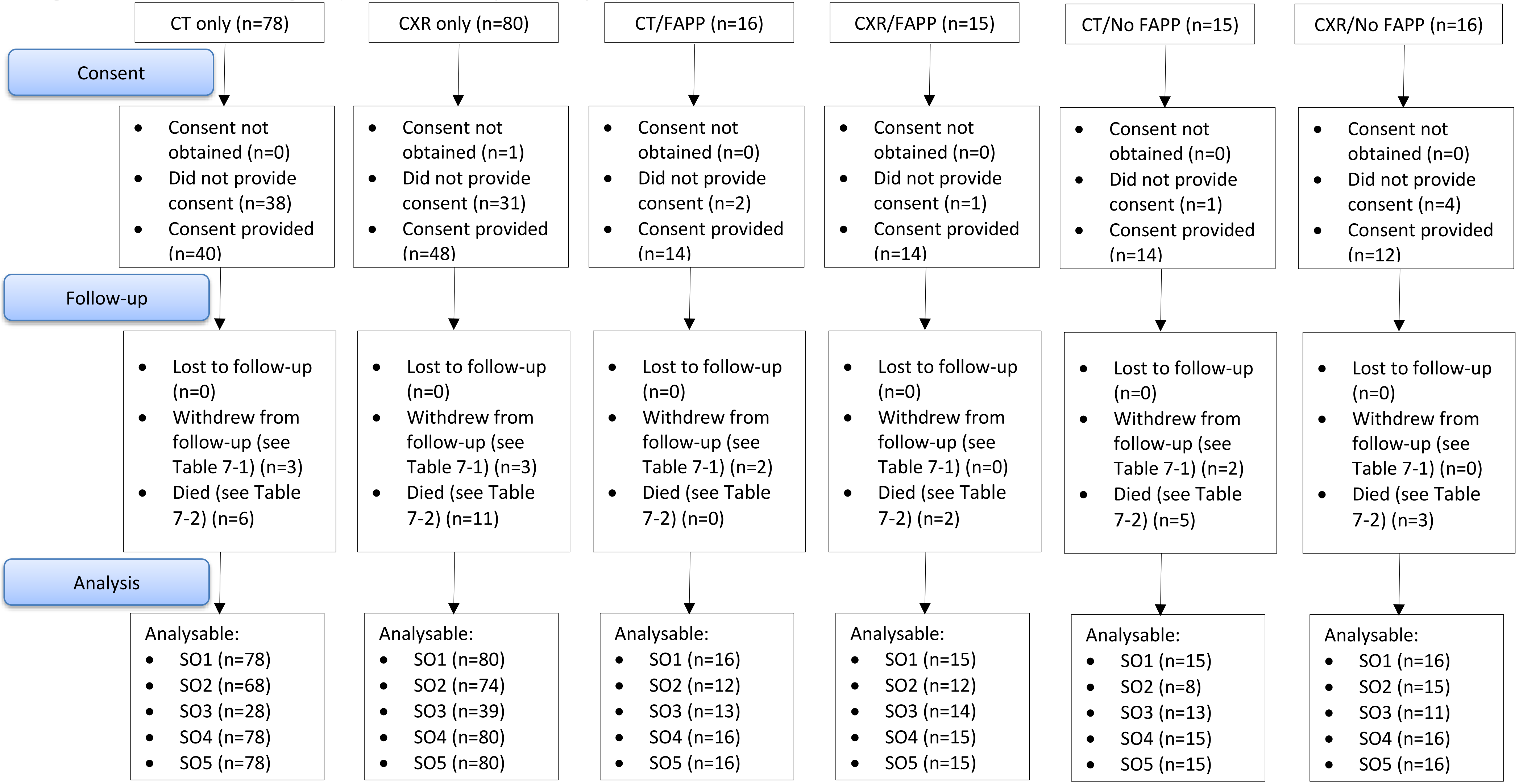

*Created using: X:\HAP-FAST\Statistical Analysis - Blinded\Final Analysis\Version 1.0\Programs\Section03_CONSORT.sas*

SO1 = Time to clinical cure (all patients randomised to CT scan/ X-ray are analysable for this outcome)
SO2 = Antibiotic usage (all patients randomised to stage 1 are analysable for this outcome)
SO3 = EQ-5D-5L (all patients who consented and answered at least one question on the EQ-5D-5L, at any timepoint – baseline, day 10, day 28 or day 90, are analysable this outcome)
SO4 = Length of hospital stay (all patients randomised to stage 1 are analysable for this outcome)
SO5 = Mortality (all patients randomised to stage 1 are analysable for this outcome)

### 4 Recruitment

#### 4.1 Screening summary

Due to the nature of the study and the urgent need to treat, standard screening logs were not maintained for HAP-FAST. As soon as a patient was identified as having suspected HAP, they were assessed for eligibility and included in the study (if eligible). For participants who were assessed for eligibility but not randomised at stage one, ineligibility reason(s) were recorded by the online randomisation system. All screening data available is presented in the following tables.

Table 4-1 Screening summary by centre (stage 1)

| Centre (code) | No. screened | Eligible | Ineligible | Randomised | Assessed for stage 2 | Assessment for stage 2 not recorded | Assessment for stage 2 not recorded | | |
|---|---|---|---|---|---|---|---|---|---|
| | | | | | | | Consent provided | Consent not obtained[1] | Did not provide consent |
| | *[N]* N | *[a]* n (%) | *[b]* n (%) | *[c]* n (%) | *[d]* n (%) | *[e]* n (%) | *[f]* n (%) | *[g]* n (%) | *[h]* n (%) |
| **Overall** | **220** | **220 (100.0)** | **0 (0.0)** | **220 (100.0)** | **62 (28.2)** | **158 (71.8)** | **88 (55.7)** | **1 (0.6)** | **69 (43.7)** |
| Aintree (01) | 117 | 117 (100.0) | 0 (0.0) | 117 (100.0) | 40 (34.2) | 77 (65.8) | 42 (54.5) | 1 (1.3) | 34 (44.2) |
| Royal Liverpool (02) | 34 | 34 (100.0) | 0 (0.0) | 34 (100.0) | 7 (20.6) | 27 (79.4) | 22 (81.5) | 0 (0.0) | 5 (18.5) |
| Royal Preston (03) | 32 | 32 (100.0) | 0 (0.0) | 32 (100.0) | 13 (40.6) | 19 (59.4) | 14 (73.7) | 0 (0.0) | 5 (26.3) |
| Wythenshawe (04) | 37 | 37 (100.0) | 0 (0.0) | 37 (100.0) | 2 (5.4) | 35 (94.6) | 10 (28.6) | 0 (0.0) | 25 (71.4) |

*Created using: X:\HAP-FAST\Statistical Analysis - Blinded\Final Analysis\Version 1.0\Programs\Section04_ScreeningRandomisationRecruitment.sas*

Table 4-2 Calculating percentages for Table 4-1

| Percentages | Equation |
|---|---|
| Percent eligible | a/N |
| Percent ineligible | b/N |
| Percent randomised | c/a |
| Percent assessed for stage 2 | d/c |
| Percent assessment for stage 2 not recorded | e/c |
| Assessment for stage 2 not recorded: Percent consent provided | f/e |
| Assessment for stage 2 not recorded: Percent consent not obtained | g/e |
| Assessment for stage 2 not recorded: Percent did not provide consent | h/e |

[1] For some participants consent may not be able to be obtained, e.g. if a patient is lost to follow-up before providing consent.

Table 4-3 Screening summary by centre (stage 2)

| Centre (code) | No. assessed for stage 2 | Eligible | Ineligible | Ineligible for stage 2 | | | Randomised | Randomised to stage 2 | | |
|---|---|---|---|---|---|---|---|---|---|---|
| | | | | Consent provided | Consent not obtained[2] | Did not provide consent | | Consent provided | Consent not obtained[2] | Did not provide consent |
| | *[N]* N | *[a]* n (%) | *[b]* n (%) | *[c]* n (%) | *[d]* n (%) | *[e]* n (%) | *[f]* n (%) | *[g]* n (%) | *[h]* n (%) | *[i]* n (%) |
| **Overall** | **62** | **62 (100.0)** | **0 (0.0)** | **0 (0.0)** | **0 (0.0)** | **0 (0.0)** | **62 (100.0)** | **54 (87.1)** | **0 (0.0)** | **8 (12.9)** |
| Aintree (01) | 40 | 40 (100.0) | 0 (0.0) | 0 (0.0) | 0 (0.0) | 0 (0.0) | 40 (100.0) | 35 (87.5) | 0 (0.0) | 5 (12.5) |
| Royal Liverpool (02) | 7 | 7 (100.0) | 0 (0.0) | 0 (0.0) | 0 (0.0) | 0 (0.0) | 7 (100.0) | 6 (85.7) | 0 (0.0) | 1 (14.3) |
| Royal Preston (03) | 13 | 13 (100.0) | 0 (0.0) | 0 (0.0) | 0 (0.0) | 0 (0.0) | 13 (100.0) | 12 (92.3) | 0 (0.0) | 1 (7.7) |
| Wythenshawe (04) | 2 | 2 (100.0) | 0 (0.0) | 0 (0.0) | 0 (0.0) | 0 (0.0) | 2 (100.0) | 1 (50.0) | 0 (0.0) | 1 (50.0) |

*Created using: X:\HAP-FAST\Statistical Analysis - Blinded\Final Analysis\Version 1.0\Programs\Section04_ScreeningRandomisationRecruitment.sas*

Table 4-4 Calculating percentages for Table 4-3

| **Percentages** | **Equation** |
|---|---|
| Percent eligible | a/N |
| Percent ineligible | b/N |
| Ineligible for stage 2: Percent consent provided | c/b |
| Ineligible for stage 2: Percent consent not obtained | d/b |
| Ineligible for stage 2: Percent did not provide consent | e/b |
| Percent randomised | f/a |
| Randomised to stage 2: Percent consent provided | g/f |
| Randomised to stage 2: Percent consent not obtained | h/f |
| Randomised to stage 2: Percent did not provide consent | i/f |

[2] For some participants consent may not be able to be obtained, e.g. if a patient is lost to follow-up before providing consent.

Table 4-5 Reasons for ineligibility (stage 1)

*Table 4-5 cannot be produced because no patients were screened as ineligible.*
*Created using: X:\HAP-FAST\Statistical Analysis - Blinded\Final Analysis\Version 1.0\Programs\Section04_ScreeningRandomisationRecruitment.sas*

[a] Patients may be ineligible for one or more reasons. Percentages are based on the number of ineligible patients for stage 1 at each centre.
[b] Reasons for ineligibility:

R1= Aged less than 18 years
R2= Does not have suspected HAP
R3= Already received a chest X-ray to confirm suspected HAP diagnosis
R4= Diagnosis or suspected diagnosis of ventilator acquired pneumonia
R5= Intention to palliate rather than cure
R6= Interventions cannot be completed before administration of second antibiotic dose
R7= Cannot be randomised to low-dose, non-contrast CT scan on clinical grounds e.g. strong suspicion of PE
R8= Pregnant
R9= Previous study participation (patients with second of third episodes of HAP will not be re-recruited)

Table 4-6 Reasons for ineligibility (stage 2)

*Table 4-6 cannot be produced because no patients were screened as ineligible.*
*Created using: X:\HAP-FAST\Statistical Analysis - Blinded\Final Analysis\Version 1.0\Programs\Section04_ScreeningRandomisationRecruitment.sas*

[a] Patients may be ineligible for one or more reasons. Percentages are based on the number of ineligible patients for stage 2 at each centre.
[b] Reasons for ineligibility:

R1= Following the CXR or CT the clinician does not intend to treat with antibiotics for either HAP or a hospital acquired RTI
R2= A sputum sample cannot be obtained before the second dose of antibiotic
R3= Following the CXR or CT the clinician decides not to treat with antibiotics for either HAP or hospital acquired RTI

Table 4-7 Reasons for not providing consent (randomised to stage 1 only)

| Site (code) | No. of patients who did not provide consent | Reasons for not providing consent [a, b], n (%) | | | | | | | | | | | | | |
|---|---|---|---|---|---|---|---|---|---|---|---|---|---|---|---|
| | | R1 | R2 | R3 | R4 | R5 | R6 | R7 | R8 | R9 | R10 | R11 | R12 | R13 | R14 |
| **Overall** | **69 (100.0)** | **42 (60.9)** | **6 (8.7)** | **4 (5.8)** | **4 (5.8)** | **3 (4.3)** | **2 (2.9)** | **1 (1.4)** | **1 (1.4)** | **1 (1.4)** | **1 (1.4)** | **1 (1.4)** | **1 (1.4)** | **1 (1.4)** | **1 (1.4)** |
| Aintree (01) | 34 (100.0) | 22 (64.7) | 4 (11.8) | 3 (8.8) | 0 (0.0) | 1 (2.9) | 0 (0.0) | 1 (2.9) | 1 (2.9) | 1 (2.9) | 1 (2.9) | 0 (0.0) | 0 (0.0) | 0 (0.0) | 0 (0.0) |
| Royal Liverpool (02) | 5 (100.0) | 1 (20.0) | 2 (40.0) | 0 (0.0) | 0 (0.0) | 1 (20.0) | 0 (0.0) | 0 (0.0) | 0 (0.0) | 0 (0.0) | 0 (0.0) | 1 (20.0) | 0 (0.0) | 0 (0.0) | 0 (0.0) |
| Royal Preston (03) | 5 (100.0) | 1 (20.0) | 0 (0.0) | 0 (0.0) | 1 (20.0) | 1 (20.0) | 0 (0.0) | 0 (0.0) | 0 (0.0) | 0 (0.0) | 0 (0.0) | 0 (0.0) | 1 (20.0) | 1 (20.0) | 0 (0.0) |
| Wythenshawe (04) | 25 (100.0) | 18 (72.0) | 0 (0.0) | 1 (4.0) | 3 (12.0) | 0 (0.0) | 2 (8.0) | 0 (0.0) | 0 (0.0) | 0 (0.0) | 0 (0.0) | 0 (0.0) | 0 (0.0) | 0 (0.0) | 1 (4.0) |

*Created using: X:\HAP-FAST\Statistical Analysis - Blinded\Final Analysis\Version 1.0\Programs\Section04_ScreeningRandomisationRecruitment.sas*

[a] Percentages are based on the number of patients who were only randomised to stage 1 and did not provide consent at each site.
[b] Reasons for not providing consent:

R1 = Declined consent - no reason given
R2 = Not eligible
R3 = Died before providing consent
R4 = Unable to gain consent
R5 = Did not want to participate
R6 = Lost to follow-up before providing consent
R7 = Family declined consent - no reason given
R8 = Family declined consent due to state of patient
R9 = Not being treated for HAP - patient did not feel answers will be beneficial to study
R10 = Felt too unwell
R11 = Patient disapproves of research generally
R12 = Family declined consent due to dementia
R13 = Does not have the time and energy to read and understand the purpose of the study
R14 = Decided not to go ahead with research due to diagnosis of new cancer

Table 4-8 Reasons for not providing consent (randomised to stages 1 and 2)

| **Site (code)** | **No. of patients who did not provide consent** | **Reasons for not providing consent [a, b], n (%)** | | | | |
|---|---|---|---|---|---|---|
| | | **R1** | **R2** | **R3** | **R4** | **R5** |
| **Overall** | **8 (100.0)** | **3 (37.5)** | **2 (25.0)** | **1 (12.5)** | **1 (12.5)** | **1 (12.5)** |
| Aintree (01) | 5 (100.0) | 3 (60.0) | 1 (20.0) | 1 (20.0) | 0 (0.0) | 0 (0.0) |
| Royal Liverpool (02) | 1 (100.0) | 0 (0.0) | 1 (100.0) | 0 (0.0) | 0 (0.0) | 0 (0.0) |
| Royal Preston (03) | 1 (100.0) | 0 (0.0) | 0 (0.0) | 0 (0.0) | 1 (100.0) | 0 (0.0) |
| Wythenshawe (04) | 1 (100.0) | 0 (0.0) | 0 (0.0) | 0 (0.0) | 0 (0.0) | 1 (100.0) |

*Created using: X:\HAP-FAST\Statistical Analysis - Blinded\Final Analysis\Version 1.0\Programs\Section04_ScreeningRandomisationRecruitment.sas*

[a] Percentages are based on the number of patients who were randomised to stages 1 and 2 and did not provide consent at each site.
[b] Reasons for not providing consent:

R1 = Declined consent - no reason given
R2 = Did not want to participate
R3 = Not eligible
R4 = Family declined consent - feels trial procedures can put a disabled patient in too much discomfort
R5 = Unable to gain consent

#### 4.2 Randomisation

##### 4.2.1 Randomisation

Table 4-9 Randomisation details for recruiting centres

| Centre (code) | Date of centre opening | Date of first stage 1 randomisation | Date of most recent stage 1 randomisation | Randomised | | Did not provide consent |
|---|---|---|---|---|---|---|
| | | | | Stage 1 [N] n | Stage 2 [a] n (%) | [b] n (%) |
| Aintree (01) | 2023-06-07 | 2023-06-13 | 2024-03-12 | 117 | 40 (34.2) | 39 (33.3) |
| Royal Liverpool (02) | 2023-06-07 | 2023-06-15 | 2024-03-11 | 34 | 7 (20.6) | 6 (17.6) |
| Royal Preston (03) | 2023-06-14 | 2023-07-05 | 2024-02-29 | 32 | 13 (40.6) | 6 (18.8) |
| Wythenshawe (04) | 2023-06-23 | 2023-06-28 | 2024-03-06 | 37 | 2 (5.4) | 26 (70.3) |

*Created using: X:\HAP-FAST\Statistical Analysis - Blinded\Final Analysis\Version 1.0\Programs\Section04_ScreeningRandomisationRecruitment.sas*

Table 4-10 Calculating percentages for Table 4-9

| Percentages | Equation |
|---|---|
| Percent randomised to stage 2 | a/N |
| Percent did not provide consent | b/N |

#### 4.3 Recruitment graph

##### 4.3.1 Recruitment graph

Figure 4-1 Recruitment graph

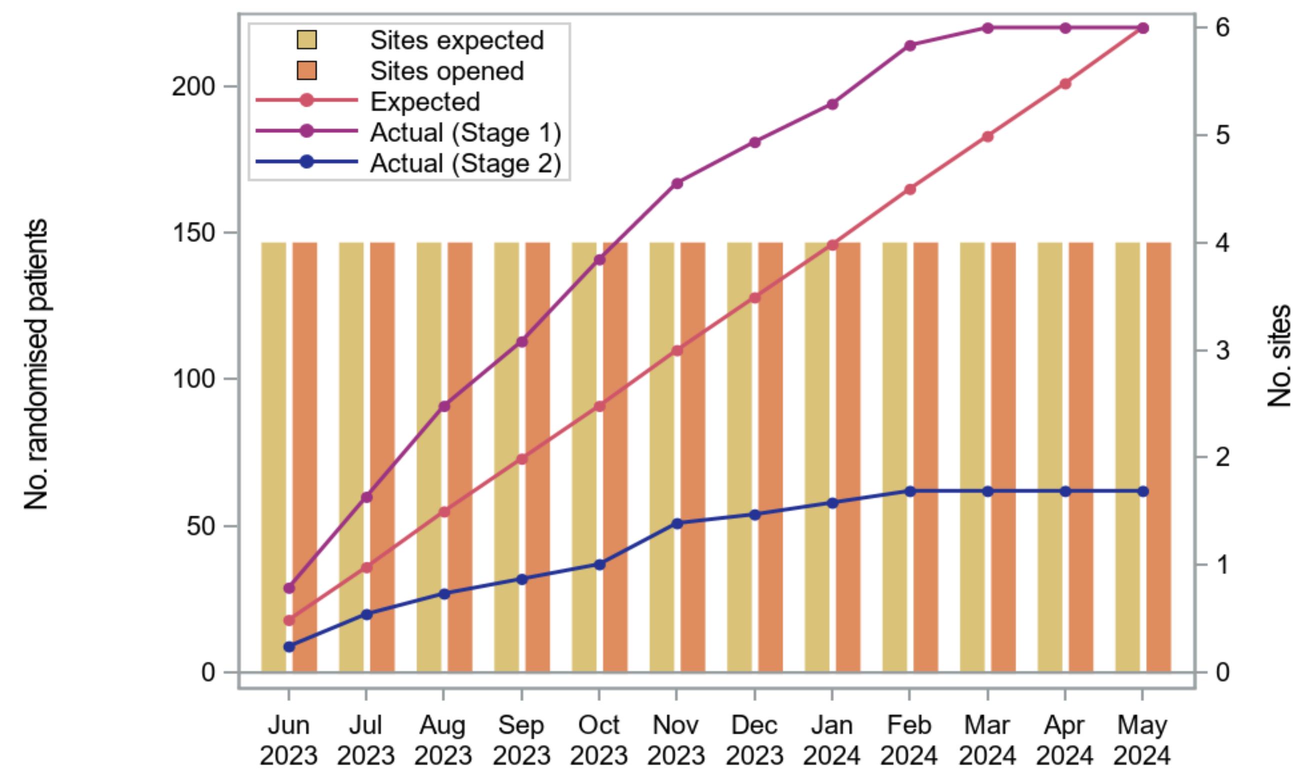

| | Jun 2023 | Jul 2023 | Aug 2023 | Sep 2023 | Oct 2023 | Nov 2023 | Dec 2023 | Jan 2024 | Feb 2024 | Mar 2024 | Apr 2024 | May 2024 |
|---|---|---|---|---|---|---|---|---|---|---|---|---|
| Sites expected | 4 | 4 | 4 | 4 | 4 | 4 | 4 | 4 | 4 | 4 | 4 | 4 |
| Sites opened | 4 | 4 | 4 | 4 | 4 | 4 | 4 | 4 | 4 | 4 | 4 | 4 |
| Expected | 18 | 36 | 55 | 73 | 91 | 110 | 128 | 146 | 165 | 183 | 201 | 220 |
| Actual (Stage 1) | 29 | 60 | 91 | 113 | 141 | 167 | 181 | 194 | 214 | 220 | 220 | 220 |
| Actual (Stage 2) | 9 | 20 | 27 | 32 | 37 | 51 | 54 | 58 | 62 | 62 | 62 | 62 |

*Created using: X:\HAP-FAST\Statistical Analysis - Blinded\Final Analysis\Version 1.0\Programs\Section04_ScreeningRandomisationRecruitment.sas*

### 5 Randomisation

#### 5.1 Randomisation procedure

There were two stages of randomisation in the pilot study. Both used a secure (24-hour) web-based randomisation system controlled centrally by the LCTC. Randomisation lists were generated with random variable block sizes and stratified by site.

**Randomisation 1: Choice of imaging**
Participants were randomised to undergo either CT scan (CT) or chest X-ray (CXR) in a ratio of 1:1.

**Randomisation 2: FAPP or No FAPP**
Once imaging had been completed, and a clinical judgement made, participants who were:

- Treated as HAP or a hospital acquired RTI, and
- Able to produce a sputum sample

were randomised to FAPP or No FAPP (in a ratio of 1:1).

#### 5.2 Randomisation checks

Table 5-1 Output from randomisation checks

| Check | Pass/ fail |
|---|---|
| No missing randomisation numbers | Pass |
| Randomisation numbers are unique (no duplication) | Fail[3] |
| Randomisation numbers are sequential by date randomised | Fail[4] |
| Allocations provided to site by web-based system match those within the lists provided by the randomising statistician | Pass |
| Balance in patient numbers across interventions at each stage of randomisation (CT vs X-ray) and (FAPP vs no FAPP) | Pass |

*Created using: T:\Ctrcis\Statistical Analysis\HAP-FAST\Randomisation\Randomisation output\Randomisation checking\7_FinalCheck_20250124\ HAP-FAST_RandomisationChecks_20250129.do*

#### 5.3 Randomisation problems

HAP-FAST had three randomisation problems:

1. Participant 01020 was entered into the randomisation system under the wrong site (Aintree University Hospital rather than Liverpool Royal). It was agreed that they would keep their allocation as allocated at randomisation but would be reassigned to the correct site in REDCap.
2. Participant 03027 had a duplicate stage 2 randomisation. Both allocations are the same and therefore there is no concern that the participant received the incorrect treatment.
3. Participant 02097 was randomised in the test system. In order to maintain their allocation in the live system a matching allocation from the end of the list for their site was selected. When this participant was entered into the live system, the incorrect site was selected (Aintree University Hospital rather than Liverpool Royal)

[3] Participant 3027 had a duplicate stage 2 randomisation due to site error. Both allocations are the same so there is no concern that the participant received the wrong treatment.
[4] Participant 2097 was initially randomised in the test system in the error. In order to maintain the allocation they had received in the live system they were assigned to an out of sequence allocation at the end of the list for that site

### 6 Serious breaches

HAP-FAST did not have any serious breaches that were reportable to the Research Ethics Committee.

### 7 Completeness of follow-up

77 (35.0%) participants did not provide consent and consent was not able to be obtained from 1 (0.5%) participant.

Table 7-1 Reasons for withdrawal from follow-up, among patients who consented.

| | Stage 1 allocation | Time from stage 1 randomisation to withdrawal (days) | Decision to withdraw made by | Reason for withdrawal | Type of withdrawal |
|---|---|---|---|---|---|
| 1 | CT Scan | 20 | Participant/consultee | No reason given | Withdrawal of consent for ALL further follow up |
| 2 | Chest X-ray | 15 | Participant/consultee | Death | Withdrawal of consent for ALL further follow up |
| 3 | CT Scan | 4 | Clinician | Patient ineligible as had CTPA instead of CT chest | Clinician-initiated withdrawal |
| 4 | CT Scan | 4 | Participant/consultee | No reason given | Withdrawal of consent for ALL further follow up |
| 5 | Chest X-ray | 7 | Participant/consultee | Unhappy after day 3 bloods resulted in lots of bruising and discomfort. Said he was unwell enough and this was an added 'burden'. Consented to all data up to this point to be used. | Withdrawal of consent for ALL further follow up |
| 6 | CT Scan | 0 | Clinician | UN ABLE TO PROGRESS WITH CT SCAN TODAY DUE TO MAJOR TRAUMA PRIORITY | Clinician-initiated withdrawal |
| 7 | Chest X-ray | 105 | Participant/consultee | Participant stated that she has terminal cancer and does not wish to continue with the study as she has other things to worry about. | Withdrawal of consent for ALL further follow up |
| 8 | Chest X-ray | 0 | Clinician | Patient feels the study is too much of commitment for him in terms | Clinician-initiated withdrawal |

| | | | | | |
|---|---|---|---|---|---|
| | | | | of giving bloods and attending outpatient review on the 28th day. | |
| 9 | CT Scan | 5 | Participant/consultee | Patient stated that they don't want to do the regular follow for CAP SYM & day 28 follow up with bloods. | Withdrawal of consent for ALL further follow up |
| 10 | CT Scan | 6 | Clinician | New diagnosis of metastatic lung cancer with rapid deterioration .Patient discharge to Hospice for End of life Care. | Clinician-initiated withdrawal |

*Created using: X:\HAP-FAST\Statistical Analysis - Blinded\Final Analysis\Version 1.0\Programs\Section07_FollowUp.sas*

Table 7-2 Deaths (all causes), all patients including those who died without providing consent

| | **Stage 1 allocation** | **Time from stage 1 randomisation to death (days)** | **Cause of death** |
|---|---|---|---|
| 1 | Chest X-ray | 227 | Pneumonia |
| 2 | Chest X-ray | 15 | significant deterioration overnight, no clear signs of respiratory or cardiac function on inspection. |
| 3 | CT Scan | 26 | pt put onto palliative pathway, significant deterioration |
| 4 | CT Scan | 2 | Not known |
| 5 | Chest X-ray | 4 | Significant deterioration |
| 6 | Chest X-ray | 26 | Not known |
| 7 | Chest X-ray | 137 | Not known |
| 8 | Chest X-ray | 8 | Not known |
| 9 | CT Scan | 39 | Not known |
| 10 | CT Scan | 0 | Significant deterioration |
| 11 | CT Scan | 8 | Significant Deterioration over weekend |
| 12 | CT Scan | 6 | HAP |
| 13 | Chest X-ray | 14 | Patient deteriorated |
| 14 | CT Scan | 36 | Unknown |
| 15 | CT Scan | 16 | Significant deterioration, unresponsive, terminal event. 'Pt unwell, appeared to be approaching end of life' |
| 16 | CT Scan | 13 | Not known |

| | | | |
|---|---|---|---|
| 17 | Chest X-ray | 26 | Not known |
| 18 | Chest X-ray | 105 | Not known |
| 19 | Chest X-ray | 56 | Metastatic renal cancer |
| 20 | Chest X-ray | 39 | Frailty |
| 21 | CT Scan | 30 | Not known |
| 22 | CT Scan | 19 | HAP |
| 23 | CT Scan | 14 | Bronchopneumonia |
| 24 | Chest X-ray | 4 | Pneumonia |
| 25 | Chest X-ray | 4 | CCF |
| 26 | Chest X-ray | 75 | Pulmonary embolism and pneumonia |
| 27 | CT Scan | 37 | Pneumonia |
| 28 | CT Scan | 39 | Not known |
| 29 | Chest X-ray | 30 | Not known |
| 30 | Chest X-ray | 26 | Co-morbidities (AF, SCC, anaemia, CKD, fluid overload, pleural effusion, pulmonary oedema, atelectasis, jaw abscess and osteonecrosis) Sepsis, HAP and End of life |
| 31 | Chest X-ray | 42 | Not known |
| 32 | Chest X-ray | 3 | Not clarified yet. |
| 33 | Chest X-ray | 20 | Cardiac arrest but no clear cause |

*Created using: X:\HAP-FAST\Statistical Analysis - Blinded\Final Analysis\Version 1.0\Programs\Section07_FollowUp.sas*

### 8 Baseline characteristics

Table 8-1 Demographic details

| | | CT | CXR |
|---|---|---|---|
| Randomised: n (%) | Total | 109 (100.0) | 111 (100.0) |
| | Consent provided | 68 (62.4) | 74 (66.7) |
| | Consent not obtained | 0 (0.0) | 1 (0.9) |
| | Did not provide consent | 41 (37.6) | 36 (32.4) |
| Age (years) | N | 109 | 111 |
| | Mean (SD) | 75.5 (11.4) | 75.0 (14.9) |
| | Median (IQR) | 77.0 (70.0, 83.0) | 77.0 (67.0, 85.0) |
| | Range | (36, 97) | (20, 99) |
| | Missing | 17 | 10 |
| Sex: n (%) | Total | 109 (100.0) | 111 (100.0) |
| | Male | 53 (48.6) | 54 (48.6) |
| | Female | 56 (51.4) | 57 (51.4) |
| Height (cm) | N | 109 | 111 |
| | Mean (SD) | 165 (10.8) | 166 (10.2) |
| | Median (IQR) | 164 (157, 173) | 165 (157, 173) |
| | Range | (125, 188) | (147, 198) |
| | Missing | 17 | 12 |
| Weight (kg) | N | 109 | 111 |
| | Mean (SD) | 73.6 (24.9) | 70.1 (17.3) |
| | Median (IQR) | 69.7 (60.5, 82.7) | 69.3 (59.0, 79.4) |
| | Range | (35, 175) | (31, 114) |
| | Missing | 17 | 15 |
| Calculated BMI | N | 109 | 111 |
| | Mean (SD) | 27.1 (8.4) | 25.5 (6.3) |
| | Median (IQR) | 25.4 (21.9, 29.2) | 25.8 (21.3, 28.7) |
| | Range | (13, 57) | (12, 45) |
| | Missing | 17 | 15 |
| Smoking status: n (%) | Total | 109 (100.0) | 111 (100.0) |
| | Current smoker | 9 (8.3) | 19 (17.1) |
| | Ex-smoker | 36 (33.0) | 39 (35.1) |
| | Never smoked | 27 (24.8) | 27 (24.3) |
| | Not known | 20 (18.3) | 15 (13.5) |
| | Missing | 37 (33.9) | 26 (23.4) |

*Created using: X:\HAP-FAST\Statistical Analysis - Blinded\Final Analysis\Version 1.0\Programs\Section08_Baseline.sas*
*'Missing' category includes 'Not collected' and 'Not known' categories*

Table 8-2 Vital signs (at time of suspected HAP)

| | | CT | CXR |
|---|---|---|---|
| Temperature (°C) | N | 109 | 111 |
| | Mean (SD) | 36.8 (0.8) | 36.7 (0.7) |
| | Median (IQR) | 36.6 (36.4, 36.9) | 36.6 (36.2, 37.0) |
| | Range | (35, 40) | (34, 39) |
| | Not collected | 2 | 0 |
| | Not known | 0 | 0 |
| | Missing | 19 | 11 |
| Systolic blood pressure (mmHg) | N | 109 | 111 |
| | Mean (SD) | 124 (19.8) | 126 (25.5) |
| | Median (IQR) | 120 (109, 141) | 122 (109, 142) |
| | Range | (83, 175) | (84, 226) |
| | Not collected | 2 | 0 |
| | Not known | 0 | 0 |
| | Missing | 19 | 11 |
| Diastolic blood pressure (mmHg) | N | 109 | 111 |
| | Mean (SD) | 71.1 (12.2) | 70.7 (14.1) |
| | Median (IQR) | 71.0 (62.0, 79.0) | 72.0 (60.0, 79.0) |
| | Range | (48, 98) | (38, 116) |
| | Not collected | 2 | 0 |
| | Not known | 0 | 0 |
| | Missing | 19 | 12 |
| Pulse rate (beats per minute) | N | 109 | 111 |
| | Mean (SD) | 85.2 (17.7) | 89.2 (20.4) |
| | Median (IQR) | 83.0 (72.0, 96.0) | 88.0 (74.0, 99.0) |
| | Range | (54, 135) | (56, 148) |
| | Not collected | 2 | 0 |
| | Not known | 0 | 1 |
| | Missing | 19 | 12 |
| Respiratory rate (breaths per minute) | N | 109 | 111 |
| | Mean (SD) | 19.2 (3.4) | 20.4 (4.7) |
| | Median (IQR) | 18.0 (17.0, 20.0) | 19.0 (17.0, 24.0) |
| | Range | (12, 32) | (13, 40) |
| | Not collected | 2 | 0 |
| | Not known | 0 | 0 |
| | Missing | 19 | 11 |

| | | | |
|---|---|---|---|
| Oxygen saturation on room air (%) | N | 109 | 111 |
| | Mean (SD) | 94.4 (4.2) | 94.6 (2.7) |
| | Median (IQR) | 96.0 (93.0, 97.0) | 95.0 (93.0, 97.0) |
| | Range | (71, 100) | (88, 100) |
| | Not collected | 2 | 0 |
| | Not known | 15 | 15 |
| | Missing | 34 | 26 |
| NEWS2 Score | N | 109 | 111 |
| | Mean (SD) | 3.0 (2.8) | 3.4 (3.1) |
| | Median (IQR) | 2.0 (1.0, 5.0) | 3.0 (1.0, 5.0) |
| | Range | (0, 12) | (0, 13) |
| | Not collected | 2 | 0 |
| | Not known | 0 | 2 |
| | Missing | 19 | 13 |

*Created using: X:\HAP-FAST\Statistical Analysis - Blinded\Final Analysis\Version 1.0\Programs\Section08_Baseline.sas*
*‘Missing’ category includes ‘Not collected’ and ‘Not known’ categories*

Table 8-3 Inpatient details and HAP signs & symptoms

| | | CT | CXR |
|---|---|---|---|
| Ward type: n (%) | Total | 109 (100.0) | 111 (100.0) |
| | Medical | 48 (44.0) | 48 (43.2) |
| | Surgical | 13 (11.9) | 12 (10.8) |
| | A&E | 28 (25.7) | 34 (30.6) |
| | Acute medical unit | 2 (1.8) | 4 (3.6) |
| | Other | 1 (0.9) | 3 (2.7) |
| | Not known | 0 (0.0) | 0 (0.0) |
| | Missing | 17 (15.6) | 10 (9.0) |
| Clinical symptoms of HAP[5]: n (%) | N | 109 (100.0) | 111 (100.0) |
| | New/worsening shortness of breath | 47 (43.1) | 52 (46.8) |
| | Cough | 65 (59.6) | 67 (60.4) |
| | Fever | 10 (9.2) | 12 (10.8) |
| | Chest pain | 7 (6.4) | 8 (7.2) |
| | New/worsening sputum | 26 (23.9) | 23 (20.7) |
| | Confusion | 16 (14.7) | 9 (8.1) |
| | Other | 14 (12.8) | 17 (15.3) |
| | Any missing | 0 (0.0) | 0 (0.0) |
| Clinical description of respiratory finding of HAP[6]: n (%) | N | 109 (100.0) | 111 (100.0) |
| | New chest crackles | 32 (29.4) | 39 (35.1) |
| | Reduced air entry | 31 (28.4) | 26 (23.4) |
| | Respiratory distress | 20 (18.3) | 28 (25.2) |
| | Other | 25 (22.9) | 20 (18.0) |
| | Any missing | 0 (0.0) | 0 (0.0) |
| 'Medically fit for discharge' prior to HAP suspicion: n (%) | Total | 109 (100.0) | 111 (100.0) |
| | No | 62 (56.9) | 56 (50.5) |
| | Yes | 6 (5.5) | 13 (11.7) |
| | Not known | 24 (22.0) | 31 (27.9) |
| | Missing | 41 (37.6) | 42 (37.8) |

*Created using: X:\HAP-FAST\Statistical Analysis - Blinded\Final Analysis\Version 1.0\Programs\Section08_Baseline.sas*
*'Missing' category includes 'Not collected' and 'Not known' categories*

[5] Participants may have more than one symptom and/or respiratory finding.
[6] Participants may have more than one symptom and/or respiratory finding.

Table 8-4 Concomitant medications and immunosuppressant medication use (at time of suspected HAP)

| | | CT | CXR |
|---|---|---|---|
| Antibiotics taken/received in last 7 days: n (%) | Total | 109 (100.0) | 111 (100.0) |
| | No | 44 (40.4) | 49 (44.1) |
| | Yes | 47 (43.1) | 49 (44.1) |
| | Missing | 18 (16.5) | 13 (11.7) |
| Received immune suppressant mediation (excluding cancer treatment) in the 3 months prior to suspected HAP: n (%) | Total | 109 (100.0) | 111 (100.0) |
| | No | 53 (48.6) | 64 (57.7) |
| | Yes | 4 (3.7) | 1 (0.9) |
| | Not known | 34 (31.2) | 35 (31.5) |
| | Missing | 52 (47.7) | 46 (41.4) |
| Received chemotherapy for a solid organ or haematological cancer in the 3 months prior to suspected HAP: n (%) | Total | 109 (100.0) | 111 (100.0) |
| | No | 60 (55.0) | 65 (58.6) |
| | Yes | 3 (2.8) | 3 (2.7) |
| | Not known | 28 (25.7) | 32 (28.8) |
| | Missing | 46 (42.2) | 43 (38.7) |
| **Concomitant medications taken or received in the last 7 days** | | | |
| Steroids (oral, IV, inhaled): n (%) | Total | 109 (100.0) | 111 (100.0) |
| | Unchecked | 89 (81.7) | 89 (80.2) |
| | Checked | 20 (18.3) | 22 (19.8) |
| Antipsychotics: n (%) | Total | 109 (100.0) | 111 (100.0) |
| | Unchecked | 103 (94.5) | 110 (99.1) |
| | Checked | 6 (5.5) | 1 (0.9) |
| Proton pump inhibitors: n (%) | Total | 109 (100.0) | 111 (100.0) |
| | Unchecked | 89 (81.7) | 84 (75.7) |
| | Checked | 20 (18.3) | 27 (24.3) |
| Statins: n (%) | Total | 109 (100.0) | 111 (100.0) |
| | Unchecked | 91 (83.5) | 87 (78.4) |
| | Checked | 18 (16.5) | 24 (21.6) |
| None of the above concomitant medications: n (%) | Total | 109 (100.0) | 111 (100.0) |
| | Unchecked | 60 (55.0) | 55 (49.5) |
| | Checked | 49 (45.0) | 56 (50.5) |

*Created using: X:\HAP-FAST\Statistical Analysis - Blinded\Final Analysis\Version 1.0\Programs\Section08_Baseline.sas*
*‘Missing’ category includes ‘Not collected’ and ‘Not known’ categories*

Table 8-5 Admission details

| | | CT | CXR |
|---|---|---|---|
| Time from admission to HAP suspected[7] (days) | N | 109 | 111 |
| | Mean (SD) | 17.5 (43.0) | 9.9 (20.8) |
| | Median (IQR) | 7.5 (0.0, 15.9) | 4.9 (0.0, 13.9) |
| | Range | (0, 377) | (0, 186) |
| | Missing | 17 | 12 |
| Reason for admission: n (%) | Total | 109 (100.0) | 111 (100.0) |
| | Medical | 80 (73.4) | 89 (80.2) |
| | Surgical | 12 (11.0) | 11 (9.9) |
| | Missing | 17 (15.6) | 11 (9.9) |
| Had the following conditions at the point of admission: n (%)[8] | N | 109 (100.0) | 111 (100.0) |
| | None of below | 26 (23.9) | 30 (27.0) |
| | Congestive cardiac failure | 12 (11.0) | 11 (9.9) |
| | Dementia | 6 (5.5) | 7 (6.3) |
| | Other form of cognitive impairment | 9 (8.3) | 4 (3.6) |
| | Myocardial Infarction | 6 (5.5) | 4 (3.6) |
| | Chronic pulmonary disease | 30 (27.5) | 36 (32.4) |
| | Rheumatic disease | 1 (0.9) | 3 (2.7) |
| | AIDS (the syndrome not just HIV positive) | 0 (0.0) | 0 (0.0) |
| | Hemiplegia or paraplegia | 0 (0.0) | 1 (0.9) |
| | Renal disease | 13 (11.9) | 21 (18.9) |
| | Liver disease | 1 (0.9) | 1 (0.9) |
| | Diabetes | 23 (21.1) | 10 (9.0) |
| | Any malignancy | 9 (8.3) | 17 (15.3) |
| | Any missing | 0 (0.0) | 0 (0.0) |
| Oxygen saturation on room air at time of admission (%) | N | 109 | 111 |
| | Mean (SD) | 94.7 (4.7) | 94.7 (3.0) |
| | Median (IQR) | 96.0 (93.0, 98.0) | 95.0 (93.0, 97.0) |
| | Range | (71, 100) | (84, 100) |
| | Missing | 26 | 21 |
| Could eat and drink independently prior to admission: n (%) | Total | 109 (100.0) | 111 (100.0) |
| | No | 18 (16.5) | 20 (18.0) |
| | Yes | 50 (45.9) | 64 (57.7) |
| | Not known | 24 (22.0) | 16 (14.4) |
| | Missing | 41 (37.6) | 27 (24.3) |
| Clinical frailty score: n (%) | Total | 109 (100.0) | 111 (100.0) |
| | 1 | 0 (0.0) | 1 (0.9) |
| | 2 | 0 (0.0) | 0 (0.0) |
| | 3 | 2 (1.8) | 4 (3.6) |
| | 4 | 2 (1.8) | 1 (0.9) |
| | 5 | 8 (7.3) | 9 (8.1) |
| | 6 | 8 (7.3) | 8 (7.2) |
| | 7 | 1 (0.9) | 1 (0.9) |
| | 8 | 0 (0.0) | 4 (3.6) |
| | Not known | 56 (51.4) | 55 (49.5) |
| | N/A (under 65) | 15 (13.8) | 17 (15.3) |
| | Missing | 88 (80.7) | 83 (74.8) |

*Created using: X:\HAP-FAST\Statistical Analysis - Blinded\Final Analysis\Version 1.0\Programs\Section08_Baseline.sas*
*'Missing' category includes 'N/A', 'Not collected' and 'Not known' categories*

#### 8.1 Baseline characteristics by stage 2 randomisation

The tables in this subsection are supplementary, as baseline tables by stage 2 randomisation were not originally planned in the statistical analysis plan.

Supplementary Table 8-1 Demographic details by stage 2 allocation

| | | FAPP | No FAPP |
|---|---|---|---|
| Randomised: n (%) | Total | 31 (100.0) | 31 (100.0) |
| | Consent provided | 28 (90.3) | 26 (83.9) |
| | Consent not obtained | 0 (0.0) | 0 (0.0) |
| | Did not provide consent | 3 (9.7) | 5 (16.1) |
| Age (years) | N | 31 | 31 |
| | Mean (SD) | 71.1 (12.8) | 70.5 (14.8) |
| | Median (IQR) | 74.0 (64.0, 80.0) | 73.0 (65.5, 81.5) |
| | Range | (24, 87) | (20, 88) |
| | Missing | 0 | 3 |
| Sex: n (%) | Total | 31 (100.0) | 31 (100.0) |
| | Male | 13 (41.9) | 16 (51.6) |
| | Female | 18 (58.1) | 15 (48.4) |
| Height (cm) | N | 31 | 31 |
| | Mean (SD) | 164 (11.8) | 165 (10.1) |
| | Median (IQR) | 163 (160, 170) | 165 (157, 173) |
| | Range | (125, 198) | (150, 180) |
| | Missing | 0 | 3 |
| Weight (kg) | N | 31 | 31 |
| | Mean (SD) | 68.1 (17.0) | 74.4 (25.3) |
| | Median (IQR) | 67.8 (57.2, 76.0) | 69.5 (58.5, 89.8) |
| | Range | (31, 114) | (35, 149) |
| | Missing | 0 | 3 |
| Calculated BMI | N | 31 | 31 |
| | Mean (SD) | 25.6 (8.1) | 27.3 (8.8) |
| | Median (IQR) | 23.6 (20.7, 27.1) | 27.3 (20.5, 32.1) |
| | Range | (12, 48) | (14, 49) |
| | Missing | 0 | 3 |
| Smoking status: n (%) | Total | 31 (100.0) | 31 (100.0) |
| | Current smoker | 4 (12.9) | 5 (16.1) |
| | Ex-smoker | 14 (45.2) | 11 (35.5) |
| | Never smoked | 4 (12.9) | 7 (22.6) |
| | Not known | 9 (29.0) | 5 (16.1) |
| | Missing | 9 (29.0) | 8 (25.8) |

*Created using: X:\HAP-FAST\Statistical Analysis - Blinded\Final Analysis\Version 1.0\Programs\Appendix01_BaselineStage2.sas*
*'Missing' category includes 'Not collected' and 'Not known' categories*

[7] HAP suspected date is the date of stage 1 randomisation.
[8] Multiple conditions can be present at time of admission.

Supplementary Table 8-2 Vital signs (at time of suspected HAP) by stage 2 allocation

| | | **FAPP** | **No FAPP** |
|---|---|---|---|
| Temperature (°C) | N | 31 | 31 |
| | Mean (SD) | 36.5 (0.6) | 36.9 (0.9) |
| | Median (IQR) | 36.5 (36.1, 36.8) | 36.7 (36.4, 37.3) |
| | Range | (35, 38) | (36, 39) |
| | Not collected | 1 | 0 |
| | Not known | 0 | 0 |
| | Missing | 1 | 3 |
| Systolic blood pressure (mmHg) | N | 31 | 31 |
| | Mean (SD) | 123 (21.9) | 124 (28.0) |
| | Median (IQR) | 119 (109, 142) | 116 (110, 132) |
| | Range | (84, 179) | (90, 226) |
| | Not collected | 1 | 0 |
| | Not known | 0 | 0 |
| | Missing | 1 | 3 |
| Diastolic blood pressure (mmHg) | N | 31 | 31 |
| | Mean (SD) | 73.2 (12.6) | 69.3 (12.5) |
| | Median (IQR) | 75.0 (61.0, 80.0) | 71.0 (59.5, 77.5) |
| | Range | (52, 98) | (44, 104) |
| | Not collected | 1 | 0 |
| | Not known | 0 | 0 |
| | Missing | 2 | 3 |
| Pulse rate (beats per minute) | N | 31 | 31 |
| | Mean (SD) | 93.7 (23.0) | 94.8 (21.5) |
| | Median (IQR) | 89.5 (77.0, 107) | 94.5 (78.0, 109) |
| | Range | (60, 148) | (62, 135) |
| | Not collected | 1 | 0 |
| | Not known | 0 | 0 |
| | Missing | 1 | 3 |
| Respiratory rate (breaths per minute) | N | 31 | 31 |
| | Mean (SD) | 20.2 (4.3) | 21.5 (5.8) |
| | Median (IQR) | 18.5 (17.0, 20.0) | 19.5 (18.0, 22.0) |
| | Range | (16, 32) | (16, 40) |
| | Not collected | 1 | 0 |
| | Not known | 0 | 0 |
| | Missing | 1 | 3 |

| | | | |
|---|---|---|---|
| Oxygen saturation on room air (%) | N | 31 | 31 |
| | Mean (SD) | 94.1 (3.1) | 92.6 (5.8) |
| | Median (IQR) | 94.0 (92.0, 96.0) | 94.0 (91.0, 96.0) |
| | Range | (90, 100) | (71, 98) |
| | Not collected | 1 | 0 |
| | Not known | 1 | 5 |
| | Missing | 2 | 8 |
| NEWS2 Score | N | 31 | 31 |
| | Mean (SD) | 4.2 (3.2) | 4.2 (3.4) |
| | Median (IQR) | 3.5 (2.0, 7.0) | 3.0 (1.5, 6.0) |
| | Range | (0, 10) | (0, 13) |
| | Not collected | 1 | 0 |
| | Not known | 0 | 0 |
| | Missing | 1 | 3 |

*Created using: X:\HAP-FAST\Statistical Analysis - Blinded\Final Analysis\Version 1.0\Programs\Appendix01_BaselineStage2.sas*
*'Missing' category includes 'Not collected' and 'Not known' categories*

Supplementary Table 8-3 Inpatient details and HAP signs & symptoms by stage 2 allocation

| | | FAPP | No FAPP |
|---|---|---|---|
| Ward type: n (%) | Total | 31 (100.0) | 31 (100.0) |
| | Medical | 6 (19.4) | 11 (35.5) |
| | Surgical | 7 (22.6) | 3 (9.7) |
| | A&E | 18 (58.1) | 9 (29.0) |
| | Acute medical unit | 0 (0.0) | 3 (9.7) |
| | Other | 0 (0.0) | 2 (6.5) |
| | Not known | 0 (0.0) | 0 (0.0) |
| | Missing | 0 (0.0) | 3 (9.7) |
| Clinical symptoms of HAP[9]: n (%) | N | 31 (100.0) | 31 (100.0) |
| | New/worsening shortness of breath | 18 (58.1) | 21 (67.7) |
| | Cough | 23 (74.2) | 23 (74.2) |
| | Fever | 4 (12.9) | 3 (9.7) |
| | Chest pain | 6 (19.4) | 1 (3.2) |
| | New/worsening sputum | 12 (38.7) | 13 (41.9) |
| | Confusion | 2 (6.5) | 2 (6.5) |
| | Other | 3 (9.7) | 3 (9.7) |
| | Any missing | 0 (0.0) | 0 (0.0) |
| Clinical description of respiratory finding of HAP[10]: n (%) | N | 31 (100.0) | 31 (100.0) |
| | New chest crackles | 12 (38.7) | 11 (35.5) |
| | Reduced air entry | 13 (41.9) | 5 (16.1) |
| | Respiratory distress | 9 (29.0) | 12 (38.7) |
| | Other | 2 (6.5) | 4 (12.9) |
| | Any missing | 0 (0.0) | 0 (0.0) |
| 'Medically fit for discharge' prior to HAP suspicion: n (%) | Total | 31 (100.0) | 31 (100.0) |
| | No | 18 (58.1) | 16 (51.6) |
| | Yes | 1 (3.2) | 0 (0.0) |
| | Not known | 12 (38.7) | 12 (38.7) |
| | Missing | 12 (38.7) | 15 (48.4) |

*Created using: X:\HAP-FAST\Statistical Analysis - Blinded\Final Analysis\Version 1.0\Programs\Appendix01_BaselineStage2.sas*
*'Missing' category includes 'Not collected' and 'Not known' categories*

[9] Participants may have more than one symptom and/or respiratory finding.
[10] Participants may have more than one symptom and/or respiratory finding.

Supplementary Table 8-4 Concomitant medications and immunosuppressant medication use (at time of suspected HAP) by stage 2 allocation

| | | FAPP | No FAPP |
|---|---|---|---|
| Antibiotics taken/received in last 7 days: n (%) | Total | 31 (100.0) | 31 (100.0) |
| | No | 15 (48.4) | 12 (38.7) |
| | Yes | 14 (45.2) | 15 (48.4) |
| | Missing | 2 (6.5) | 4 (12.9) |
| Received immune suppressant mediation (excluding cancer treatment) in the 3 months prior to suspected HAP: n (%) | Total | 31 (100.0) | 31 (100.0) |
| | No | 21 (67.7) | 14 (45.2) |
| | Yes | 0 (0.0) | 0 (0.0) |
| | Not known | 10 (32.3) | 14 (45.2) |
| | Missing | 10 (32.3) | 17 (54.8) |
| Received chemotherapy for a solid organ or haematological cancer in the 3 months prior to suspected HAP: n (%) | Total | 31 (100.0) | 31 (100.0) |
| | No | 20 (64.5) | 16 (51.6) |
| | Yes | 1 (3.2) | 0 (0.0) |
| | Not known | 10 (32.3) | 12 (38.7) |
| | Missing | 10 (32.3) | 15 (48.4) |
| **Concomitant medications taken or received in the last 7 days** | | | |
| Steroids (oral, IV, inhaled): n (%) | Total | 31 (100.0) | 31 (100.0) |
| | Unchecked | 28 (90.3) | 26 (83.9) |
| | Checked | 3 (9.7) | 5 (16.1) |
| Antipsychotics: n (%) | Total | 31 (100.0) | 31 (100.0) |
| | Unchecked | 31 (100.0) | 31 (100.0) |
| | Checked | 0 (0.0) | 0 (0.0) |
| Proton pump inhibitors: n (%) | Total | 31 (100.0) | 31 (100.0) |
| | Unchecked | 24 (77.4) | 25 (80.6) |
| | Checked | 7 (22.6) | 6 (19.4) |
| Statins: n (%) | Total | 31 (100.0) | 31 (100.0) |
| | Unchecked | 25 (80.6) | 27 (87.1) |
| | Checked | 6 (19.4) | 4 (12.9) |
| None of the above concomitant medications: n (%) | Total | 31 (100.0) | 31 (100.0) |
| | Unchecked | 12 (38.7) | 12 (38.7) |
| | Checked | 19 (61.3) | 19 (61.3) |

*Created using: X:\HAP-FAST\Statistical Analysis - Blinded\Final Analysis\Version 1.0\Programs\Appendix01_BaselineStage2.sas*
*'Missing' category includes 'Not collected' and 'Not known' categories*

Supplementary Table 8-5 Admission details by stage 2 allocation

| | | FAPP | No FAPP |
|---|---|---|---|
| Time from admission to HAP suspected[11] (days) | N | 31 | 31 |
| | Mean (SD) | 9.6 (23.1) | 9.7 (15.2) |
| | Median (IQR) | 0.0 (0.0, 9.2) | 3.0 (0.0, 14.1) |
| | Range | (0, 106) | (0, 66) |
| | Missing | 0 | 3 |
| Reason for admission: n (%) | Total | 31 (100.0) | 31 (100.0) |
| | Medical | 26 (83.9) | 24 (77.4) |
| | Surgical | 5 (16.1) | 4 (12.9) |
| | Missing | 0 (0.0) | 3 (9.7) |
| Had the following conditions at the point of admission: n (%) | N | 31 (100.0) | 31 (100.0) |
| | None of below | 16 (51.6) | 8 (25.8) |
| | Congestive cardiac failure | 1 (3.2) | 3 (9.7) |
| | Dementia | 1 (3.2) | 0 (0.0) |
| | Other form of cognitive impairment | 0 (0.0) | 2 (6.5) |
| | Myocardial Infarction | 0 (0.0) | 3 (9.7) |
| | Chronic pulmonary disease | 8 (25.8) | 12 (38.7) |
| | Rheumatic disease | 0 (0.0) | 1 (3.2) |
| | AIDS (the syndrome not just HIV positive) | 0 (0.0) | 0 (0.0) |
| | Hemiplegia or paraplegia | 0 (0.0) | 1 (3.2) |
| | Renal disease | 2 (6.5) | 1 (3.2) |
| | Liver disease | 0 (0.0) | 0 (0.0) |
| | Diabetes | 5 (16.1) | 4 (12.9) |
| | Any malignancy | 1 (3.2) | 1 (3.2) |
| | Any missing | 0 (0.0) | 0 (0.0) |
| Oxygen saturation on room air at time of admission (%) | N | 31 | 31 |
| | Mean (SD) | 94.2 (4.5) | 93.0 (6.2) |
| | Median (IQR) | 95.0 (92.0, 97.0) | 94.5 (92.0, 97.0) |
| | Range | (77, 100) | (71, 99) |
| | Missing | 0 | 5 |
| Could eat and drink independently prior to admission: n (%) | Total | 31 (100.0) | 31 (100.0) |
| | No | 3 (9.7) | 6 (19.4) |
| | Yes | 21 (67.7) | 17 (54.8) |
| | Not known | 7 (22.6) | 5 (16.1) |
| | Missing | 7 (22.6) | 8 (25.8) |
| Clinical frailty score: n (%) | Total | 31 (100.0) | 31 (100.0) |
| | 1 | 0 (0.0) | 0 (0.0) |
| | 2 | 0 (0.0) | 0 (0.0) |
| | 3 | 0 (0.0) | 1 (3.2) |
| | 4 | 1 (3.2) | 0 (0.0) |
| | 5 | 2 (6.5) | 2 (6.5) |
| | 6 | 3 (9.7) | 1 (3.2) |
| | 7 | 0 (0.0) | 0 (0.0) |
| | 8 | 0 (0.0) | 0 (0.0) |
| | Not known | 17 (54.8) | 19 (61.3) |
| | N/A (under 65) | 8 (25.8) | 5 (16.1) |
| | Missing | 25 (80.6) | 27 (87.1) |

*Created using: X:\HAP-FAST\Statistical Analysis - Blinded\Final Analysis\Version 1.0\Programs\Appendix01_BaselineStage2.sas*
*'Missing' category includes 'N/A', 'Not collected' and 'Not known' categories*

[11] HAP suspected date is the date of stage 1 randomisation.

### 9 Study population

Table 9-1 Data sets analysed

| Population | CT | CXR |
|---|---|---|
| Screened | 109 | 111 |
| Randomised | 109 | 111 |
| Intention-to-treat | 109 (100.0%) | 111 (100.0%) |

*Created using: X:\HAP-FAST\Statistical Analysis - Blinded\Final Analysis\Version 1.0\Programs\Section09_StudyPopAnalysisSetDefs.sas*

### 10 Protocol deviations

Table 10-1 Protocol deviations

| **Protocol deviations: n (% of patients)** | **CT** | **CXR** |
|---|---|---|
| Count of all protocol deviations | 120 | 118 |
| At least one protocol deviation | 56 (50.5) | 54 (49.5) |
| Missed follow-up visit time point | 51 (45.9) | 44 (40.4) |
| Day 1 | 17 (15.3) | 11 (10.1) |
| Day 2 | 12 (10.8) | 10 (9.2) |
| Day 3 | 6 (5.4) | 8 (7.3) |
| Day 4 | 3 (2.7) | 5 (4.6) |
| Day 5 | 3 (2.7) | 3 (2.8) |
| Day 6 | 3 (2.7) | 4 (3.7) |
| Day 7 | 3 (2.7) | 2 (1.8) |
| Day 8 | 2 (1.8) | 3 (2.8) |
| Day 9 | 2 (1.8) | 2 (1.8) |
| Day 10 | 4 (3.6) | 2 (1.8) |
| Day 28 | 27 (24.3) | 27 (24.8) |
| Day 90 | 33 (29.7) | 25 (22.9) |
| Failed stage 1 eligibility criteria: | 1 (0.9) | 8 (7.3) |
| Already received chest X-ray or CT scan | 0 (0.0) | 4 (3.7) |
| Patient ineligible - unknown reason | 1 (0.9) | 1 (0.9) |
| Intend to palliate patient | 0 (0.0) | 1 (0.9) |
| Suspected pulmonary embolism | 0 (0.0) | 1 (0.9) |
| Not suspected HAP | 0 (0.0) | 1 (0.9) |
| Did not meet stage 2 eligibility criteria | 3 (2.7) | 5 (4.6) |
| Sputum not obtained before second dose of antibiotic | 2 (1.8) | 4 (3.7) |
| Patient not treated with antibiotics for HAP after stage 1 scan results | 1 (0.9) | 1 (0.9) |
| Sample received before informed consent obtained | 1 (0.9) | 3 (2.8) |
| Day 0 sample | 1 (0.9) | 3 (2.8) |

*Created using: X:\HAP-FAST\Statistical Analysis - Blinded\Final Analysis\Version 1.0\Programs\Section10_Deviations.sas*

### 11 Compliance with intervention

Table 11-1 Summary statistics – time from stage 1 randomisation to scan and to scan report

| | | CT | CXR |
|---|---|---|---|
| Time from stage 1 randomisation to scan (hours) | Scans conducted | 89 | 98 |
| | Mean (SE) | 5.5 (1.3) | 3.1 (0.7) |
| | Median (IQR) | 2 (1, 4) | 1 (1, 2) |
| | Range | (0, 92) | (0, 53) |
| | Excluded: scan before randomisation | 0 | 1 |
| | Excluded: scan not done | 20 | 12 |
| Time from stage 1 randomisation to scan report (hours) | Scans conducted | 89 | 98 |
| | Mean (SE) | 23.5 (7.8) | 142 (22.1)[12] |
| | Median (IQR) | 4 (3, 13) | 50 (18, 135) |
| | Range | (0, 605) | (1, †) |
| | Excluded: scan before randomisation | 0 | 1 |
| | Excluded: scan not done | 20 | 12 |

*Created using: X:\HAP-FAST\Statistical Analysis - Blinded\Final Analysis\Version 1.0\Programs\Section11_Compliance.sas*

Table 11-2 Reasons for not receiving allocated intervention (CT scan or chest X-ray)

| | CT | CXR |
|---|---|---|
| N | 109 (100.0) | 111 (100.0) |
| Did not receive allocated intervention: n (%) | 34 (31.2) | 12 (10.8) |
| Form incomplete[13] | 17 (15.6) | 11 (9.9) |
| Operational constraints | 6 (5.5) | 0 (0.0) |
| Unable to tolerate scan | 6 (5.5) | 1 (0.9) |
| Withdrawn from study before scan conducted | 2 (1.8) | 0 (0.0) |
| Unknown reason | 3 (2.8) | 0 (0.0) |

*Created using: X:\HAP-FAST\Statistical Analysis - Blinded\Final Analysis\Version 1.0\Programs\Section11_Compliance.sas*

Table 11-3 Reasons for not receiving FAPP (if allocated to FAPP)

| | FAPP |
|---|---|
| N | 31 (100.0) |
| Did not receive FAPP: n (%) | 6 (19.4) |
| Misplaced sample | 3 (9.7) |
| FAPP result invalid | 2 (6.5) |
| Sample not obtained | 1 (3.2) |

*Created using: X:\HAP-FAST\Statistical Analysis - Blinded\Final Analysis\Version 1.0\Programs\Section11_Compliance.sas*

[12] This estimated mean is biased towards zero because the latest observation is censored.
† Quantity cannot be estimated due to censoring of later observations.
[13] In these participants, the *CT/Chest X-ray details* CRF was not filled out, and this is assumed to mean patients did not receive their allocated stage 1 intervention.

### 12 Outcome Data

#### 12.1 Primary outcome

The key objective and primary outcome of this trial is determining the feasibility of a future definitive randomised controlled trial. This will be answered by assessing the secondary outcomes.

#### 12.2 Secondary outcome 1 – Time to clinical cure

Figure 12-1 Kaplan-Meier plots of time to clinical cure for each treatment arm

*Figure 12-1 could not be produced as no participants achieved clinical cure as defined in the protocol.*
*Created using: X:\HAP-FAST\Statistical Analysis - Blinded\Final Analysis\Version 1.0\Programs\Section12-2_TimeToClinicalCure.sas*

Table 12-1 Summary statistics of time to clinical cure.

*Table 12-1 could not be produced as no participants achieved clinical cure as defined in the protocol.*
*Created using: X:\HAP-FAST\Statistical Analysis - Blinded\Final Analysis\Version 1.0\Programs\Section12-2_TimeToClinicalCure.sas*

Table 12-2 Number and percentage of participants experiencing clinical cure who went onto fail clinical cure at a later visit (days 1-10)

*Table 12-2 could not be produced as no participants achieved clinical cure as defined in the protocol.*
*Created using: X:\HAP-FAST\Statistical Analysis - Blinded\Final Analysis\Version 1.0\Programs\Section12-2_TimeToClinicalCure.sas*

Table 12-3 Summary statistics – percentage change in CAP-Sym score from baseline to time of clinical cure

*Table 12-3 could not be produced as no participants achieved clinical cure as defined in the protocol.*
*Created using: X:\HAP-FAST\Statistical Analysis - Blinded\Final Analysis\Version 1.0\Programs\Section12-2_TimeToClinicalCure.sas*

Table 12-4 Number and percentage of participants who could not be assessed for clinical cure and failed clinical cure

| **Did not experience clinical cure** | **CT** | | **CXR** | | **FAPP** | | **No FAPP** | |
|---|---|---|---|---|---|---|---|---|
| | **n** | **%** | **n** | **%** | **n** | **%** | **n** | **%** |
| Did not experience clinical cure: | 109 | 100.0 | 111 | 100.0 | 31 | 100.0 | 31 | 100.0 |
| Failed clinical cure | 0 | 0.0 | 1 | 0.9 | 0 | 0.0% | 1 | 3.2 |
| Could not be assessed | 109 | 100.0 | 110 | 99.1 | 31 | 100.0 | 30 | 96.8 |
| Component(s) clinical cure failed on[14]: | 0 | 0.0 | 1 | 100.0 | 0 | 0.0 | 1 | 100.0 |
| Clinical stability component | 0 | 0.0 | 1 | 100.0 | 0 | 0.0 | 1 | 100.0 |
| Radiological component | 0 | 0.0 | 1 | 100.0 | 0 | 0.0 | 1 | 100.0 |
| CAP-Sym component | 0 | 0.0 | 0 | 0.0 | 0 | 0.0 | 0 | 0.0 |
| Could not be assessed: | 109 | 100.0 | 110 | 100.0 | 31 | 100.0 | 30 | 100.0 |
| Discharged before repeat chest imagine | 28 | 25.7 | 40 | 36.4 | 15 | 48.4 | 8 | 26.7 |
| Died without repeat chest imaging | 4 | 3.7 | 5 | 4.5 | 0 | 0.0 | 3 | 10.0 |
| Did not consent by day 3 | 49 | 45.0 | 41 | 37.3 | 4 | 12.9 | 6 | 20.0 |
| Declined consent between days 4 and 6 | 5 | 4.6 | 3 | 2.7 | 1 | 3.2 | 0 | 0.0 |
| Withdrew consent before day 7 | 31 | 28.4 | 28 | 25.5 | 2 | 6.5 | 5 | 16.7 |
| No repeat chest imaging up to day 10 | 108 | 99.1 | 107 | 97.3 | 30 | 96.8 | 29 | 96.7 |
| Incomplete data[15] (see Table 12-19) | 109 | 100.0 | 110 | 100.0 | 31 | 100.0 | 30 | 100.0 |

*Created using: X:\HAP-FAST\Statistical Analysis - Blinded\Final Analysis\Version 1.0\Programs\Section12-2_TimeToClinicalCure.sas*

Table 12-5 Kaplan-Meier estimates and Cox Proportional Hazards models estimates – time to clinical cure

*Table 12-3 could not be produced as no participants achieved clinical cure as defined in the protocol.*
*Created using: X:\HAP-FAST\Statistical Analysis - Blinded\Final Analysis\Version 1.0\Programs\Section12-2_TimeToClinicalCure.sas*

##### 12.2.1 Relaxed clinical cure criteria

Given almost all patients could not be assessed for clinical cure, and the single patient who could was not cured, we consider patients under a relaxed set of criteria for clinical cure. In the relaxed criteria, a radiological scan is not required, but resolution of clinical signs and the same improvement in CAP-Sym score are still required. This analysis was not included in the protocol or statistical analysis plan, so results are listed as supplementary.

[14] Only the participant's best result up to day 10, i.e. the result from the day they failed on the least components, is included in this table. Participants may have failed on more than one component.
[15] Participants may be assessed for clinical cure multiple times (up to day 10). If they never had complete data to assess clinical cure on any of these days, they are classed as having incomplete data.

Supplementary Figure 1 Kaplan-Meier time to cure curves for relaxed clinical cure criteria

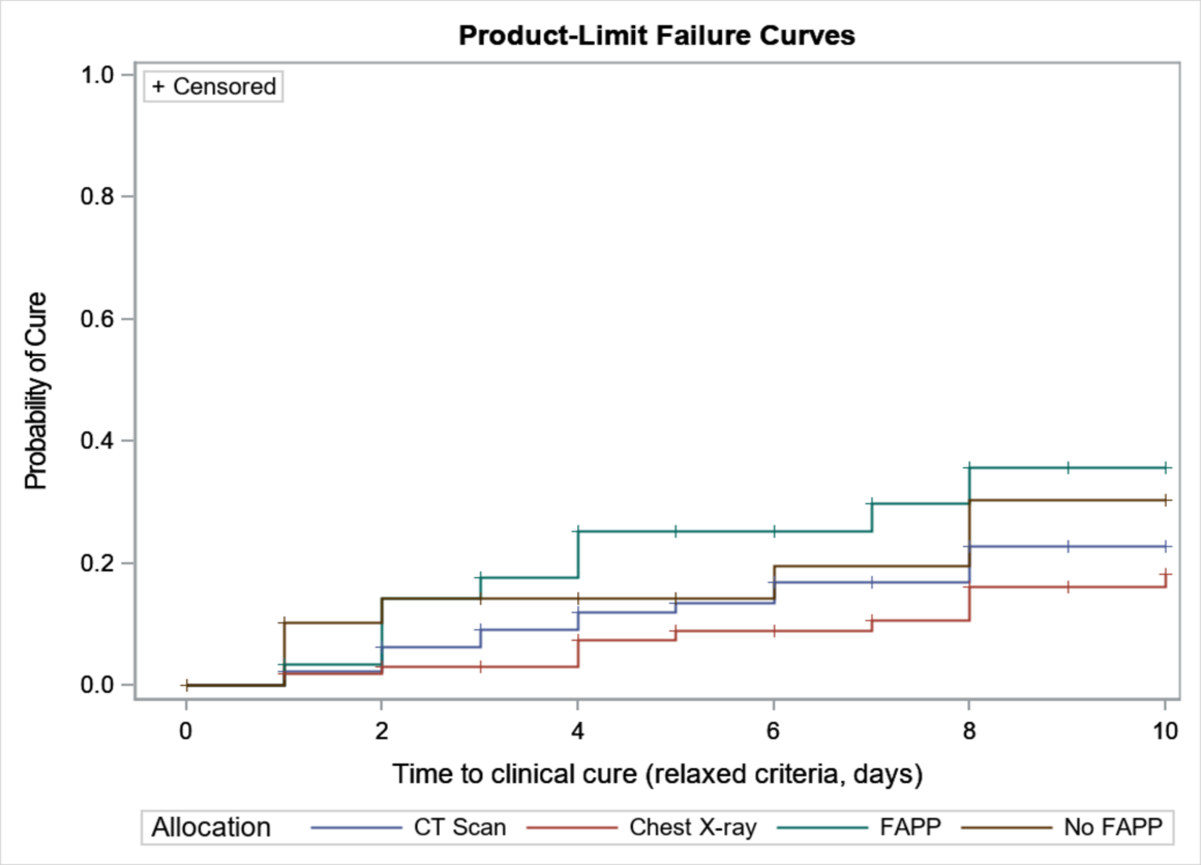

*Created using: X:\HAP-FAST\Statistical Analysis - Blinded\Final Analysis\Version 1.0\Programs\Section12-2_TimeToClinicalCure.sas*

Supplementary Table 12-1 Reasons for participants not experiencing relaxed clinical cure

| Did not experience clinical cure | CT | | CXR | | FAPP | | No FAPP | |
|---|---|---|---|---|---|---|---|---|
| | n | % | n | % | n | % | n | % |
| Did not experience clinical cure[16]: | 94 | 86.2 | 99 | 89.2 | 22 | 71.0 | 24 | 77.4 |
| Failed clinical cure | 21 | 19.3 | 32 | 28.8 | 14 | 45.2 | 9 | 29.0 |
| Could not be assessed | 73 | 67.0 | 67 | 60.4 | 8 | 25.8 | 15 | 48.4 |
| Component(s) clinical cure failed on[17]: | 21 | 100.0 | 32 | 100.0 | 14 | 100.0 | 9 | 100.0 |
| Clinical stability component | 7 | 33.3 | 10 | 31.3 | 5 | 35.7 | 3 | 33.3 |
| CAP-Sym component | 20 | 95.2 | 30 | 93.8 | 13 | 92.9 | 7 | 77.8 |
| Could not be assessed: | 73 | 100.0 | 67 | 100.0 | 8 | 100.0 | 15 | 100.0 |
| Did not consent by day 3 | 49 | 67.1 | 41 | 61.2 | 4 | 50.0 | 6 | 40.0 |
| Declined consent between days 4 and 6 | 3 | 4.1 | 3 | 4.5 | 0 | 0.0 | 0 | 0.0 |
| Withdrew consent before day 7 | 29 | 39.7 | 27 | 40.3 | 1 | 12.5 | 5 | 33.3 |
| Incomplete data[18] (see Table 12-19) | 73 | 100.0 | 67 | 100.0 | 8 | 100.0 | 15 | 100.0 |

*Created using: X:\HAP-FAST\Statistical Analysis - Blinded\Final Analysis\Version 1.0\Programs\Section12-2_TimeToClinicalCure.sas*

#### 12.3 Secondary outcome 2 – Antibiotics usage for the HAP episode

Figure 12-2 Boxplots of antibiotics usage in defined daily doses (DDD) for the HAP episode for each treatment arm

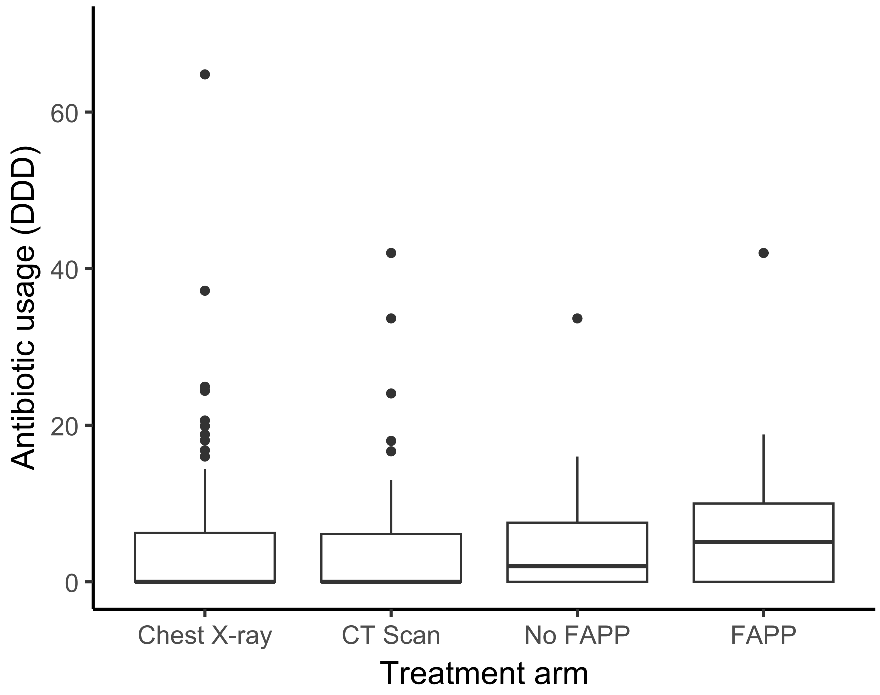

[16] For these table rows, the denominator includes all randomised patients.
[17] Only the participant's best result up to day 10, i.e. the result from the day they failed on the least components, is included in this table. Participants may have failed on more than one component. Denominator is patients who failed clinical cure.
[18] Participants may be assessed for clinical cure multiple times (up to day 10). If they never had complete data to assess clinical cure on any of these days, they are classed as having incomplete data. Denominator is patients who could not be assessed for clinical cure.

*Created using: X:/HAP-FAST/Statistical Analysis - Blinded/Final Analysis/Version 1.0/Programs/Section12-3_AntibioticUsage.Rmd*

Table 12-6 Summary statistics – antibiotic usage in DDD for the HAP episode

| Measure | CT | CXR | FAPP | No FAPP |
|---|---|---|---|---|
| N | 109 | 111 | 31 | 31 |
| Mean (SD) | 4.0 (7.2) | 4.8 (9.1) | 7.1 (9.4) | 5.4 (7.9) |
| Median (IQR) | 0.0 (0.0, 6.1) | 0.0 (0.0, 6.2) | 5.1 (0.0, 10.0) | 2.0 (0.0, 7.5) |
| Range | (0.0, 42.0) | (0.0, 64.8) | (0.0, 42.0) | (0.0, 33.6) |
| No antibiotics (% of non-missing) | 50 (56.8) | 53 (52.5) | 7 (29.2) | 10 (43.5) |
| Missing (%) | 21 (19.3) | 10 (9.0) | 7 (22.6) | 8 (25.8) |

*Created using: X:/HAP-FAST/Statistical Analysis - Blinded/Final Analysis/Version 1.0/Programs/Section12-3_AntibioticUsage.Rmd*

The Statistical Analysis Plan indicates that antibiotic usage will be modelled according to a Poisson regression model (or similar). However, antibiotic dose in defined daily doses (DDD) takes non-integer values as well as zeros, so Poisson regression cannot be used. The following language is suggested for the model description:

> *Antibiotic use, measured in defined daily doses (DDD), is modelled using a two-step gamma hurdle model (glmmTMB R package version 1.1.10). This model estimates both the probability of having no antibiotics (e.g. where a patient's scan results indicate they do not have HAP), and the amount of antibiotics used in patients that have had any antibiotics. Both parts of this model use stage 1 allocation and site as the predictors. The probability of no antibiotics is linked to the linear predictor using a logit link, and the amount of antibiotics conditional on having any antibiotics is assumed to be gamma distributed, linked to the linear predictor using an identity link. For comparing the FAPP versus No FAPP arms, the same model was used except the stage 1 allocation predictor was replaced by stage 2 allocation. Comparisons are presented as marginal effects.*

Table 12-7 Estimates of antibiotic usage for the HAP episode, from a gamma hurdle regression model. 95% confidence intervals are shown in parentheses. Adjusted measures compare a change in treatment for a patient at the same site. Marginal differences compare a change in treatment averaged over all participants. DDD: defined daily doses.

| Comparison | Adjusted odds ratio of receiving no antibiotics | Adjusted difference in mean antibiotic use, participants with any antibiotics, DDD | Marginal difference in risk of receiving no antibiotics, percentage points | Marginal difference in mean antibiotic use, all participants, DDD |
|---|---|---|---|---|
| **CT – CXR** | 1.29 (0.69, 2.41) | 1.0 (-2.2, 4.1) | 5.5 (-7.7, 18.6) | -0.1 (-1.9, 1.8) |
| **FAPP – No FAPP** | 0.35 (0.09, 1.34) | -0.5 (-6.3, 5.2) | -21.0 (-46.6, 4.5) | 1.8 (-2.7, 6.2) |

*Created using: X:/HAP-FAST/Statistical Analysis - Blinded/Final Analysis/Version 1.0/Programs/Section12-3_AntibioticUsage.Rmd*

Diagnosis for this model is inconclusive on whether the effect of randomisation differs by site (Supplementary Figure 2), which might indicate systematically different implementations of treatment between sites. The median antibiotic usage is higher for CXR than CT at Royal Liverpool and Wythenshawe hospitals, but this is reversed for Royal Preston Hospital. However, antibiotic usage varies considerably within and between sites, which suggests that these observed differences may not arise from systematic differences between sites.

Supplementary Figure 2 Boxplots of antibiotic usage by stage 1 allocation and site. DDD: defined daily doses.

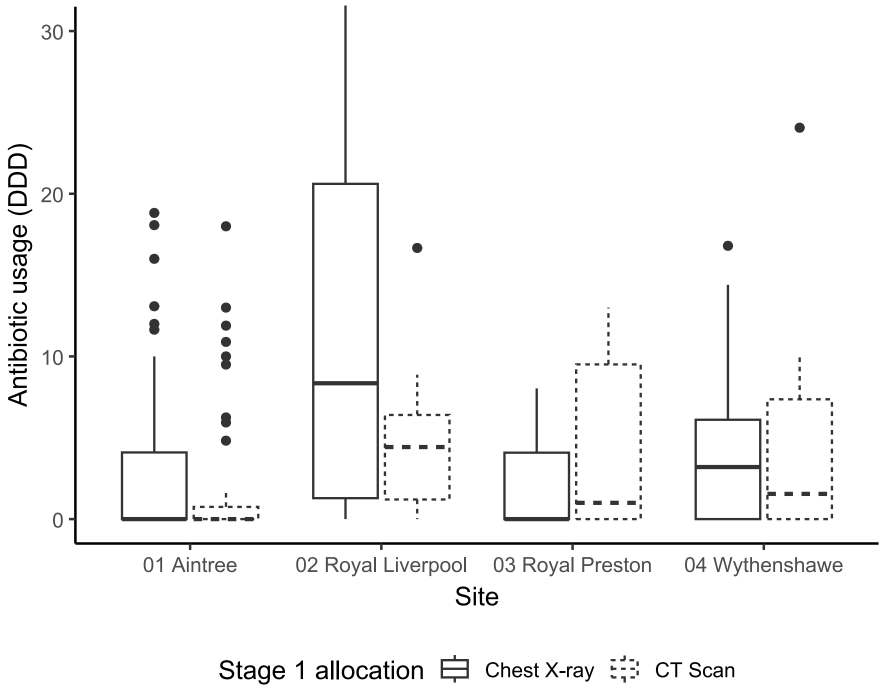

#### 12.4 Secondary outcome 3 – EQ-5D-5L

Table 12-8 Number and percentage of participants in each severity level for mobility (baseline, day 10, day 28 and day 90)

| Dimension | Timepoint | Severity level | CT | CXR | FAPP | No FAPP |
|---|---|---|---|---|---|---|
| Mobility | Baseline | Total responded | 46 (100.0) | 56 (100.0) | 24 (100.0) | 24 (100.0) |
| | | No problems | 4 (8.7) | 4 (7.1) | 2 (8.3) | 1 (4.2) |
| | | Slight problems | 2 (4.3) | 8 (14.3) | 3 (12.5) | 1 (4.2) |
| | | Moderate problems | 12 (26.1) | 13 (23.2) | 6 (25.0) | 8 (33.3) |
| | | Severe problems | 13 (28.3) | 14 (25.0) | 7 (29.2) | 5 (20.8) |
| | | Unable to | 15 (32.6) | 17 (30.4) | 6 (25.0) | 9 (37.5) |
| | | Missing | 22 | 18 | 4 | 2 |
| | Day 10 | Total responded | 21 (100.0) | 27 (100.0) | 12 (100.0) | 11 (100.0) |
| | | No problems | 4 (19.0) | 4 (14.8) | 2 (16.7) | 1 (9.1) |
| | | Slight problems | 4 (19.0) | 1 (3.7) | 0 (0.0) | 3 (27.3) |
| | | Moderate problems | 2 (9.5) | 6 (22.2) | 3 (25.0) | 2 (18.2) |
| | | Severe problems | 7 (33.3) | 7 (25.9) | 4 (33.3) | 2 (18.2) |
| | | Unable to | 4 (19.0) | 9 (33.3) | 3 (25.0) | 3 (27.3) |
| | | Missing | 40 | 44 | 15 | 12 |
| | Day 28 | Total responded | 22 (100.0) | 30 (100.0) | 12 (100.0) | 10 (100.0) |
| | | No problems | 4 (18.2) | 4 (13.3) | 1 (8.3) | 1 (10.0) |
| | | Slight problems | 2 (9.1) | 3 (10.0) | 0 (0.0) | 3 (30.0) |
| | | Moderate problems | 4 (18.2) | 9 (30.0) | 5 (41.7) | 0 (0.0) |
| | | Severe problems | 6 (27.3) | 6 (20.0) | 2 (16.7) | 3 (30.0) |
| | | Unable to | 6 (27.3) | 8 (26.7) | 4 (33.3) | 3 (30.0) |
| | | Missing | 33 | 34 | 14 | 9 |
| | Day 90 | Total responded | 23 (100.0) | 26 (100.0) | 13 (100.0) | 8 (100.0) |
| | | No problems | 3 (13.0) | 8 (30.8) | 4 (30.8) | 2 (25.0) |
| | | Slight problems | 4 (17.4) | 5 (19.2) | 1 (7.7) | 0 (0.0) |
| | | Moderate problems | 6 (26.1) | 6 (23.1) | 3 (23.1) | 3 (37.5) |
| | | Severe problems | 7 (30.4) | 2 (7.7) | 3 (23.1) | 1 (12.5) |
| | | Unable to | 3 (13.0) | 5 (19.2) | 2 (15.4) | 2 (25.0) |
| | | Missing | 28 | 33 | 13 | 9 |

*Created using: X:\HAP-FAST\Statistical Analysis - Blinded\Final Analysis\Version 1.0\Programs\Section12-4_EQ5D5L.sas*
*Patients are included if a questionnaire is expected (i.e. they had consented and had not withdrawn at that timepoint).*

Table 12-9 Number and percentage of participants in each severity level for self-care (baseline, day 10, day 28 and day 90)

| Dimension | Timepoint | Severity level | CT | CXR | FAPP | No FAPP |
|---|---|---|---|---|---|---|
| Self-care | Baseline | Total responded | 46 (100.0) | 56 (100.0) | 24 (100.0) | 24 (100.0) |
| | | No problems | 12 (26.1) | 8 (14.3) | 3 (12.5) | 3 (12.5) |
| | | Slight problems | 5 (10.9) | 6 (10.7) | 2 (8.3) | 2 (8.3) |
| | | Moderate problems | 3 (6.5) | 9 (16.1) | 4 (16.7) | 4 (16.7) |
| | | Severe problems | 12 (26.1) | 14 (25.0) | 6 (25.0) | 7 (29.2) |
| | | Unable to | 14 (30.4) | 19 (33.9) | 9 (37.5) | 8 (33.3) |
| | | Missing | 22 | 18 | 4 | 2 |
| | Day 10 | Total responded | 21 (100.0) | 27 (100.0) | 12 (100.0) | 11 (100.0) |
| | | No problems | 7 (33.3) | 4 (14.8) | 3 (25.0) | 4 (36.4) |
| | | Slight problems | 3 (14.3) | 3 (11.1) | 1 (8.3) | 1 (9.1) |
| | | Moderate problems | 4 (19.0) | 7 (25.9) | 4 (33.3) | 2 (18.2) |
| | | Severe problems | 4 (19.0) | 5 (18.5) | 1 (8.3) | 2 (18.2) |
| | | Unable to | 3 (14.3) | 8 (29.6) | 3 (25.0) | 2 (18.2) |
| | | Missing | 40 | 44 | 15 | 12 |
| | Day 28 | Total responded | 22 (100.0) | 30 (100.0) | 12 (100.0) | 10 (100.0) |
| | | No problems | 7 (31.8) | 9 (30.0) | 3 (25.0) | 5 (50.0) |
| | | Slight problems | 2 (9.1) | 1 (3.3) | 0 (0.0) | 1 (10.0) |
| | | Moderate problems | 8 (36.4) | 6 (20.0) | 3 (25.0) | 2 (20.0) |
| | | Severe problems | 0 (0.0) | 3 (10.0) | 0 (0.0) | 0 (0.0) |
| | | Unable to | 5 (22.7) | 11 (36.7) | 6 (50.0) | 2 (20.0) |
| | | Missing | 33 | 34 | 14 | 9 |
| | Day 90 | Total responded | 23 (100.0) | 26 (100.0) | 13 (100.0) | 8 (100.0) |
| | | No problems | 9 (39.1) | 12 (46.2) | 6 (46.2) | 3 (37.5) |
| | | Slight problems | 6 (26.1) | 3 (11.5) | 4 (30.8) | 1 (12.5) |
| | | Moderate problems | 3 (13.0) | 5 (19.2) | 0 (0.0) | 0 (0.0) |
| | | Severe problems | 1 (4.3) | 2 (7.7) | 0 (0.0) | 2 (25.0) |
| | | Unable to | 4 (17.4) | 4 (15.4) | 3 (23.1) | 2 (25.0) |
| | | Missing | 28 | 33 | 13 | 9 |

*Created using: X:\HAP-FAST\Statistical Analysis - Blinded\Final Analysis\Version 1.0\Programs\Section12-4_EQ5D5L.sas*
*Patients are included if a questionnaire is expected (i.e. they had consented and had not withdrawn at that timepoint).*

Table 12-10 Number and percentage of participants in each severity level for usual activities (baseline, day 10, day 28 and day 90)

| Dimension | Timepoint | Severity level | CT | CXR | FAPP | No FAPP |
|---|---|---|---|---|---|---|
| Usual activities | Baseline | Total responded | 46 (100.0) | 56 (100.0) | 24 (100.0) | 24 (100.0) |
| | | No problems | 7 (15.2) | 4 (7.1) | 4 (16.7) | 0 (0.0) |
| | | Slight problems | 0 (0.0) | 7 (12.5) | 0 (0.0) | 1 (4.2) |
| | | Moderate problems | 7 (15.2) | 6 (10.7) | 2 (8.3) | 4 (16.7) |
| | | Severe problems | 9 (19.6) | 8 (14.3) | 2 (8.3) | 6 (25.0) |
| | | Unable to | 23 (50.0) | 31 (55.4) | 16 (66.7) | 13 (54.2) |
| | | Missing | 22 | 18 | 4 | 2 |
| | Day 10 | Total responded | 21 (100.0) | 27 (100.0) | 12 (100.0) | 11 (100.0) |
| | | No problems | 3 (14.3) | 3 (11.1) | 2 (16.7) | 1 (9.1) |
| | | Slight problems | 1 (4.8) | 5 (18.5) | 1 (8.3) | 2 (18.2) |
| | | Moderate problems | 5 (23.8) | 5 (18.5) | 2 (16.7) | 3 (27.3) |
| | | Severe problems | 4 (19.0) | 5 (18.5) | 3 (25.0) | 2 (18.2) |
| | | Unable to | 8 (38.1) | 9 (33.3) | 4 (33.3) | 3 (27.3) |
| | | Missing | 40 | 44 | 15 | 12 |
| | Day 28 | Total responded | 22 (100.0) | 30 (100.0) | 12 (100.0) | 10 (100.0) |
| | | No problems | 6 (27.3) | 4 (13.3) | 1 (8.3) | 2 (20.0) |
| | | Slight problems | 1 (4.5) | 3 (10.0) | 0 (0.0) | 2 (20.0) |
| | | Moderate problems | 6 (27.3) | 5 (16.7) | 2 (16.7) | 3 (30.0) |
| | | Severe problems | 1 (4.5) | 5 (16.7) | 0 (0.0) | 1 (10.0) |
| | | Unable to | 8 (36.4) | 13 (43.3) | 9 (75.0) | 2 (20.0) |
| | | Missing | 33 | 34 | 14 | 9 |
| | Day 90 | Total responded | 23 (100.0) | 26 (100.0) | 13 (100.0) | 8 (100.0) |
| | | No problems | 3 (13.0) | 5 (19.2) | 2 (15.4) | 2 (25.0) |
| | | Slight problems | 6 (26.1) | 5 (19.2) | 4 (30.8) | 2 (25.0) |
| | | Moderate problems | 4 (17.4) | 3 (11.5) | 1 (7.7) | 0 (0.0) |
| | | Severe problems | 5 (21.7) | 5 (19.2) | 1 (7.7) | 2 (25.0) |
| | | Unable to | 5 (21.7) | 8 (30.8) | 5 (38.5) | 2 (25.0) |
| | | Missing | 28 | 33 | 13 | 9 |

*Created using: X:\HAP-FAST\Statistical Analysis - Blinded\Final Analysis\Version 1.0\Programs\Section12-4_EQ5D5L.sas*
*Patients are included if a questionnaire is expected (i.e. they had consented and had not withdrawn at that timepoint).*

Table 12-11 Number and percentage of participants in each severity level for pain or discomfort (baseline, day 10, day 28 and day 90)

| Dimension | Timepoint | Severity level | CT | CXR | FAPP | No FAPP |
|---|---|---|---|---|---|---|
| Pain or discomfort | Baseline | Total responded | 46 (100.0) | 56 (100.0) | 24 (100.0) | 24 (100.0) |
| | | No pain | 17 (37.0) | 18 (32.1) | 8 (33.3) | 6 (25.0) |
| | | Slight pain | 6 (13.0) | 10 (17.9) | 1 (4.2) | 3 (12.5) |
| | | Moderate pain | 10 (21.7) | 14 (25.0) | 5 (20.8) | 8 (33.3) |
| | | Severe pain | 9 (19.6) | 8 (14.3) | 6 (25.0) | 5 (20.8) |
| | | Extreme pain | 4 (8.7) | 6 (10.7) | 4 (16.7) | 2 (8.3) |
| | | Missing | 22 | 18 | 4 | 2 |
| | Day 10 | Total responded | 21 (100.0) | 27 (100.0) | 12 (100.0) | 11 (100.0) |
| | | No pain | 12 (57.1) | 10 (37.0) | 4 (33.3) | 4 (36.4) |
| | | Slight pain | 5 (23.8) | 8 (29.6) | 4 (33.3) | 4 (36.4) |
| | | Moderate pain | 3 (14.3) | 7 (25.9) | 2 (16.7) | 3 (27.3) |
| | | Severe pain | 1 (4.8) | 2 (7.4) | 2 (16.7) | 0 (0.0) |
| | | Extreme pain | 0 (0.0) | 0 (0.0) | 0 (0.0) | 0 (0.0) |
| | | Missing | 40 | 44 | 15 | 12 |
| | Day 28 | Total responded | 22 (100.0) | 30 (100.0) | 12 (100.0) | 10 (100.0) |
| | | No pain | 12 (54.5) | 17 (56.7) | 5 (41.7) | 7 (70.0) |
| | | Slight pain | 4 (18.2) | 2 (6.7) | 1 (8.3) | 0 (0.0) |
| | | Moderate pain | 3 (13.6) | 5 (16.7) | 4 (33.3) | 2 (20.0) |
| | | Severe pain | 2 (9.1) | 4 (13.3) | 0 (0.0) | 0 (0.0) |
| | | Extreme pain | 1 (4.5) | 2 (6.7) | 2 (16.7) | 1 (10.0) |
| | | Missing | 33 | 34 | 14 | 9 |
| | Day 90 | Total responded | 23 (100.0) | 26 (100.0) | 13 (100.0) | 8 (100.0) |
| | | No pain | 7 (30.4) | 8 (30.8) | 3 (23.1) | 1 (12.5) |
| | | Slight pain | 5 (21.7) | 7 (26.9) | 5 (38.5) | 4 (50.0) |
| | | Moderate pain | 5 (21.7) | 4 (15.4) | 3 (23.1) | 2 (25.0) |
| | | Severe pain | 6 (26.1) | 4 (15.4) | 2 (15.4) | 1 (12.5) |
| | | Extreme pain | 0 (0.0) | 3 (11.5) | 0 (0.0) | 0 (0.0) |
| | | Missing | 28 | 33 | 13 | 9 |

*Created using: X:\HAP-FAST\Statistical Analysis - Blinded\Final Analysis\Version 1.0\Programs\Section12-4_EQ5D5L.sas*
*Patients are included if a questionnaire is expected (i.e. they had consented and had not withdrawn at that timepoint).*

Table 12-12 Number and percentage of participants in each severity level for anxiety or depression (baseline, day 10, day 28 and day 90)

| Dimension | Timepoint | Severity level | CT | CXR | FAPP | No FAPP |
|---|---|---|---|---|---|---|
| Anxiety or depression | Baseline | Total responded | 46 (100.0) | 56 (100.0) | 24 (100.0) | 24 (100.0) |
| | | Not anxious or depressed | 18 (39.1) | 22 (39.3) | 9 (37.5) | 6 (25.0) |
| | | Slightly anxious or depressed | 7 (15.2) | 14 (25.0) | 4 (16.7) | 9 (37.5) |
| | | Moderately anxious or depressed | 12 (26.1) | 8 (14.3) | 6 (25.0) | 3 (12.5) |
| | | Severely anxious or depressed | 6 (13.0) | 9 (16.1) | 3 (12.5) | 5 (20.8) |
| | | Extremely anxious or depressed | 3 (6.5) | 3 (5.4) | 2 (8.3) | 1 (4.2) |
| | | Missing | 22 | 18 | 4 | 2 |
| | Day 10 | Total responded | 21 (100.0) | 27 (100.0) | 12 (100.0) | 11 (100.0) |
| | | Not anxious or depressed | 13 (61.9) | 16 (59.3) | 8 (66.7) | 6 (54.5) |
| | | Slightly anxious or depressed | 3 (14.3) | 4 (14.8) | 1 (8.3) | 0 (0.0) |
| | | Moderately anxious or depressed | 4 (19.0) | 4 (14.8) | 2 (16.7) | 5 (45.5) |
| | | Severely anxious or depressed | 1 (4.8) | 3 (11.1) | 1 (8.3) | 0 (0.0) |
| | | Extremely anxious or depressed | 0 (0.0) | 0 (0.0) | 0 (0.0) | 0 (0.0) |
| | | Missing | 40 | 44 | 15 | 12 |
| | Day 28 | Total responded | 22 (100.0) | 30 (100.0) | 12 (100.0) | 10 (100.0) |
| | | Not anxious or depressed | 14 (63.6) | 13 (43.3) | 5 (41.7) | 5 (50.0) |
| | | Slightly anxious or depressed | 5 (22.7) | 8 (26.7) | 3 (25.0) | 4 (40.0) |
| | | Moderately anxious or depressed | 3 (13.6) | 4 (13.3) | 2 (16.7) | 0 (0.0) |
| | | Severely anxious or depressed | 0 (0.0) | 3 (10.0) | 1 (8.3) | 1 (10.0) |
| | | Extremely anxious or depressed | 0 (0.0) | 2 (6.7) | 1 (8.3) | 0 (0.0) |
| | | Missing | 33 | 34 | 14 | 9 |
| | Day 90 | Total responded | 23 (100.0) | 26 (100.0) | 13 (100.0) | 8 (100.0) |
| | | Not anxious or depressed | 14 (60.9) | 13 (50.0) | 9 (69.2) | 3 (37.5) |
| | | Slightly anxious or depressed | 1 (4.3) | 5 (19.2) | 2 (15.4) | 0 (0.0) |
| | | Moderately anxious or depressed | 4 (17.4) | 4 (15.4) | 1 (7.7) | 3 (37.5) |
| | | Severely anxious or depressed | 1 (4.3) | 1 (3.8) | 0 (0.0) | 0 (0.0) |
| | | Extremely anxious or depressed | 3 (13.0) | 3 (11.5) | 1 (7.7) | 2 (25.0) |
| | | Missing | 28 | 33 | 13 | 9 |

*Created using: X:\HAP-FAST\Statistical Analysis - Blinded\Final Analysis\Version 1.0\Programs\Section12-4_EQ5D5L.sas*
*Patients are included if a questionnaire is expected (i.e. they had consented and had not withdrawn at that timepoint).*

##### 12.4.1 VAS score

Figure 12-3 Boxplots of VAS score for each treatment arm (baseline, day 10, day 28 and day 90)

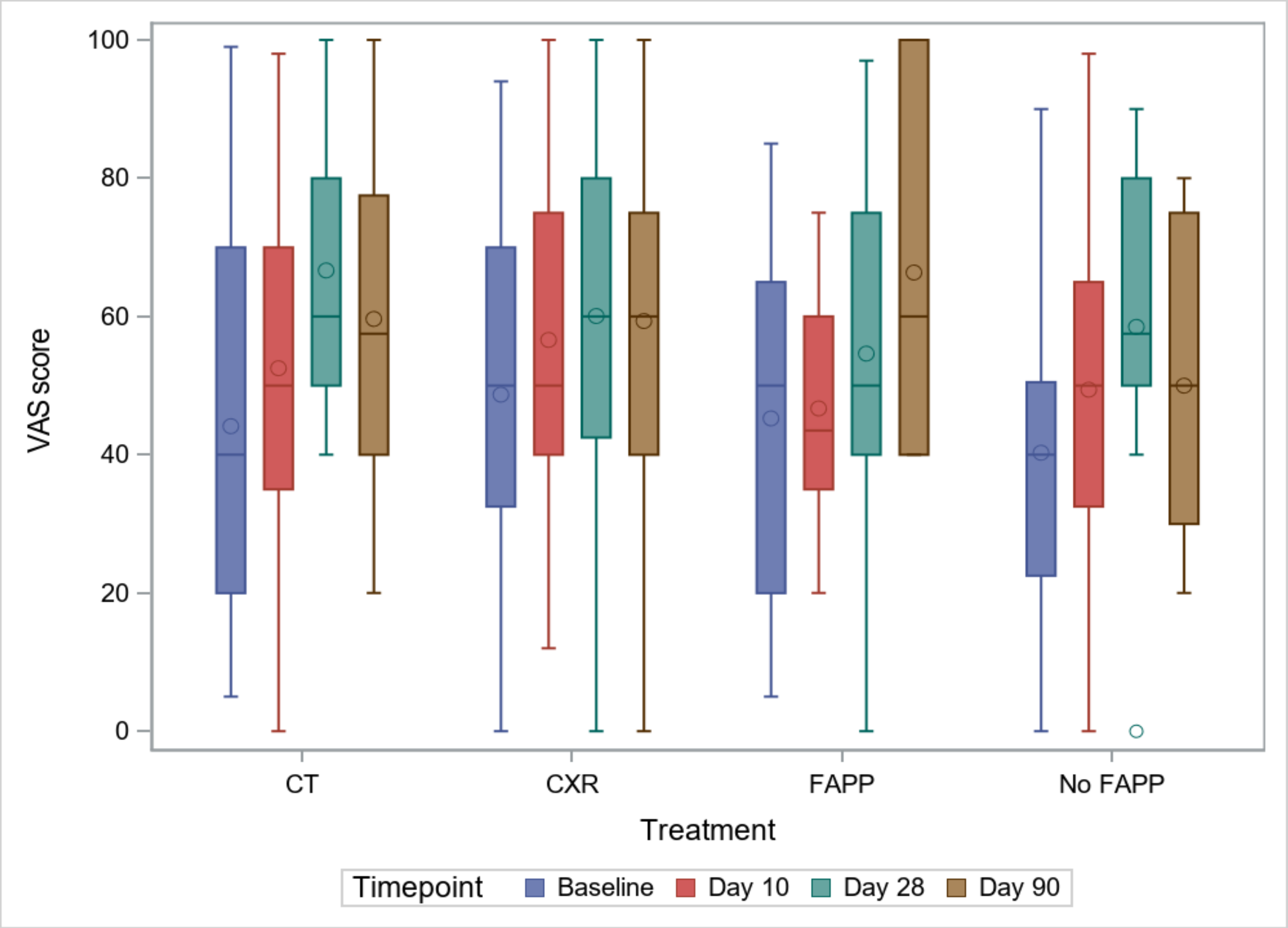

*Created using: X:\HAP-FAST\Statistical Analysis - Blinded\Final Analysis\Version 1.0\Programs\Section12-4_EQ5D5L.sas*
*Patients are included if a questionnaire is expected (i.e. they had consented and had not withdrawn at that timepoint).*

Table 12-13 Summary statistics – VAS score (baseline, day 10, day 28 and day 90)

| Timepoint | Statistic | CT | CXR | FAPP | No FAPP |
|---|---|---|---|---|---|
| Baseline | N | 68 | 74 | 28 | 26 |
| | Mean (SD) | 44.1 (29.0) | 48.7 (26.8) | 45.3 (24.3) | 40.3 (26.4) |
| | Median (IQR) | 40.0 (20.0, 70.0) | 50.0 (32.5, 70.0) | 50.0 (20.0, 65.0) | 40.0 (22.5, 50.5) |
| | Range | (5, 99) | (0, 94) | (5, 85) | (0, 90) |
| | Missing | 23 | 18 | 5 | 2 |
| Day 10 | N | 61 | 71 | 27 | 23 |
| | Mean (SD) | 52.5 (26.2) | 56.6 (24.1) | 46.7 (17.5) | 49.4 (25.5) |
| | Median (IQR) | 50.0 (35.0, 70.0) | 50.0 (40.0, 75.0) | 43.5 (35.0, 60.0) | 50.0 (32.5, 65.0) |
| | Range | (0, 98) | (12, 100) | (20, 75) | (0, 98) |
| | Missing | 40 | 45 | 17 | 11 |
| Day 28 | N | 55 | 64 | 26 | 19 |
| | Mean (SD) | 66.7 (19.4) | 60.1 (28.3) | 54.6 (28.2) | 58.5 (26.7) |
| | Median (IQR) | 60.0 (50.0, 80.0) | 60.0 (42.5, 80.0) | 50.0 (40.0, 75.0) | 57.5 (50.0, 80.0) |
| | Range | (40, 100) | (0, 100) | (0, 97) | (0, 90) |
| | Missing | 34 | 36 | 15 | 9 |
| Day 90 | N | 51 | 59 | 26 | 17 |
| | Mean (SD) | 59.7 (24.4) | 59.3 (25.7) | 66.4 (24.5) | 50.0 (22.9) |
| | Median (IQR) | 57.5 (40.0, 77.5) | 60.0 (40.0, 75.0) | 60.0 (40.0, 100) | 50.0 (30.0, 75.0) |
| | Range | (20, 100) | (0, 100) | (40, 100) | (20, 80) |
| | Missing | 31 | 36 | 15 | 10 |

*Created using: X:\HAP-FAST\Statistical Analysis - Blinded\Final Analysis\Version 1.0\Programs\Section12-4_EQ5D5L.sas*
*Patients are included if a questionnaire is expected (i.e. they had consented and had not withdrawn at that timepoint).*

Figure 12-4 Boxplots of change in VAS score from baseline to day 90 for each treatment arm

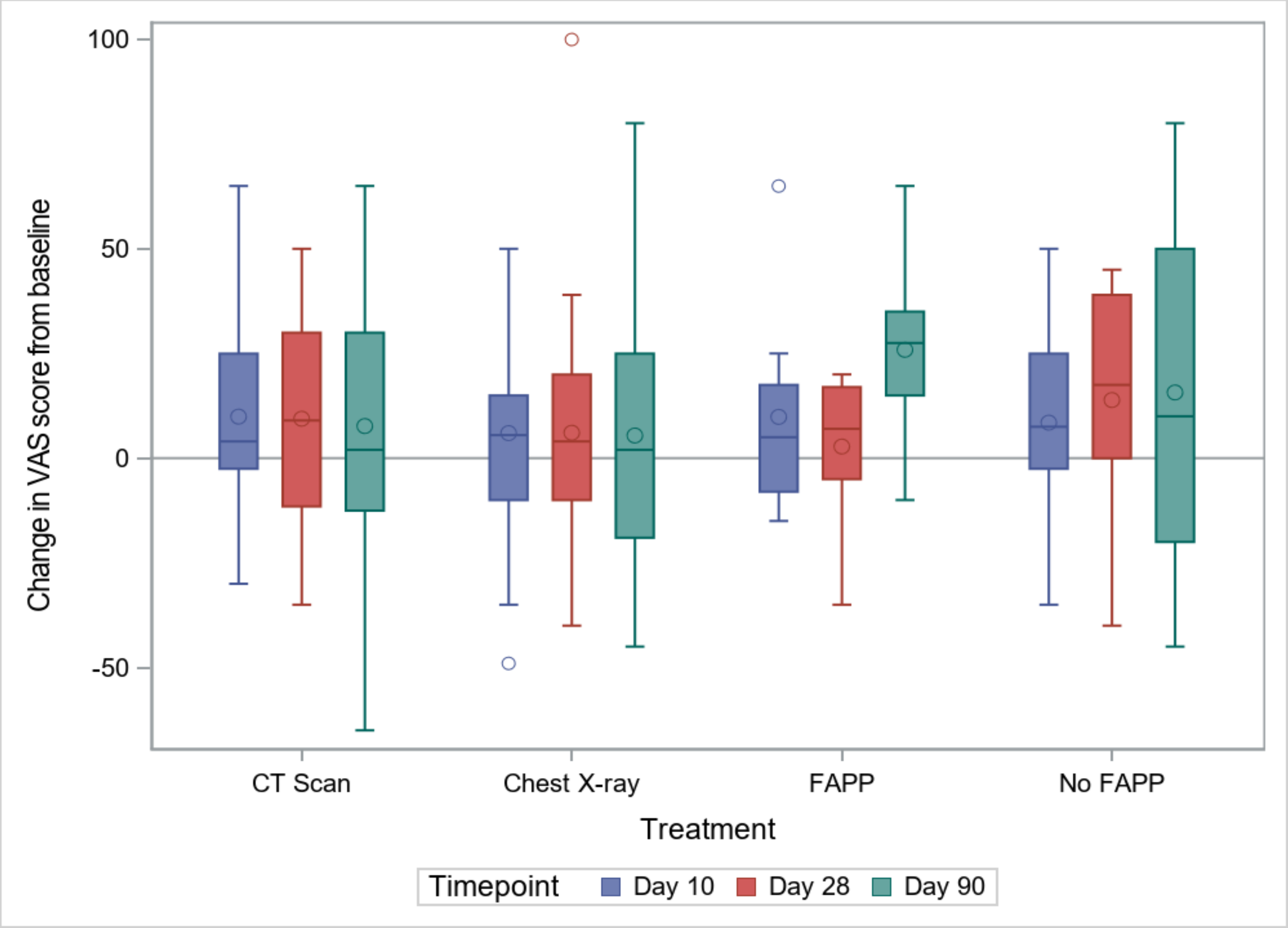

*Created using: X:\HAP-FAST\Statistical Analysis - Blinded\Final Analysis\Version 1.0\Programs\Section12-4_EQ5D5L.sas*
*Patients are included if a questionnaire is expected (i.e. they had consented and had not withdrawn at baseline and the selected timepoint).*

Table 12-14 Summary statistics – Change in VAS score from baseline to day 90. Patients are included if a VAS score is expected (i.e. they had consented and had not withdrawn from the study at day 90).

| Timepoint | Statistic | CT | CXR | FAPP | No FAPP |
|---|---|---|---|---|---|
| Change from baseline to day 90 | N | 51 | 59 | 26 | 17 |
| | Mean (SD) | 7.7 (33.0) | 5.4 (30.8) | 25.9 (21.1) | 15.7 (43.4) |
| | Median (IQR) | 2.0 (-13, 30.0) | 2.0 (-19, 25.0) | 27.5 (15.0, 35.0) | 10.0 (-20, 50.0) |
| | Range | (-65, 65) | (-45, 80) | (-10, 65) | (-45, 80) |
| | Missing | 35 | 38 | 16 | 10 |

*Created using: X:\HAP-FAST\Statistical Analysis - Blinded\Final Analysis\Version 1.0\Programs\Section12-4_EQ5D5L.sas*
*Patients are included if a questionnaire is expected at both baseline and day 90 (i.e. they had consented and had not withdrawn at those timepoints).*

Table 12-15 Linear regression models estimates – Change in VAS score from baseline to day 90

| Comparison | Treatment group | N (Missing) | Mean difference (95% CI) |
|---|---|---|---|
| CT vs CXR | CT | 51 (35) | -0.9 (-16.6, 14.8) |
| | CXR | 59 (38) | |
| FAPP vs No FAPP | FAPP | 26 (16) | 17.7 (-6.0, 41.4) |
| | No FAPP | 17 (10) | |

*Created using: X:\HAP-FAST\Statistical Analysis - Blinded\Final Analysis\Version 1.0\Programs\Section12-4_EQ5D5L.sas*
*Patients are included if a questionnaire is expected at both baseline and day 90 (i.e. they had consented and had not withdrawn at those timepoints).*

#### 12.5 Secondary outcome 4 – Length of hospital stay post HAP diagnosis

##### 12.5.1 Time to event

Table 12-16 Summary statistics – Length of stay in hospital from suspected HAP.

| | CT | CXR | FAPP | No FAPP |
|---|---|---|---|---|
| N | 109 | 111 | 31 | 31 |
| Mean (SE) | 53.5 (8.5) | 21.1 (2.1)[19] | 14.5 (2.4) | 18.2 (2.4)[19] |
| Median (IQR) | 19 (8, 52) | 13 (6, 42) | 9 (7, 16) | 17 (9, †) |
| Range | (0, 176) | (0, †) | (0, 45) | (2, †) |

*Created using: X:\HAP-FAST\Statistical Analysis - Blinded\Final Analysis\Version 1.0\Programs\Section12-5_LengthOfStay.sas*

Figure 12-5 Kaplan-Meier plots of length of hospital stay following suspected HAP for each treatment arm

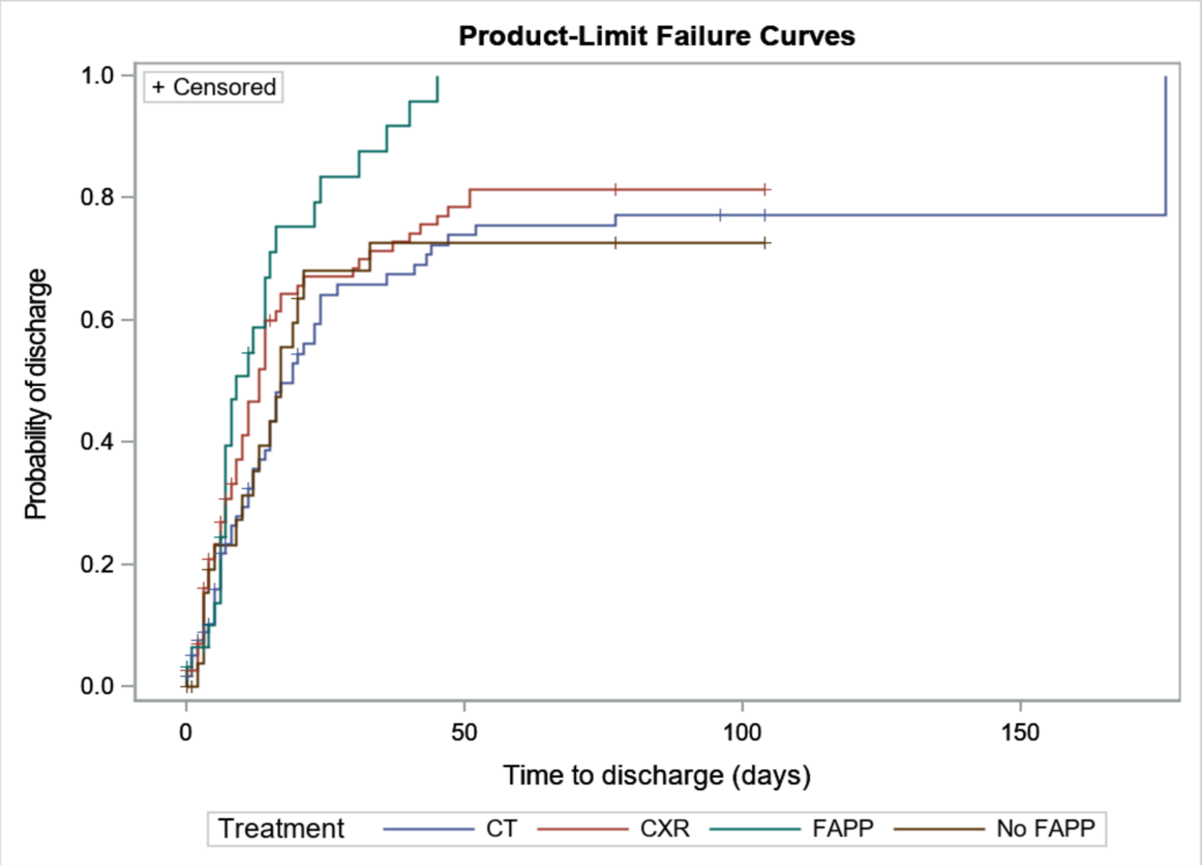

*Created using: X:\HAP-FAST\Statistical Analysis - Blinded\Final Analysis\Version 1.0\Programs\Section12-5_LengthOfStay.sas*

[19] This estimated mean is biased towards zero because the latest observation is censored.
† Quantity cannot be estimated due to censoring of later observations.

#### 12.6 Secondary outcome 5 – Mortality

Figure 12-6 Kaplan-Meier plots of survival following suspected HAP for each treatment arm

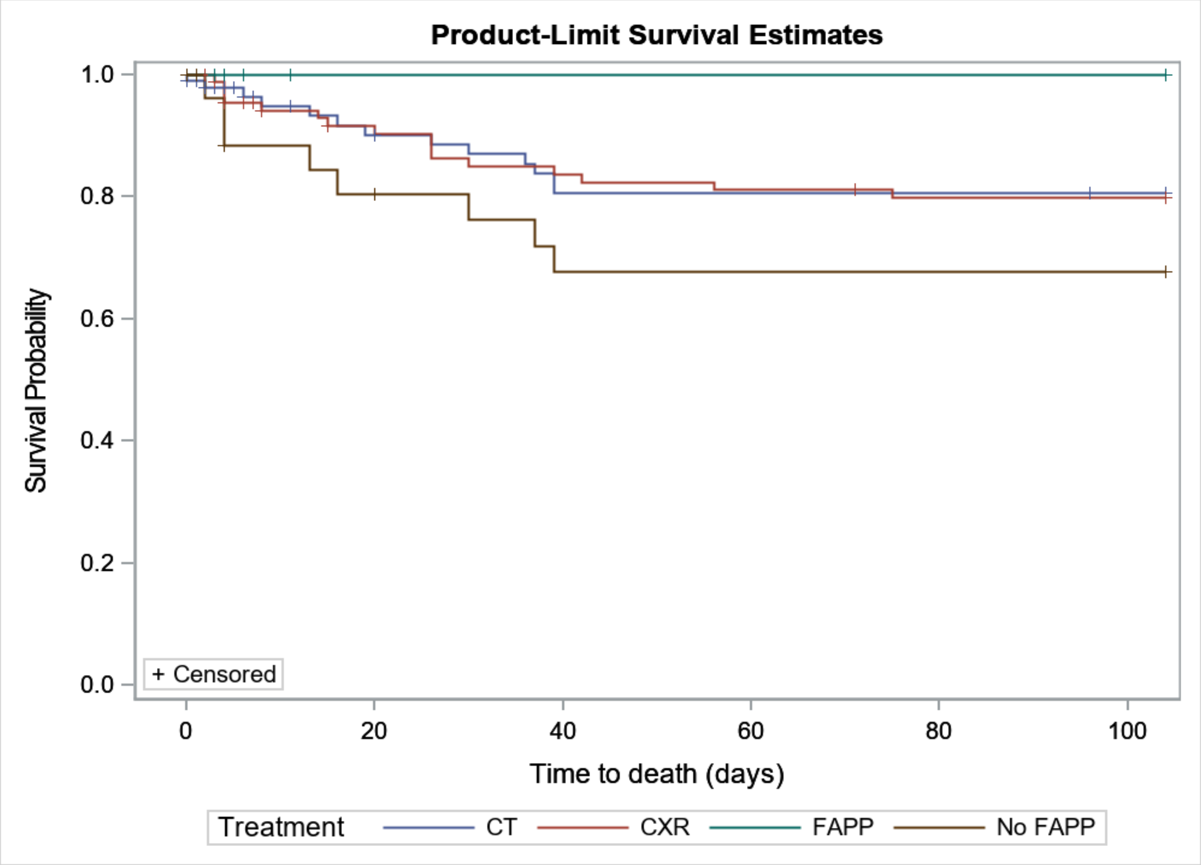

*Created using: X:\HAP-FAST\Statistical Analysis - Blinded\Final Analysis\Version 1.0\Programs\Section12-6_Mortality.sas*

Table 12-17 Kaplan-Meier survival rates and 95% confidence intervals at days 14, 28 and 90, by treatment group

| | | CT | CXR | FAPP | No FAPP |
|---|---|---|---|---|---|
| Day 14 | N total | 109 | 111 | 31 | 31 |
| | Surviving | 60 | 73 | 26 | 21 |
| | Died | 5 | 6 | 0 | 4 |
| | Censored | 44 | 32 | 5 | 6 |
| | Survival rate | 93.3 (84.4, 97.2) | 92.9 (84.8, 96.7) | 100.0 (100.0, 100.0) | 84.4 (63.7, 93.9) |
| Day 28 | N total | 109 | 111 | 31 | 31 |
| | Surviving | 56 | 66 | 26 | 19 |
| | Died | 8 | 11 | 0 | 5 |
| | Censored | 45 | 34 | 5 | 7 |
| | Survival rate | 88.6 (78.3, 94.2) | 86.4 (76.7, 92.2) | 100.0 (100.0, 100.0) | 80.4 (59.1, 91.4) |
| Day 90 | N total | 109 | 111 | 31 | 31 |
| | Surviving | 51 | 60 | 26 | 16 |
| | Died | 13 | 16 | 0 | 8 |
| | Censored | 45 | 35 | 5 | 7 |
| | Survival rate | 80.7 (68.9, 88.4) | 79.8 (69.1, 87.1) | 100.0 (100.0, 100.0) | 67.7 (45.6, 82.4) |

*Created using: X:\HAP-FAST\Statistical Analysis - Blinded\Final Analysis\Version 1.0\Programs\Section12-6_Mortality.sas*

Table 12-18 Difference in Kaplan-Meier survival rates between treatment arms, percentage points, with 95% confidence intervals.

| | CT vs CXR | FAPP vs No FAPP |
|---|---|---|
| Day 14 | 0.4 (-7.6, 8.4) | 15.6 (1.5, 29.6) |
| Day 28 | 2.2 (-8.4, 12.9) | 19.6 (4.2, 35.0) |
| Day 90 | 0.9 (-12.1, 13.9) | 32.3 (13.8, 50.8) |

*Created using: X:\HAP-FAST\Statistical Analysis - Blinded\Final Analysis\Version 1.0\Programs\Section12-6_Mortality.sas*

#### 12.7 Secondary outcome 6 – Completion of outcomes

219 (99.5%) participants could not be assessed for clinical cure:

- 68 (31.1%) discharged without repeat chest imaging
- 9 (4.1%) died without repeat chest imaging
- 90 (41.1%) did not consent by day 3
- 8 (3.7%) declined consent between day 4 and day 6
- 59 (26.9%) withdrew consent before day 7
- 215 (98.2%) had no repeat chest imaging up to day 10
- 219 (100.0%) had incomplete data[20] (see below).

Table 12-19 Time to clinical cure missing data

| Incomplete data | Missing data n (%) |
|---|---|
| Incomplete data | 219 (100.0) |
| Sign/ symptom/ radiological sign missing[21]: | |
| Temperature | 28 (12.8) |
| Heart rate | 28 (12.8) |
| Respiratory rate | 28 (12.8) |
| Systolic blood pressure | 28 (12.8) |
| Oxygen saturation on room air | 40 (18.3) |
| Oxygen saturation on room air (pre-HAP state)[22] | 47 (21.5) |
| Ability to eat and drink independently | 67 (30.6) |
| Ability to eat and drink independently (pre-HAP state)[22] | 68 (31.1) |
| Cognitive function | 76 (34.7) |
| Radiological scan result[23] | 218 (99.5) |
| CAP-Sym | 118 (53.9) |
| CAP-Sym (baseline not completed within 3 days) | 117 (53.4) |

*Created using: X:\HAP-FAST\Statistical Analysis - Blinded\Final Analysis\Version 1.0\Programs\Section12-7_CompletenessTTC.sas*

[20] Participants may be assessed for clinical cure multiple times (up to day 10). If they never had complete data to assess clinical cure on any of these days, they are classed as having incomplete data.

[21] Only the participant's least missing assessment up to day 10, i.e. the day that has the least components missing, is included in this table. Participants may have more than one sign/ symptom/ radiological sign missing.

[22] For some participants this data may be not known at admission. The clinical cure criteria for oxygen saturation (≥ 90% on room air or return to pre-HAP state) and ability to eat and drink independently (yes or return to pre-HAP state) are not fully dependent on the pre-HAP state being collected so participants may still meet the clinical cure criteria without admission data for oxygen saturation and ability to eat and drink independently.

[23] Further imaging and findings are collected on the Day 90 visit eCRF. Only imaging carried out up to day 10 is applicable to this outcome. If the participant does not have a Day 90 eCRF or their eCRF indicates that they had further imaging up to day 10 and the finding(s) are not recorded on the eCRF, they will be considered as missing their radiological scan result for that day.

##### 12.7.1 Questionnaires

In the following tables:

- E = Expected – questionnaire is expected if the participant has not declined consent prior to that timepoint.
- A = Actual – questionnaire is classed as actually returned if at least one question has been answered.
- C = Completed – questionnaire has been completed to an analysable standard and within the visit window.

Table 12-20 EQ-5D-5L return and completion rates, n (%)

| | **Baseline** | | | **Day 10 or discharge (if discharged before day 10)** | | | **Day 28**[24] | | | **Day 90**[25] | | |
|---|---|---|---|---|---|---|---|---|---|---|---|---|
| | **E** | **A** | **C** | **E** | **A** | **C** | **E** | **A** | **C** | **E** | **A** | **C** |
| **EQ-5D-5L**[26]**:** | 142 (100.0) | 104 (73.2) | | 132 (100.0) | 49 (37.1) | | 119 (100.0) | 52 (43.7) | | 110 (100.0) | 49 (44.5) | |
| Mobility | | | 102 (71.8) | | | 48 (36.4) | | | 52 (43.7) | | | 49 (44.5) |
| Self-care | | | 102 (71.8) | | | 48 (36.4) | | | 52 (43.7) | | | 49 (44.5) |
| Usual activities | | | 102 (71.8) | | | 48 (36.4) | | | 52 (43.7) | | | 49 (44.5) |
| Pain/ Discomfort | | | 102 (71.8) | | | 48 (36.4) | | | 52 (43.7) | | | 49 (44.5) |
| Anxiety/ Depression | | | 102 (71.8) | | | 48 (36.4) | | | 52 (43.7) | | | 49 (44.5) |
| VAS score | | | 101 (71.1) | | | 47 (35.6) | | | 49 (41.2) | | | 43 (39.1) |

*Created using: X:\HAP-FAST\Statistical Analysis - Blinded\Final Analysis\Version 1.0\Programs\Section12-7_CompletenessEQ5D.sas*

[24] The 28-day follow-up questionnaire should be completed within the visit window for the follow-up (28 days +/- 7 days).
[25] The 90-day follow-up questionnaire should be completed within the visit window for the follow-up (90 days +/- 14 days).
[26] Questionnaires can only be completed after consent so only consenting participants are expected to have questionnaires at each timepoint.

Table 12-21 CAP-Sym return and completion rates, n (%)

| | **CAP-Sym[27]** | | | | |
|---|---|---|---|---|---|
| | **E** | **A** | **C** | **Number of missing items (for questionnaires actually returned)** | |
| | | | | **Mean (SD)** | **Median (IQR)** |
| Baseline | 74 (100.0) | 66 (89.2) | 66 (89.2) | 0.0 (0.0) | 0 (0, 0) |
| Day 1 | 102 (100.0) | 52 (51.0) | 50 (49.0) | 0.3 (2.2) | 0 (0, 0) |
| Day 2 | 113 (100.0) | 54 (47.8) | 53 (46.9) | 0.1 (0.4) | 0 (0, 0) |
| Day 3 | 117 (100.0) | 70 (59.8) | 69 (59.0) | 0.0 (0.2) | 0 (0, 0) |
| Day 4 | 112 (100.0) | 63 (56.3) | 62 (55.4) | 0.0 (0.1) | 0 (0, 0) |
| Day 5 | 109 (100.0) | 55 (50.5) | 55 (50.5) | 0.0 (0.0) | 0 (0, 0) |
| Day 6 | 103 (100.0) | 60 (58.3) | 57 (55.3) | 0.1 (0.7) | 0 (0, 0) |
| Day 7 | 95 (100.0) | 61 (64.2) | 60 (63.2) | 0.2 (1.3) | 0 (0, 0) |
| Day 8 | 90 (100.0) | 46 (51.1) | 46 (51.1) | 0.0 (0.0) | 0 (0, 0) |
| Day 9 | 87 (100.0) | 38 (43.7) | 38 (43.7) | 0.0 (0.0) | 0 (0, 0) |
| Day 10 | 83 (100.0) | 42 (50.6) | 42 (50.6) | 0.0 (0.0) | 0 (0, 0) |
| Day 10 or discharge (if discharged before day 10) | 132 (100.0) | 73 (55.3) | 72 (54.5) | 0.2 (1.9) | 0 (0, 0) |
| Day 28[28] | 119 (100.0) | 52 (43.7) | 50 (42.0) | 0.1 (0.6) | 0 (0, 0) |
| Day 90[29] | 110 (100.0) | 51 (46.4) | 49 (44.5) | 0.4 (2.4) | 0 (0, 0) |

*Created using: X:\HAP-FAST\Statistical Analysis - Blinded\Final Analysis\Version 1.0\Programs\Section12-7_CompletenessCapSym.sas*

[27] Questionnaires can only be completed after consent so only consenting participants are expected to have questionnaires at each timepoint. Daily CAP-Sym questionnaires are only expected on days before the patient is discharged.
[28] The 28-day follow-up questionnaire should be completed within the visit window for the follow-up (28 days +/- 7 days).
[29] The 90-day follow-up questionnaire should be completed within the visit window for the follow-up (90 days +/- 14 days).

Table 12-22 Post-discharge indirect cost survey return and completion rates

| | Day 90[30] | | |
|---|---|---|---|
| | **E[31]** | **A** | **C** |
| **Post-discharge indirect cost survey[32]:** | 110 | 55 (50.0) | |
| Since your discharge from hospital, have you had a GP appointment? | | | 52 (47.3) |
| If yes, how many appointments? | 29 | | 28 (96.6) |
| What were the reasons for these appointments? | 29 | | 28 (96.6) |
| Since your discharge, have you had to go back to hospital? | | | 55 (50.0) |
| What were your symptoms that prompted you to go back to hospital? | 29 | | 29 (100.0) |
| How long were you in hospital for? | 29 | | 26 (89.7) |
| Since your discharge from hospital, have you had any further investigations (for example blood tests, scans, breathing tests or camera tests)? | | | 53 (48.2) |
| Do you know why the doctor ordered these tests? | 35 | | 34 (97.1) |
| After you left hospital did you go to a respite or rehabilitation bed? | | | 54 (49.1) |
| If yes, what kind of facility did you go to? | 8 | | 8 (100.0) |
| How many days were you there? | 8 | | 2 (25.0) |
| Since your discharge, have you gone to a hospital clinic appointment? | | | 53 (48.2) |
| If yes, what was the reason for the clinic appointment? | 23 | | 23 (100.0) |
| Have you had any ***NEW*** help from the following community services? | N/A – only collected if applicable | | |
| How long do their visits last? | 47 | | 25 (53.2) |
| How many times a week do they come to help? | 47 | | 21 (44.7) |
| What is the reason you need this help? | 47 | | 36 (76.6) |
| Since your discharge from hospital, have you started taking any ***NEW*** medications prescribed by your GP? | | | 46 (41.8) |
| If yes, what were these medications? | 11 | | 11 (100.0) |
| Course length (if long term, please leave blank) | N/A – only collected if not long term | | |

[30] The 90-day follow-up questionnaire should be completed within the visit window for the follow-up (90 days +/- 14 days).
[31] Some questions are only expected depending on the answer to a previous question. These conditional questions have an expected number of answers shown, other questions have a grey box.
[32] The survey overall is expected for all patients who had consented and not withdrawn at day 90.

| | | | |
|---|---|---|---|
| Have you missed work due to being ill since your discharge from hospital? | | | 41 (37.3) |
| If yes, how many days have you missed? | 5 | | 5 (100.0) |
| How much do you earn an hour? Approximately (£) | 5 | | 3 (60.0) |
| How many hours do you work in a normal working day? | 5 | | 5 (100.0) |
| What is the reason you had time off work? | 5 | | 5 (100.0) |
| Since your discharge from hospital, have friends or family had to take time off work to help you? | | | 50 (45.5) |
| If yes, how many days have they missed? | 11 | | 9 (81.8) |
| How much do they earn an hour? Approximately (£) | 11 | | 5 (45.5) |
| How many hours do they work in a normal working day? | 11 | | 5 (45.5) |
| What is the reason you need their help? | 11 | | 5 (45.5) |

*Created using: X:\HAP-FAST\Statistical Analysis - Blinded\Final Analysis\Version 1.0\Programs\Section12-7_CompletenessCosts.sas*

#### 12.8 Secondary outcome 7 – Eligibility, recruitment and consent rates

##### 12.8.1 Recruitment

Table 12-23 Rate of recruitment

| | Total number recruited | Total number of months with site(s) open | Recruitment rate/month |
|---|---|---|---|
| **Overall** | **220** | **9** | **24.4** |
| Aintree (01) | 117 | 9 | 13.0 |
| Royal Liverpool (02) | 34 | 9 | 3.8 |
| Royal Preston (03) | 32 | 8 | 4.0 |
| Wythenshawe (04) | 37 | 8 | 4.6 |

*Created using: X:\HAP-FAST\Statistical Analysis - Blinded\Final Analysis\Version 1.0\Programs\Section12-8_Rates.sas*

##### 12.8.2 Eligibility

Table 12-24 Proportion screened that meet eligibility criteria

*Table12-24 cannot be created because data on number of HAP cases is not available.*

*Created using: X:\HAP-FAST\Statistical Analysis - Blinded\Final Analysis\Version 1.0\Programs\Section12-8_Rates.sas*

##### 12.8.3 Consent rates

Table 12-25 Proportion eligible that consent and where they present

| Ward type | Total number eligible | Total number consented | Proportion |
|---|---|---|---|
| **Overall** | **220** | **142** | **64.5%** |
| Medical | 96 | 65 | 67.7% |
| Surgical | 25 | 20 | 80.0% |
| A&E | 62 | 49 | 79.0% |
| Acute medical unit | 6 | 5 | 83.3% |
| Other | 4 | 3 | 75.0% |
| Not known | 27 | 0 | 0.0% |

*Created using: X:\HAP-FAST\Statistical Analysis - Blinded\Final Analysis\Version 1.0\Programs\Section12-8_Rates.sas*

#### 12.9 Secondary outcome 8 – Rates of successful follow-up

##### 12.9.1 Completed the full study pathway as per protocol

Table 12-26 Proportion consented and randomised that complete the full study pathway as per protocol

| Treatment arm | Total number completing study pathway | Total number consented and randomised | Proportion |
|---|---|---|---|
| **Overall** | **28** | **142** | **19.7%** |
| CT Scan | 13 | 68 | 19.1% |
| Chest X-ray | 15 | 74 | 20.3% |
| FAPP | 8 | 28 | 28.6% |
| No FAPP | 4 | 26 | 15.4% |

*Created using: X:\HAP-FAST\Statistical Analysis - Blinded\Final Analysis\Version 1.0\Programs\Section12-9_Pathways.sas*

##### 12.9.2 Withdrew from study intervention or follow up

Table 12-27 Proportion consented and randomised that withdrew from study intervention or follow up

| Treatment arm | Total number withdrew | Total number consented and randomised | Proportion |
|---|---|---|---|
| **Overall** | **25** | **142** | **17.6%** |
| CT Scan | 16 | 68 | 23.5% |
| Chest X-ray | 9 | 74 | 12.2% |
| FAPP | 8 | 28 | 28.6% |
| No FAPP | 2 | 26 | 7.7% |

*Created using: X:\HAP-FAST\Statistical Analysis - Blinded\Final Analysis\Version 1.0\Programs\Section12-9_Pathways.sas*

#### 12.10 Secondary outcome 9 – Summary statistics relating to antibiotic use in the pilot study with a comparison between the DTRs

Table 12-28 Amount of antibiotics given for treatment of HAP/RTI, by treatment pathway, in defined daily doses (DDD).

| | CXR Sputum FAPP | CXR Sputum No FAPP | CXR No Sputum No FAPP | CT Sputum FAPP | CT Sputum No FAPP | CT No sputum No FAPP |
|---|---|---|---|---|---|---|
| N | 15 | 16 | 50 | 16 | 15 | 42 |
| Mean (SD) | 6.0 (6.0) | 4.5 (5.2) | 6.0 (12.0) | 8.2 (12.1) | 6.9 (11.7) | 3.8 (4.4) |
| Median (IQR) | 5.1 (0.3, 9.7) | 4.0 (0.0, 8.4) | 1.1 (0.0, 6.3) | 4.1 (0.0, 10.0) | 1.8 (0.0, 8.9) | 2.2 (0.0, 6.6) |
| Range | (0, 19) | (0, 16) | (0, 65) | (0, 42) | (0, 34) | (0, 17) |
| Missing | 3 | 1 | 4 | 4 | 7 | 6 |

*Created using: X:\HAP-FAST\Statistical Analysis - Blinded\Final Analysis\Version 1.0\Programs\Section12-10_AntibioticsSummary.sas*

Figure 12-7 Amount of antibiotics given for treatment of HAP/RTIA, by dynamic treatment regimen, in daily defined doses (DDD).

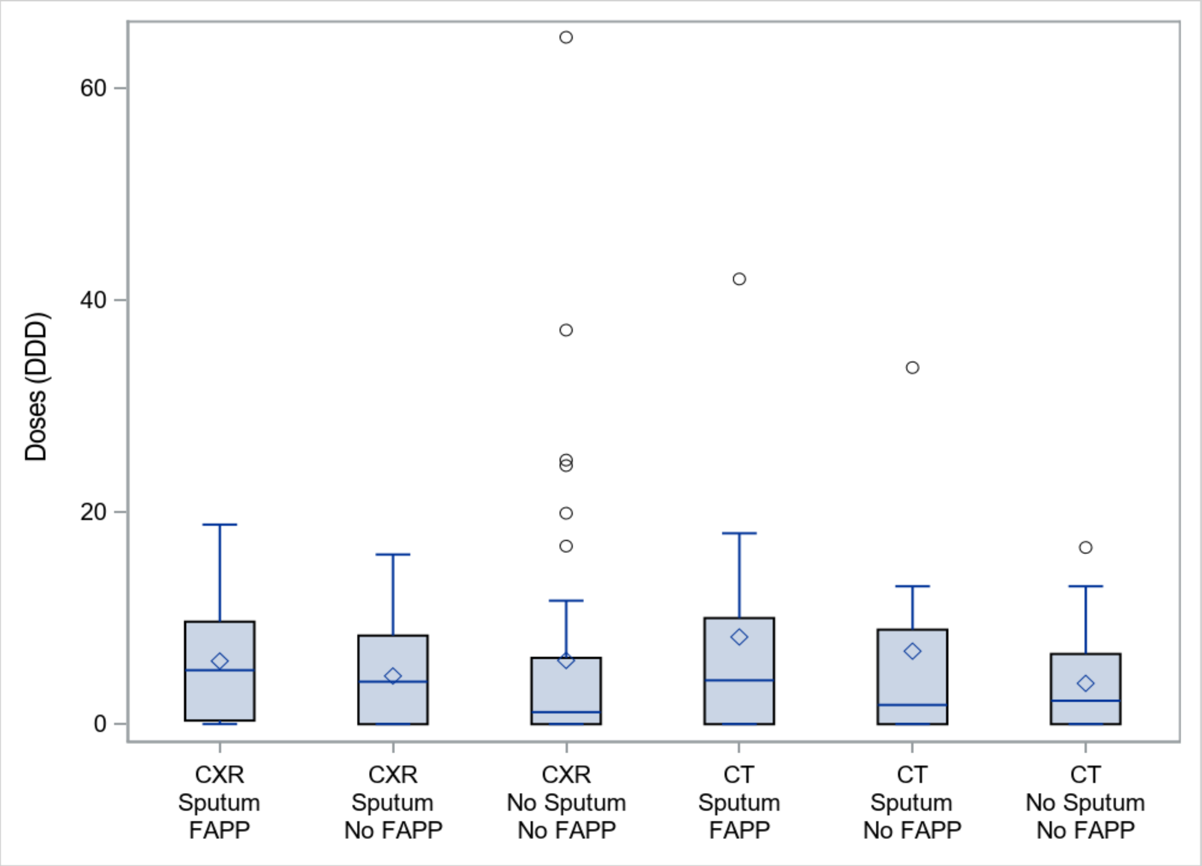

*Created using: X:\HAP-FAST\Statistical Analysis - Blinded\Final Analysis\Version 1.0\Programs\Section12-10_AntibioticsSummary.sas*

### 13 Safety Data

Since HAP-FAST only incorporates well-established and non-invasive diagnostic investigations that would normally be carried out as standard of care, safety events are not recorded as part of the study.

#### 13.1 Non-serious adverse events

Not applicable.

#### 13.2 Serious adverse events

Not applicable.

### 14 Lay Summary of Study Results

#### 14.1 Trial delivery and processes

HAP-FAST opened on the 6th June 2023. Patients were randomised at four hospitals, Aintree, Royal Liverpool, Royal Preston and Wythenshawe at an average of 24.4 patients per month.
The trial closed 3 months ahead of schedule having reached its target of 220 randomised patients by 12th March 2024. In common with many trials, the lead site Aintree recruited half of the patients with the other half being recruited evenly by the other hospitals.
62/220 (28%) of the patients who had stage 1 randomisation (chest x-ray (CXR) versus CT scan) went on to stage 2 randomisation (Film Array Pneumonia Panel (FAPP) versus no FAPP). The reason fewer participants were randomised at stage 2 is that many patients with pneumonia could not produce the sputum sample needed for FAPP analysis. One hospital struggled to implement the FAPP test, so most FAPP data is from the other three hospitals.
65% of those randomised gave consent to ongoing participation in the trial. Of those who did not provide consent common reasons were the burden of blood sampling and trial visits and some patients were upset by a recent cancer diagnosis.
The average age of patients recruited was 77 years which is the same as “real life” audits of HAP in the NHS. Patient characteristics were similar between the groups following both stages of randomisation. This is good evidence that the randomisation process worked and reassuringly there were very few technical issues using the online randomisation process.
Some participants had missing data relating to a small number of the clinical parameters that site teams were asked to collect from the medical record. The trial team will need to consider if those parameters are essential in a future trial and if so will need to put in processes and training to reduce the proportion with missing data.
There were no serious protocol breaches. However, a large majority of participants missed one or more timepoints for data collection. A future trial would likely benefit from a simplified protocol with a reduced number of visits.
Of the participants who were initially randomised to have a CT scan some could not receive the scan and had a CXR instead. The reasons for this were a combination of operational restraints relating to access to the scanner and patients being unable to tolerate lying flat in the scanner.
Of the participants who were initially randomised to have their sputum investigated by FAPP, some did not yield useful results, either because their sample went missing, or the test failed for technical reasons. These issues relating to delivering the trial interventions will need to be borne in mind when considering the size of a future trial and any training sites might need to run the trial.
CT scans and their reports were delivered quickly, and this is a real positive for a future trial as it suggests it would be safe and that any beneficial effect of scans would not be diluted by slow scanning and reporting.

#### 14.2 Clinical effects (outcomes) of the trial

##### 14.2.1 Cure

An attempt was made to analyse the proportion of patients who were cured of their pneumonia. We initially used a definition of cure that included three components:
improvement in patients’ symptoms
improvement in measured clinical observations (for example blood pressure, temperature and breathing rate)
improvement (or lack of deterioration) of patient’s radiology (CXR or CT scan)

The above definition of cure was applied daily up to day 10 following the diagnosis of HAP. We applied this definition because a published paper suggested that the consensus opinion of some experts was that it was likely to be a useful one.

However, we discovered that using the above definition only one of our participants could be assessed for cure as almost all patients had incomplete data. The overriding reason for data being missing was that a repeat CXR or CT scan was not performed and so the patients could not be assessed for improvement in their radiology. This finding reflects normal clinical practice where doctors don't repeat CXRs until about 6-12 weeks after an episode of pneumonia.

We therefore reanalysed the data using a revised definition of clinical cure which included only the first two components – symptoms and clinical observations.

Using this new definition, we were able to assess clinical cure in 36% of participants. This was an improvement upon the original definition but still meant most participants could not be assessed. This was mainly due to some components of the clinical observations not being available or one or both key timepoints for recording symptoms being missed. This will need careful thought and revision for a future trial to ensure that a representative sample of participants can be assess for this key parameter.

Those randomised to the FAPP were more likely to be cured by day 10 than those randomised to no FAPP. Those who were randomised to the CT scan were more likely to be cured than those randomised to CXR.

Overall rates of cure by day 10 were low based on the new definition. It is likely that for a future trial the threshold of 35% improvement in symptoms we chose was too stringent and that either a longer timepoint for symptom assessment should be chosen or a smaller improvement in symptoms at day 10.

##### 14.2.2 Antibiotic use

Being randomised to CT scan meant that, on average, it was more common for antibiotics to be withheld compared to participants who were randomised to a CXR. However, in the CT group, if antibiotics were given, then those participants, on average, received slightly more antibiotic than those who were given antibiotics in the CXR group. The overall effect of CT scans was to slightly reduce the amount of antibiotic used compared to CXR.

Being randomised to FAPP made it less likely that participants' antibiotics were stopped when compared to participants who were randomised to no FAPP. However, if antibiotics were continued then those participants received, on average, fewer antibiotic doses than those randomised to no FAPP. The overall effect of using the FAPP was to increase the amount of antibiotic compared to not using the FAPP.

##### 14.2.3 Quality of life

HAP-FAST wanted to assess if a questionnaire called the EQ-5D-5L could be used to measure changes in patients' quality of life in patients suspected of HAP and if enough participants would respond to provide an assessment of changes in quality of life that might be caused by the different tests used in the trial.

Among randomised patients, roughly half filled out the questionnaire at baseline, 21% at day 10, 23% at day 28, and 22% on day 90. Part of this decrease is due to mortality, or patients withdrawing consent during follow-up. Among patients alive and consenting at each of those timepoints, between 36% and 73% completed the questionnaire. These data are useful for planning the size of a future trial using this questionnaire and they suggest that day 10 or around discharge from hospital is a particularly challenging timepoint. It may be that this timepoint should be dropped or that specific emphasis be put on trying to focus resource to help participants complete the questionnaire then.

Among those who did complete the EQ-5D-5L questionnaire, there was evidence to suggest that it appropriately describes improvement in quality of life during treatment for suspected HAP.

###### 14.2.4 Length of hospital stay

Of the 220 participants, those who were investigated with CT scan had a longer length of stay compared to those who had a CXR. Of the 62 participants who were randomised between FAPP and no FAPP, those receiving the FAPP had the shortest length of stay of all.

###### 14.2.5 Deaths

Comparing all 220 randomised participants, at every timepoint following enrolment (14, 28 and 90 days) there were fewer deaths in the CT group compared to the CXR group. Since the number of participants in this study was small, we did not employ a statistical test to this difference in deaths – but it was small and whilst it may be confirmed by future larger studies, it could also have occurred by chance.

Following stage 2 randomisation, of the 31 patients randomised to FAPP there were no deaths at any timepoint (14, 28 and 90 days) compared to 4/31, 5/31 and 8/31 deaths in the no FAPP group. Thus we observed that whilst all 31 patients randomised to FAPP survived until withdrawal of consent or day 90 only two thirds of patients (67.7%) without FAPP investigation survived to day 90

##### 14.3 Feasibility of a future trial or trials

The trial groups decision about whether a future trial of this nature might be feasible will be based on three broad assessments: -

Firstly, did we manage to recruit enough patients during the planned timeframe, were we able to capture the relevant data and was the experience of the trial acceptable for patients and the staff who had to deliver it.

Secondly, we will need to show a “signal of efficacy” to convince a funder that a future trial is worth doing. A signal or signals of efficacy means evidence that the tests we trialled might lead to meaningful benefits for patients.

The third and final assessment of feasibility relates to the size of a future trial.

Regarding the first of these feasibility assessments, the data presented here is favourable. It demonstrates that patients can be recruited to this protocol at a rate that exceeded our predictions. However, some hospitals were able to recruit faster than others so we will have to be cautious with future assumptions of recruitment rates. The data demonstrates that the CT intervention is applicable to a much wider proportion of patients suspected of HAP than the FAPP intervention – because only a third of patients can produce a sputum sample. That will need to be factored in when planning a future trial. Moreover, some of the outcome measures we chose to evaluate here, for example clinical cure, need adjusting to make them feasible to implement in a larger trial - and the frequency and timing of their administration should be adjusted to minimise the amount of missing data. In terms of acceptability – this cannot be assessed from this quantitative data and will be determined by qualitative analysis of our interviews and focus groups.

Regarding the signals of efficacy, we found evidence that both CT scans and the FAPP alter the total amount of antibiotics prescribed – but further analysis of the types of antibiotics used is needed to complete this assessment. Potential benefits were suggested in length of hospital stay, clinical cure and survival and these results are in favour of conducing a fully powered future trial of bother interventions of two separate single intervention trials.

The size of a future trial will be perhaps the most significant factor in deciding the feasibility of a future trial. Once we have decided on the outcome measure, we should use for a future trial we can use the data presented here to estimate the required sample size we would need to recruit. This factor will be the focus of our trial groups planning going forward.

### 15 Mapping between report shell and SAP

This report has been created following the HAP-FAST Statistical Analysis Plan V1.0 (dated 01/11/2024).

The following table lists each item (tables, figures and section when applicable) in this report and maps each to the relevant SAP section that describes the methods used to compute it.

Table 15-1 Mapping between report shell and SAP

| Section/subsection of SAP | Item within report | Additional details (if required) |
|---|---|---|
| Section 19.1: Screening, eligibility and recruitment | Table 4-1 to Table 4-10<br>Figure 4-1 | |
| Section 19: Disposition of Participants | Figure 3-1 and Figure 3-2 | |
| Section 19.2: Post randomisation discontinuations | Table 7-1 and Table 7-2 | |
| Section 23: Baseline Characteristics | Table 8-1 to Table 8-5 | |
| Section 22: Analysis Datasets | Table 9-1 | |
| Section 20: Protocol Deviations | Table 10-1 | |
| Section 24: Compliance with Intervention | Table 11-1 to Table 11-3 | |
| Section 25.2.3.1: Secondary outcome 1: Time to clinical cure | Table 12-1 to Table 12-5<br>Figure 12-1 | |
| Section 25.2.3.2: Secondary outcome 2: Antibiotic usage for the HAP episode | Table 12-6 and Table 12-7<br>Figure 12-2 | |
| Section 25.2.3.3: Secondary outcome 3: EQ-5D-5L | Table 12-8 to Table 12-15<br>Figure 12-3 and Figure 12-4 | |
| Section 25.2.3.4: Secondary outcome 4: Length of hospital stay post HAP diagnosis | Table 12-16<br>Figure 12-5 | |
| Section 25.2.3.5: Secondary outcome 5: Mortality | Table 12-17 and Table 12-18<br>Figure 12-6 | |
| Section 25.2.3.6: Secondary outcome 6: Completion of outcomes | Table 12-19 to Table 12-22 | |
| Section 25.2.3.7: Secondary outcome 7: Eligibility, recruitment and consent rates | Table 12-23 to Table 12-25 | |
| Section 25.2.3.8: Secondary outcome 8: Rates of successful follow-up | Table 12-26 and Table 12-27 | |
| Section 25.2.3.9: Secondary outcome 9: Summary statistics relating to antibiotic use in the pilot study with a comparison between the DTRs | Figure 12-7<br>Table 12-28 | |

### 16 Version history

Table 16-1 Version history

| Updated shell version no. | Shell section changed | Description of change | Date changed | Initials |
|---|---|---|---|---|
